# Links between gut microbiome composition and fatty liver disease in a large population sample

**DOI:** 10.1101/2020.07.30.20164962

**Authors:** Matti O. Ruuskanen, Fredrik Åberg, Ville Männistö, Aki S. Havulinna, Guillaume Méric, Yang Liu, Rohit Loomba, Yoshiki Vázquez-Baeza, Anupriya Tripathi, Liisa M. Valsta, Michael Inouye, Pekka Jousilahti, Veikko Salomaa, Mohit Jain, Rob Knight, Leo Lahti, Teemu J. Niiranen

**Author notes:** Correspondence: Matti Ruuskanen.

## Abstract

Fatty liver disease is the most common liver disease in the world. Its connection with the gut microbiome has been known for at least 80 years, but this association remains mostly unstudied in the general population because of underdiagnosis and small sample sizes. To address this knowledge gap, we studied the link between the Fatty Liver Index (FLI), a well-established proxy for fatty liver disease, and gut microbiome composition in a representative, ethnically homogeneous population sample of 6,269 Finnish participants. We based our models on biometric covariates and gut microbiome compositions from shallow metagenome sequencing. Our classification models could discriminate between individuals with a high FLI (≥ 60, indicates likely liver steatosis) and low FLI (< 60) in internal cross-region validation, consisting of 30% of the data not used in model training, with an average AUC of 0.75 and AUPRC of 0.56 (baseline at 0.30). In addition to age and sex, our models included differences in 11 microbial groups from class *Clostridia*, mostly belonging to orders *Lachnospirales* and *Oscillospirales*. Our models were also predictive of the high FLI group in a different Finnish cohort, consisting of 258 participants, with an average AUC of 0.77 and AUPRC of 0.51 (baseline at 0.21). Pathway analysis of representative genomes of the positively FLI-associated taxa in (NCBI) *Clostridium* subclusters IV and XIVa indicated the presence of *e*.*g*., ethanol fermentation pathways. These results support several findings from smaller case-control studies, such as the role of endogenous ethanol producers in the development of fatty liver.

## Introduction

Fatty liver disease affects roughly a quarter of the world’s population.^1^ It is characterized by accumulation of fat in the liver cells and is intimately linked with pathophysiology of metabolic syndrome.^2–4^ Fatty liver disease can be broadly divided into two variants: non-alcoholic fatty liver disease (NAFLD), attributed to high caloric intake, and alcohol associated fatty liver disease, attributed to high alcohol consumption. Even though the rate of progressions and underlying causes of both diseases might be different, they can be broadly sub-divided into those who have fat accumulation in the liver with no or minimal inflammation or those who have additional features of cellular injury and active inflammation with or without fibrosis typically seen in peri-sinusoidal area.^5^ Patients with steatohepatitis may progress to cirrhosis and hepatocellular carcinoma and have increased risk of liver-related morbidity and mortality, globally amounting to hundreds of thousands of deaths.^6^

The human gut harbors up to 10^12^ microbes per gram of content,^7^ and is intimately connected with the liver. Thus, it is no surprise that gut microbiome composition appears to have a strong connection with liver disease.^8^ Numerous studies over the past 80 years have reported associations between gut microbial composition and liver disease.^9^ For example, gut permeability and overgrowth of bacteria in the small intestine,^10^ changes in *Gammaproteobacteria* and *Erysipelotrichi* abundance during choline deficiency,^11^ elevated abundance of ethanol-producing bacteria,^12,13^ metagenomic signatures of specific bacterial species,^14,15^ have all been linked to NAFLD in small case-control patient samples. However, the microbial signatures often overlap between NAFLD and metabolic diseases, while those of more serious liver disease such as steatohepatitis and cirrhosis are more clear.^16^ For example, oral taxa appear to invade the gut in liver cirrhosis,^17^ and this phenotype can accurately be detected by analyzing the fecal microbiome composition (AUC = 0.87 in a validation cohort).^8^ Furthermore, we recently demonstrated good prediction accuracy for incident liver disease diagnoses (AUC = 0.83 for non-alcoholic liver disease, AUC = 0.96 for alcoholic liver disease, during ∼15 years),^18^ showing that the signatures of serious future liver disease are easy to detect.

The mechanisms underlying the contribution of gut microbiome content with fatty liver disease are thought to be primarily linked to gut bacterial metabolism. Bacterial metabolites can indeed be translocated from the gut through the intestinal barrier into the portal vein and transported to the liver, where they interact with liver cells, and can lead to inflammation and steatosis.^19^ Short-chain fatty acid production, conversion of choline into methylamines, modification of bile acids (BA) into secondary BA, and ethanol production, all of which are mediated by gut bacteria, are also known to be aggravating factors for NAFLD.^19^ Recent studies have also suggested that endogenous ethanol production by gut bacteria could lead to an increase in gut membrane permeability.^13^ This can facilitate the translocation of bacterial metabolites and cell components such as lipopolysaccharides from the gut to the liver, leading to further inflammation and possible development of NAFLD.^20^

Liver biopsy assessment is the current gold standard for diagnosis of fatty liver disease and its severity,^21^ but it is also impractical and unethical in a population-based setting. Ultrasound and MRI based assessment can help detect presence of fatty liver, however, this data is not available in our cohort. Regardless, recent studies have shown that indices based on anthropometric measurements and standard blood tests can be a reliable tool for non-invasive diagnosis of fatty liver particularly in population-based epidemiologic studies.^22,23^

Here, we designed and conducted computational analyses to examine the links between fatty liver and gut microbiome composition in a representative population sample of 7211 extensively phenotyped Finnish individuals.^24^ Because fatty liver disease is generally underdiagnosed in the general population,^25^ we used population-wide measurements of BMI, waist circumference, blood triglycerides and gamma-glutamyl transferase (GGT) to calculate a previously validated Fatty Liver Index (FLI) for each participant as a proxy for fatty liver.^26^ In parallel, we used shallow shotgun sequencing to analyze gut microbiome composition,^27^ which also enabled the use of phylogenetic and pathway prediction methods. In this work, we describe high-resolution associations between fatty liver and individual gut microbial taxa and clades, which are replicable in an external Finnish cohort, and thus generalizable in the Finnish population.

## Results

### Bacterial community structure is correlated with Fatty Liver Index in a population sample

In our main analyses, we classified our reads against the Genome Taxonomy Database (GTDB)^28^. This study mainly follows the GTDB taxonomy, unless otherwise noted. The Centrifuge/GTDB microbiome data used in our main analyses was based on archaeal and bacterial phylogenetic “balances”. This method was used to associate larger groups or clades of related organisms with fatty liver disease, and to avoid grouping of taxa on strict hierarchical taxonomic ranks featuring varying ranges of evolutionary divergence.^28^ Here, we used the PhILR transform, where each balance represents a single internal node in a phylogenetic tree, and its value is a log-ratio of the abundances of the two descending clades (for details, see methods and ref. ^29^). Positive values of the balance signify that the clade in the numerator is more abundant, and negative values that the clade in the denominator is more abundant. Thus, each association of a balance with the target variable necessarily includes both microbial clades descending from the node, one of them positively and the other negatively associated with the target variable. The clades in the numerator and denominator can be also freely switched by changing the sign of the balance value to retain the equivalence. Notably, we used this feature to show all balance-FLI associations in the positive direction to facilitate the comparison of their effect sizes (in **Figures S4, S7**, and **S9**).

Because the combined approach of using the GTDB taxonomy and the recently introduced PhILR -phylogenetic transform complicates the comparison of our results to previous studies, we also conducted more traditional statistical analyses with NCBI-annotated data to anchor our results to previous findings on the associations between fatty liver disease and gut microbiome composition. Overall, the Centrifuge/GTDB classification assigned 5.3 billion reads in the 6,269 samples (after exclusions in FINRISK 2002) to 23,457 bacterial and 1,248 archaeal taxa, and the SHOGUN/NCBI classification assigned 5.5 billion reads to 5,024 bacterial and 261 archaeal taxa. Starting from high level descriptions of the microbial communities in the high and low FLI groups (< 60 or ≥ 60 FLI; **Figure 1A**), the phylum-level distributions of bacterial and archaeal taxa appeared to be highly similar between the groups (**Figures S1, S2**). However, the proportion of taxa assigned to *Firmicutes* in Centrifuge/GTDB appeared to be slightly higher than in the SHOGUN/NCBI data. Furthermore, only 58% of the number of reads assigned in Centrifuge/GTDB to 6 main archaeal phyla were assigned to a single main archaeal phylum in SHOGUN/NCBI. Alpha diversity (as Shannon diversity) was significantly lower in the high FLI group, in both the SHOGUN/NCBI data (14.7% lower; AIC = 6685; all P < 1×10^−6^) and the Centrifuge/GTDB (13.4% lower; AIC = 6607; all *P* < 1×10^−4^) data, while adjusting for age, sex, and self-reported alcohol use in both models.

**Figure 1.**
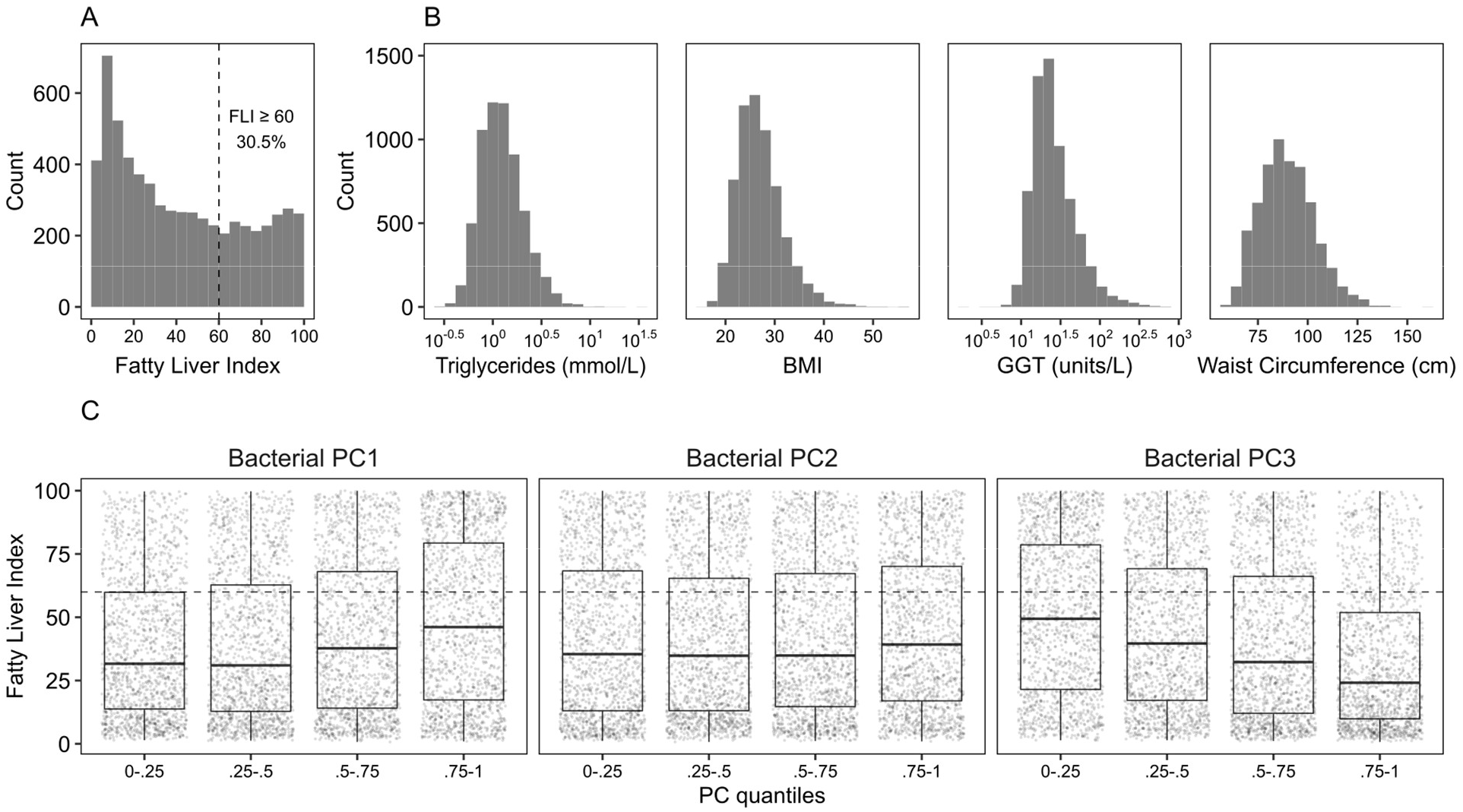
Relative of FLI (A), its components (B), and FLI in quantiles of the first three PC components of the fecal bacterial composition of the participants (C). The cutoff at FLI = 60 used to divide the participants is indicated with a dashed line in panels A and C.

To further examine the high-level associations between FLI (as a proxy of fatty liver disease) and microbial community composition in FINRISK 2002, we fit a linear regression model on the three first principal component (PC) axes of the fecal bacterial beta diversity (between individuals), sex, age, and alcohol. Log_10_(FLI) significantly correlated with all three bacterial PC axes, sex, age, and alcohol use in Centrifuge/GTDB data (adjusted R^2^ = 0.29; all *P* < 1×10^−6^), and PC1, PC3, sex, age, and alcohol use in SHOGUN/NCBI data (adjusted R^2^ = 0.27; all *P* < 1×10^−4^). Correlations between FLI and archaeal PC axes were not significant in Centrifuge/GTDB data (at the chosen significance level, *P* > 0.001), and between FLI and bacterial PC2 in SHOGUN/NCBI data (*P* > 0.001). In Centrifuge/GTDB data, the effect size estimate on log_10_(FLI) was a magnitude larger for PC1 (0.11 ± 0.008) than for PC2 (0.04 ± 0.008) and PC3 (−0.06 ± 0.008). The relationships between FLI and the bacterial PC components representing their beta diversity in Centrifuge/GTDB data are visualized for each of the three components in **Figure 1C**. A comparison of these relationships in Centrifuge/GTDB and SHOGUN/NCBI is included in the SI (**Figure S3**).

We also further assessed the phylogenetic balances contributing to the PC axes in the Centrifuge/GTDB data. Bacterial clades associated with higher FLI values, on the positive side of the balances contributing to PC1, included members of orders *Lachnospirales* and *Oscillospirales*, class *Bacilli*, and the *Ruminococcaceae, Bacteroidaceae* and *Lachnospiraceae* families (**Figure S4**). Several clades had a negative association with FLI, on the negative side of the balances contributing to PC1, such as order *Christensenellales* and genus *Faecalibacterium*. In addition, genus *Bifidobacterium* in PC2, and family *Bifidobacteriaceae* in PC3 had negative associations with continuous FLI.

### Several bacterial taxa are differently abundant between the low and high FLI groups

We also assessed significant differences in abundances of individual taxa between the high and low FLI groups in FINRISK 2002. In Centrifuge/GTDB data, we identified 244 taxa (1% of total) with an increased abundance, and 437 taxa (1.9%) with a decreased abundance in the high FLI group (all *Q* values < 0.001; **Table S7**). In SHOGUN/NCBI data, 80 taxa (1.6%) had an increased abundance, and 44 (0.9%) had a decreased abundance in the high FLI group (all *Q* values < 0.001). While the number of associated taxa was higher in the Centrifuge/GTDB data than SHOGUN/NCBI data, the proportion of significantly associated taxa was similar between the two methods. In both data sets, family *Lachnospiraceae* comprised over 40% of taxa positively associated with the high FLI group and *Bacterioidaceae* were in the top 3 most common families. The negatively associated taxa were much more diverse, but *Ruminococcaceae* and *Oscillospiraceae* were among the top 3 most common families in both data sets (at least > 6% of all negatively associated taxa).

### Bacterial lineages within the NCBI Clostridium subclusters IV and XIVa associate with FLI

Continuous FLI and differences between FLI groups in the FINRISK 2002 cohort (FLI < 60, *N* = 4,359 and FLI ≥ 60, *N* = 1,910; see **Figures 1A, 1B, Table S1**) were modeled with gradient boosting regression or classification using Leave-One-Group-Out Cross-Validation (LOGOCV) between participants from different regions. Only the bacterial PhILR transformed Centrifuge/GTDB data were used here, to find robust associations between phylogenetically related bacterial clades and fatty liver disease (instead of single taxa).

After feature selection and Bayesian hyperparameter optimization, the correlation between the predictions of the final regression models (age, sex, self-reported alcohol use, and 18 bacterial balances as features; each trained on the data from 5/6 regions) and true values in unseen data from the omitted region averaged R^2^ = 0.30 (0.26 – 0.33). After feature selection and optimization, the main classification models (age, sex, and 11 bacterial balances as features; each trained on the data from 5/6 regions) averaged AUC = 0.75 (**Table S2**) and AUPRC = 0.56 (baseline at 0.30; **Table S3**) on (unseen) test data from the omitted region. Models trained using only the covariates averaged AUC = 0.71 (AUPRC = 0.47) and using only the 11 bacterial balances they averaged AUC = 0.66 (AUPRC = 0.47) on test data. Alternative models were constructed by excluding participants with FLI between 30 and 60 (N = 1,583) and discerning between groups of FLI < 30 (N = 2,776) and FLI ≥ 60 (N = 1,910). These models averaged AUC = 0.80 (AUPRC = 0.75, baseline at 0.41) on their respective test data (**Tables S2, S3**). They averaged AUC = 0.76 (AUPRC = 0.68) when using only the covariates, and AUC = 0.70 (AUPRC = 0.63) when using only the 20 bacterial balances.

Because training data from all 6 regions was used to prevent overfitting in the selection of core features for all of the models, and similarly in searching for common hyperparameters, participants from the validation region of each model (in the training partition) partly influenced these parameters. Thus, we also constructed classification models discerning between the FLI < 60 and FLI ≥ 60 groups, where data of the validation region was completely excluded in the feature selection and hyperparameter optimization of each LOGOCV model. These models, using their individual feature sets and hyperparameters, averaged AUC = 0.75 and AUPRC = 0.57 (baseline at 0.30) on test data from their respective validation regions (**Table S4**). Using only covariates, they averaged AUC = 0.71 (AUPRC = 0.47), and AUC = 0.67 (AUPRC = 0.48) with only the bacterial balances.

Our external validation data consisted of 258 participants after exclusion of pregnant participants or those on antibiotics in the past 6 months, in the FINRISK 2007 population cohort^30^ (**Table S1, Figure S5**). The participants originate from North Karelia and Helsinki/Vantaa regions in Finland, and their samples were processed with the same methodology as was used for FINRISK 2002 (with Centrifuge/GTDB approach and PhILR). In this external validation, the 6 full models trained with covariates and the 11 bacterial balances in FINRISK 2002 averaged AUC = 0.77 (AUPRC = 0.51, baseline at 0.21; **Table S5**). The covariate-only models averaged AUC = 0.72 (AUPRC = 0.40) and the balance-only models averaged AUC = 0.69 (AUPRC = 0.44). The receiver operating characteristic and precision-recall curves based on the averaged predictions of the models, tested on this external validation data, also display good predictive ability (AUC = 0.78, AUPRC = 0.51 with baseline at 0.51; **Figure S6**)

To facilitate interpretability of the results, we continued examining the main classification models using a common set of core features. In these models, the median effect sizes of the features on the model predictions at their minimum and maximum values were highest for age, followed by sex, and the 11 balances in the phylogenetic tree (**Figures S7, S8**). All 11 associated balances were in phylum *Firmicutes*, class *Clostridia*, and largely in the NCBI *Clostridium* subclusters IV and XIVa (**Figure 2**). The specific taxa represented standardized GTDB genera (NCBI in brackets) *Negativibacillus* (*Clostridium*), *Clostridium M* (*Lachnoclostridium* / *Clostridium*), *CAG-81* (*Clostridium*), *Dorea* (*Merdimonas* / *Mordavella* / *Dorea* / *Clostridium* / *Eubacterium*), *Faecalicatena* (*Blautia* / *Ruminococcus* / *Clostridium*), *Blautia* (*Blautia*), *Sellimonas* (*Sellimonas* / *Drancourtella*), *Clostridium Q* (*Lachnoclostridium* [*Clostridium*]) and *Tyzzerella* (*Tyzzerella* / *Coprococcus*). Notably, all but one of the features in the main classification models (n226) were identified in the feature selection for the alternative models (constructed otherwise identically, but FLI < 30 was compared against FLI ≥ 60 in different data partitions), together with 10 additional balances (**Figure S9**). Only one of the balances in the alternative models was outside phylum *Firmicutes* (n1712 in *Bacteroidota*), and in addition, 4 balances were outside class *Clostridia* (n481 in *Negativicutes*; n826, n1009 and n918 in *Bacilli*). Also, negative associations with the high FLI group were seen for *An181 sp00216032*5 in the balance n266, where it is compared against the clade including *Dorea, Faecalicatena, Sellimonas* and *Tyzzerella* species (**Figures 2, S8**). A higher abundance of the clade including *Angelakisella, D5, Anaerotruncus* and *Phocea* species (against *Negativibacillus sp00435195* in balance n97) was also negatively associated with high FLI.

**Figure 2.**
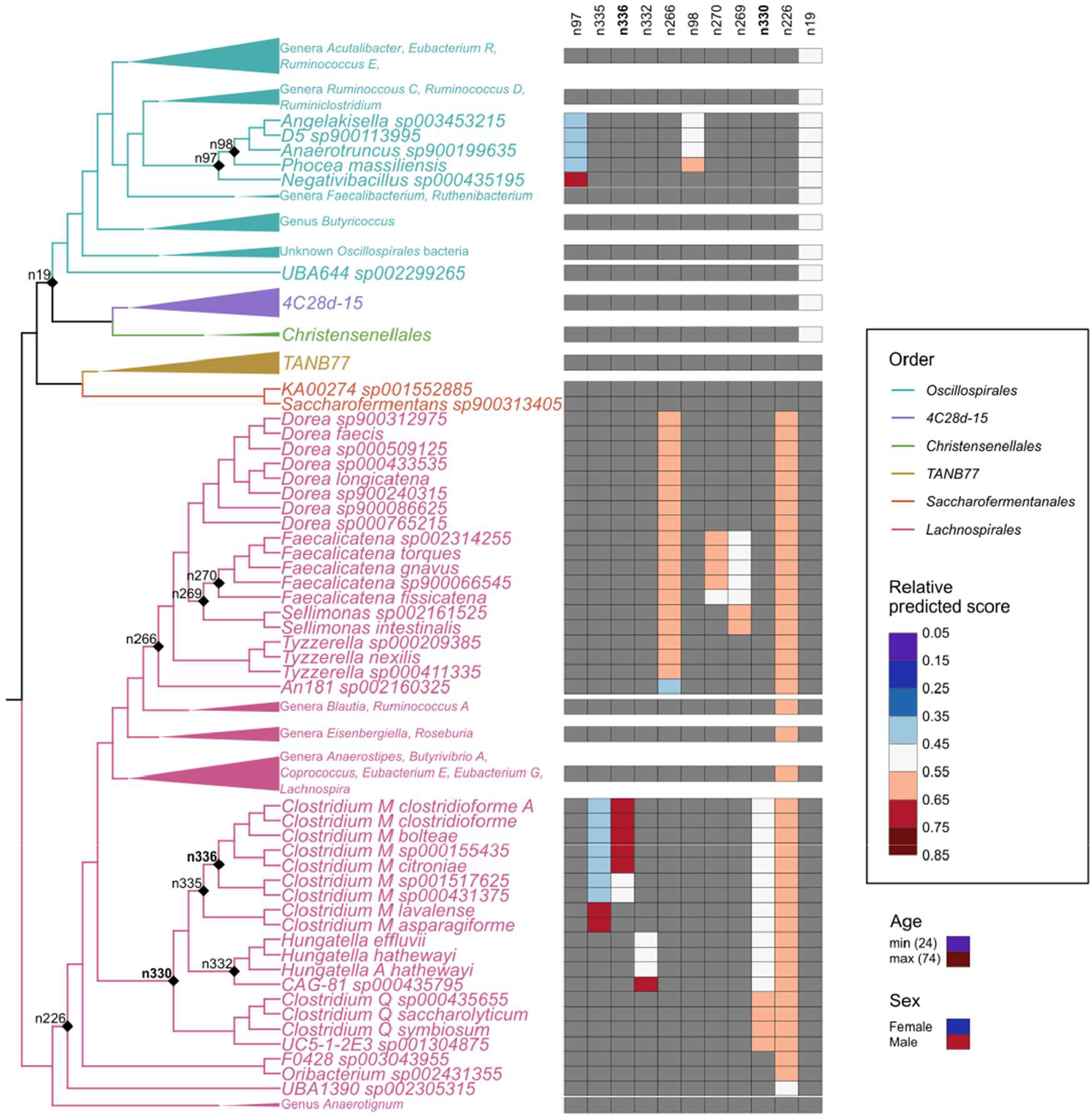
Relative effects of predictive balances and covariates on the FLI < 60 and FLI ≥ 60 classification model (AUC = 0.75) predictions. Nodes of the balances are indicated in the cladogram and the relative effect sizes of their clades (opposite sides of each balance) are shown in the associated heatmap. The relative effect sizes of the covariates (age and sex) are shown below the legend with a heatmap on the same scale as was used for the balances. The two liver-specific balances associated with triglyceride and GGT levels are indicated with bold font. Clades with redundant information have been collapsed but their major genera are indicated. The complete tree is included in **Figure S8**.

In addition to blood test results, FLI is based on two anthropometric markers linked to metabolic syndrome, waist circumference and BMI. This prompted us to dissect the Fatty Liver Index and identify which of the covariates and associated microbial balances from the phylogenetic tree can be linked to blood GGT and triglycerides measurements (see **Figure 1B**), and therefore would be most specific to hepatic steatosis and liver damage.^31^ To do so, we performed feature selection (similarly to continuous FLI) for GGT and triglycerides measurements in subsets of participants grouped by age, sex, and BMI. The feature selection identified two balances within the NCBI *Clostridia* XIVa subcluster (identified as n336 and n330) which were important for both GGT and triglyceride level prediction, and thus likely specific to liver function (**Figure 2**). Bacterial taxa were positively linked to liver function in these balances, and included (NCBI species) *Clostridium clostridioforme, C. bolteae, C. citroniae, C. saccharolyticum* and *C. symbiosum*. On the opposite, negatively associated side of the balances were, among others, (NCBI species) *Hungatella effluvii, H. hathewayi*, and two new GTDB-defined species *Clostridium M sp001517625* and *C. M sp000431375*.

### Ethanol and acetate production pathways are identified in representative bacterial genomes from taxa linked to high FLI

The values of predictive balances in the phylogenetic tree cannot be summarized for individual taxa, which means that only a qualitative investigation of the associations between their metabolism and fatty liver was possible in this study. We identified genetic pathways predicted to encode for SCFA (acetate, propanoate, butanoate) and ethanol production, BA metabolism, and choline degradation to trimethylamine (TMA) in representative genomes from the taxa we identified to be linked to liver function (**Figure S8**). These processes were chosen because they have been previously identified to have a mechanistic link to NAFLD (see *e*.*g*., ref. ^19^).

Acetate and ethanol production pathways appeared to be more common in the representative genomes of the taxa which had a positive association with FLI. In the liver function specific clades, n336 and n330, MetaCyc pathways for pyruvate fermentation to ethanol III (PWY-6587) and L-glutamate degradation V (via hydroxyglutarate; P162-PWY; produces acetate and butanoate) were present only in genomes positively associated with FLI. In balance n336, also heterolactic fermentation (P122-PWY; produces ethanol and lactate) was more often encoded in the clade positively associated with the high FLI group (3/5) than the opposing negatively associated clade (1/2). In representative genomes from the liver-specific balance n336, potential ethanol producers (PWY-6587) were seen in the positively associated clade (*Clostridum M clostridoforme A* and *Clostridum M sp000155435*), and not in the negatively associated clade (*Clostridium M sp001517625* and *Clostridium M sp000431375*). However, for most balances such trends were not clear in the qualitative analysis. Furthermore, we did not detect any of these pathways in the representative genomes of two individual taxa positively associated with FLI, *Negativibacillus sp000435195* and *Phocea massiliensis* (**Figure S8**).

## Discussion

The pathophysiology of fatty liver disease in general, and NAFLD in particular, is complex and its clinical diagnosis can be difficult.^32^ In this study, we utilized metagenomic data from a large population cohort (FINRISK 2002^30^) to identify broad links between the overall gut microbiome composition and fatty liver disease, using FLI as a recognized proxy (**Figure 1C**), and identified specific microbial taxa and lineages positively and negatively associated with the high FLI group (**Figure 2**). It should be noted, that FLI used in our study as a proxy for liver disease also includes features such as BMI and waist circumference, which associate with metabolic syndrome and diabetes.^16^ Links between these diseases and gut microbiome composition are well documented in previous studies.^33^ However, fatty liver disease is increasingly thought to be a component of the metabolic syndrome,^4,34^ and while diabetes prevalence is higher in the high FLI group in FINRISK 2002, affected participants still consist only 11% of this group (**Table S1**). Furthermore, we would like to emphasize that our results are not suitable for current clinical application, and should be validated by further, preferably mechanistic studies. We also do not know if our results generalize outside the Finnish population, as all participants in this study were exclusively from Finnish cohorts.

Considering that the predictive ability of FLI for clinically diagnosed NAFLD ranges between AUC = 0.81 – 0.93, in populations of Caucasian ethnicity such as the Finnish population,^23^ our models were able to reasonably predict the FLI group with AUC = 0.75 (AUPRC = 0.56, baseline at 0.30), in our internal cross-region validation. Furthermore, the performance of our predictive models was highly similar in an external, Finnish validation cohort (AUC = 0.77, AUPRC = 0.51, baseline at 0.21).

Our additional analyses support these main results. While a thorough method comparison is beyond the scope of the current study, the results from the two taxa assignments were very similar despite their differences, such as the fourfold higher number of taxa in the Centrifuge/GTDB data. In the machine learning models (performed only with Centrifuge/GTDB data), excluding participants with intermediate FLI (between 30 – 60) increased the accuracy slightly in the internal cross-validation (to AUC = 0.8 and AUPRC = 0.75, baseline at 0.41). However, discerning between participants with probable fatty liver disease (FLI ≥ 60) from others presents a clinically more relevant target for detecting changes in microbiome composition associated with development of the disease. In another set of models, we negated the influence of validation region data in the individual models also for feature selection and hyperparameter optimization during training. This led to individualized sets of features and parameters in the models, but the average performance of the models was almost identical on validation region samples in the internal cross-validation (AUC = 0.75 and AUPRC 0.57, baseline at 0.30). The aim of our study was to find patterns in microbiome composition which would be generalizable across the 6 sampled geographic regions in Finland and easy to interpret. Thus, we consider the use of all training data to define the common core feature set justified. This goal also guided our overall modeling architecture and likely led to a lower performance than if we instead performed interpolation within a smaller scale (see *e*.*g*., ref. ^35^).

When interpreting our results, several levels of associations can be considered according to types of fatty liver disease and the gut microbiome composition. Because FLI has been mostly validated with simple steatosis and NAFLD,^23,26^ we can conservatively contextualize our findings with previous associative work that used these diagnoses or clinical manifestations, only. The cutoff used in our study at FLI ≥ 60 has been used to rule in liver steatosis in a Caucasian cohort comparable to ours,^26^ but also a cutoff at FLI ≥ 48 has been found appropriate for simple steatosis in a Portuguese cohort.^36^ Much lower cutoffs (FLI ≥ 20 to 30) have been used in Asian cohorts.^37–39^ Thus, it is likely that our high FLI groups include most participants with liver steatosis or fibrosis in both FINRISK cohorts, but the low FLI group also likely includes participants with low grade steatosis.

### Traditional statistical analyses replicate previous findings on gut microbiome composition and fatty liver disease when using FLI as a risk index

Among the significant high level FLI-associated differences in the gut microbiomes of the participants in FINRISK 2002, we found a 14.7% lower Shannon alpha diversity in the high FLI group with SHOGUN/NCBI taxa assignments and 13.4% lower diversity with Centrifuge/GTDB assignments. These results are in good accordance with previous results of decreased gut bacterial diversity in patients with biopsy-proven non-alcoholic steatohepatitis (NASH), the most serious form of NAFLD.^40^ In this case-control study, the Shannon diversity of gut microbiomes in NASH patients without liver cirrhosis was on average 7% lower compared to controls, and in patients with cirrhosis, 14% lower. A significantly decreased gut microbiome alpha diversity of similar magnitude was also seen in cohort participants with persistent NAFLD compared to controls.^41^

In both the SHOGUN/NCBI and Centrifuge/GTDB data, we found significant linear correlations between FLI and beta diversity, or two or three main bacterial PC-axes of the samples, respectively (**Figures 1C, S3**). The model fit was slightly better with Centrifuge/GTDB data, which might be due to the higher number of identified taxa, and thus increased taxonomic resolution (although including putative species in GTDB). Our results support previous observations of differences in beta diversity in relation to persistent NAFLD,^41^ and along the NAFLD-cirrhosis spectrum.^8^ Through the loadings of the phylogenetic balances on the PC axes in the Centrifuge/GTDB data, we detected several previously known connections between microbial clades and FLI (**Figure S4**). Among others, we observed a positive association between high FLI and family *Lachnospiraceae* and negative associations for order *Christensenellales*, genus *Faecalibacterium*, and genus *Bifidobacterium*. The positive association is supported by previous findings of their connection with obesity,^42^ and the negative associations by connections to lean individuals and healthy gut microbiome composition.^43–45^

Our differential abundance analysis also detected a high number of taxa with significantly increased or decreased abundance in the high FLI group. All following results were observed both in the Centrifuge/GTDB and SHOGUN/NCBI data sets, unless otherwise noted. Majority of the taxa with increased abundance in the high FLI group were from family *Lachnospiraceae*, which supports their positive association with NAFLD reported previously in a number of studies,^46^ but also with obesity (**Table S7**).^42^ The increased abundance of genus *Roseburia* has also been highlighted as a characteristic change in gut microbiome related to NAFLD.^46,47^ In the current study, two members of genus *Roseburia* were in the top 10 taxa most strongly associated with high FLI. Furthermore, our results support previous findings on the positive associations of, for example, *Collinsella*,^40^ *Prevotella copri*,^48^ *Dorea*,^47^ with NAFLD. We also detected increases in *Sutterella* and *Streptococcus*, previously associated with cirrhosis.^49^ However, we did not find increases in families *Kiloniellaceae* and *Pasteurellaceae*, previously associated with NAFLD.^46^ Among the individual taxa negatively associated with high FLI, families *Ruminococcaceae* and *Oscillospiraceae* (such as genus *Oscillibacter*) were common, which supports previous findings on their connections with NAFLD.^12,41,46^ A high number of putative (GTDB) species were negatively associated with FLI in the Centrifuge/GTDB data, which were understandably not present in the SHOGUN/NCBI data. Many of these were classified in the recently described order *Christensenellales*,^28^ including families such as CAG-74, associated with healthy participants,^50^ and *Christensenellaceae*, which are widespread, highly heritable, and associated with health.^44,51^

While our results from common statistical experiments mainly supported previous findings, we chose to leverage the phylogenetic information included in the GTDB data to find robust associations between larger bacterial clades and fatty liver disease in the Finnish population. This was accomplished by constructing predictive models to classify participants in the FLI groups based on the phylogenetic balances and covariates, subjected to feature selection and geographical cross-validation.

### Predictive modeling of FLI reveals consistent associations between gram-positive Clostridia and fatty liver disease

Strikingly, the strongest associations with FLI in our machine learning models were all inside the *Firmicutes* phylum. A possible reason for this might be the higher relative abundance of phylum *Firmicutes* at high latitudes,^52^ where Finland is. Among the associations we identified, *Faecalicatena gnavus* (NCBI: *Ruminococcus gnavus*) was positively linked with FLI as part of 3 predictive balances, and associated in previous studies with liver cirrhosis.^17^ In their study, oral *Firmicutes*, such as *Veillonella*, were suggested to invade the gut. While our balance-based approach did not detect these taxa, *Megasphaera elsdenii* were positively associated with the high FLI group in our differential abundance analyses (**Table S7**). This might be due to the strict feature selection employed prior to the predictive modeling.

Two individual taxa, *Negativibacillus sp000435195* and *Phocea massiliensis*, both had strong positive associations with the high FLI group (**Figure 2**), but the balances including these species were not predictive of the liver function-specific components (triglycerides and GGT). Positive associations of these taxa with fatty liver disease have not been documented previously. However, a decreasing abundance of both bacteria, *Negativibacillus sp000435195* (NCBI: *Clostridium* sp. CAG:169) and *Phocea massiliensis* (NCBI: *Phocea massiliensis*), were seen when the intake of meat and refined cereal was reduced isocalorically in favor of fruit, vegetables, wholegrain cereal, legumes, fish and nuts in overweight and obese subjects in Italy.^53^ While comparisons between these studies are difficult due to differences in taxa annotations, bacteria such as *Faecalicatena gnavus* (NCBI: *Ruminococcus gnavus*) and *Clostridium Q saccharolyticum* (NCBI: *Clostridium saccharolyticum*) were also found to respond negatively to the Mediterranean diet. Thus, further study on the connections of these bacteria with gut health and diet is warranted.

Among the taxa negatively associated with high FLI, *Hungatella* (see balance n332, **Figure 2**) have been previously shown to correlate negatively with the obesity phenotype in mice^54^ and *H. hathewayi* was found to be a common commensal in the gut of healthy volunteers.^55^ However, genus *Hungatella* has also been positively associated with concentrations of trimethylamine-N-oxide (TMAO),^56^ a metabolite associated with cardiovascular disease and NAFLD. In our study, on the positively associated side (of balance n332) opposite to genus *Hungatella* was a novel GTDB species, *CAG-81 sp000435795*, previously included in NCBI genus *Clostridium*. The *CAG-81* genus was recently positively associated with TMAO levels in urine in a study using the GTDB classification.^57^ While we did not find the pathway for TMA (precursor to TMAO) production in its genome, this would explain the positive association of the *CAG-81* species with high FLI. Furthermore, the previous contradictive results among these taxa could be explained by grouping of putatively TMA producing taxa in *CAG-81* together with the closely related genus *Hungatella*.

Most taxa in our study with a positive association with FLI belonged to the broadly defined *Clostridium* NCBI genus, which supports several previous observations.^14,46,58^ However, taxonomic standardization according to GTDB has identified the *Clostridium* genus as the most phylogenetically inconsistent of all bacterial genera in the NCBI taxonomy, and divides it into a total of 121 monophyletic genera in 29 distinct families.^28^ The GTDB reassignment complicates comparisons to previous studies, but it is phylogenetically and biologically sensible, and can thus provide new insights into the microbiomes. Our results also strongly suggest that despite its higher cost compared to metabarcoding, the increased resolution of (shallow) shotgun metagenomic sequencing is highly useful in identifying specific taxon-disease associations (see *e*.*g*., refs. ^27,59^).

### Bacterial taxa positively associated with high FLI have a genetic potential to exacerbate the development of fatty liver disease

We identified several plausible new associations between individual taxa and clades of bacteria and fatty liver. All taxa were from class *Clostridia*, which are obligate anaerobes. We observed that reference genomes from the bacterial taxa positively associated with high FLI in the liver-specific balances harbored several genetic pathways necessary for ethanol production.

Specifically, genes predicted to enable the fermentation of pyruvate to ethanol (MetaCyc PWY-6587) appeared to be common. Endogenous production of ethanol has been known to both induce hepatic steatosis and increase intestinal permeability,^60^ and several of the taxa associated with the high FLI group have also been experimentally shown to produce ethanol, such as *C. M asparagiforme, C. M bolteae, C. M clostridioforme / C. M clostridioforme A* ^61^, and *C. Q Saccharolyticum*.^62^ The relative abundances of these putatively ethanol-producing taxa were also predictive of FLI groups in previously unseen data. However, the self-reported alcohol consumption from the participants was not among the best predictors for the FLI groups, as it was excluded in the feature selection step.

All reference genomes from taxa positively associated with FLI in balance n330 harbored genes predicted to encode for the L-glutamate fermentation V (P162-PWY; **Figure S8**) pathway, which results in the production of acetate and butanoate. Glutamate fermentation could lead to increased microbial protein fermentation in the gut, which has been previously been linked with obesity, diabetes and NAFLD.^63^ Recently, the combined intake of fructose and microbial acetate production in the gut was experimentally observed to contribute to lipogenesis in the liver in a mouse model.^64^ Interestingly, *C. Q saccharolyticum* (in our study, a taxa positively associated with high FLI deriving from balance n330), was experimentally shown to ferment various carbohydrates, including fructose, to acetate, hydrogen, carbon dioxide, and ethanol.^62^ Furthermore, while our own pathway analysis did not detect BA modification pathways in the reference genome of *C. Q saccharolyticum*, a strain of this species has been highlighted as a probable contributor to NAFLD development through the synthesis of secondary BA.^15^ The links between dietary intake and gene regulation, combined with microbial fermentation in the gut warrant further mechanistic experiments to elucidate their links with fatty liver, and likely other metabolic diseases.

NAFLD-associated ethanol producing bacteria in previous cohort studies have all been gram-negatives, such as (NCBI-defined) *Klebsiella pneumoniae*,^13^ and *Escherichia coli*.^12^ In our population sample, instead of gram-negatives, bacteria from the *C. M bolteae, C. M clostridioforme / C. M clostridioforme A* and *C. M citroniae* species (positively associated with high FLI in balance n336) have been described as opportunistic pathogens,^65^ and are hypothesized to exacerbate fatty liver development similarly through endogenous ethanol production. This result suggests that geographical,^35^ and ethnic variability,^66^ might also strongly affect gut microbiome composition and its associations with disease. In addition to putative endogenous ethanol producers, we identified other taxa positively associated with high FLI in balance n330, for which reference genomes harbored a genetic pathway predicted to encode for the ability to ferment L-lysine to acetate and butyrate. While the production of these SCFAs is often considered beneficial for gut health, other metabolism of proteolytic bacteria might negatively contribute to fatty liver disease.^67^

Through modeling a previously validated index for fatty liver, FLI, we found replicable associations with specific microbial taxa and likely liver disease of the participants. In addition, sex and age of participants were also strongly predictive of the FLI group in our models (**Figures 2, S7**). Their similar positive associations with fatty liver disease are known from previous studies.^68,69^ The associated microbial balances could be used to improve the predictions above the baseline of these covariates on 5/6 regions in Finland in the main cohort. For example, in the model cross-validated with Lapland the balances were more predictive of FLI group than the covariates by themselves, and their combination increased the AUC further. Yet, when testing the model where Turku/Loimaa region was used for internal cross-validation, the microbial balances were slightly predictive of FLI group but failed to improve the AUC over the covariates (**Table S2**). This pattern might stem from the cultural and genetic west-east division in Finland,^70,71^ with a closer proximity of the Helsinki/Vantaa region to eastern regions than Turku/Loimaa, in both terms. It is thus likely that further incorporation and investigation on the use of spatial information in microbiome modeling would elucidate these geographical patterns in taxa-disease associations.

Our models were also able to accurately predict the FLI group of participants in the external validation cohort, which were from the North Karelia and Helsinki/Vantaa regions. The observed difficulty to geographically extrapolate taxa-disease associations^35^ might mean that associations reported in our study are specific to Finland and nearby regions. Notably, many of the positive associations between specific taxa and fatty liver disease have not been reported previously, but the functional potential of these taxa inferred from genomic data is similar to taxa positively associated with NAFLD in previous studies. Thus, the geographical limits of taxa-disease associations reported in studies such as ours warrant further study. Unfortunately, generalization of our own results outside of Finland also remains to be addressed.

It is likely that not all associations in the current study are related solely to liver steatosis, because FLI is based on measurements related to metabolic syndrome. However, our approach is supported by recent views of NAFLD as the integral liver component of the metabolic syndrome.^34,72^ Indeed, the prevalences of diabetes and cardiovascular disease in both FINRISK 2002 and 2007 cohorts are elevated in the high FLI group, although the majority of the high FLI participants did not have either of these diagnoses at the time of sampling (**Table S1**). We also dissected the FLI by dividing participants into age/sex/BMI groups and detected microbial groups specific to the blood work measurements of liver damage, triglycerides and GGT. These associated taxa can thus be thought of as most closely associated with liver function, if such a division is deemed practical.

## Conclusions

Modeling an established risk index for fatty liver enabled the detection of associations between the disease and gut microbiome composition, to the level of individual taxa. While utilizing FLI as a proxy, NCBI taxa identified with standard statistical methods were supportive of previously reported differences between NAFLD cases and healthy controls. In our machine learning framework, all clades robustly predictive of the FLI group were from the obligately anaerobic gram-positive class *Clostridia*, representing several redefined GTDB genera previously included in the NCBI genus *Clostridium*. Many of the representative genomes of taxa positively associated with high FLI had genomic potential for endogenous ethanol production. Our results support previous findings on the likely contribution of ethanol and increased gut permeability on the induction of hepatic steatosis. Further support was also found for the involvement TMA and SCFAs, especially acetate, in the likely pathophysiology of fatty liver disease. Our models were able to predict the FLI group of participants in Finland across geographical regions and in an external Finnish cohort, showing that the associations are robust and generalizable in this population. Based on our results, mechanistic connections between specific microbes and fatty liver disease, and the geographical differences in such taxa-disease associations, should be addressed in further studies.

## Materials and Methods

### Survey details and sample collection

Cardiovascular disease risk factors have been monitored in Finland since 1972 by conducting a representative population survey every five years.^30^ In the FINRISK 2002 survey, a stratified random population sample was conducted on six geographical regions in Finland. These are North Karelia and Northern Savo in eastern Finland, Turku and Loimaa regions in southwestern Finland, the cities of Helsinki and Vantaa in the capital region, the provinces of Northern Ostrobothnia and Kainuu in northwestern Finland, and the province of Lapland in northern Finland.

Briefly, at baseline examination the participants filled out a questionnaire form, and trained nurses carried out a physical examination and blood sampling in local health centers or other survey sites. Data was collected for physiological measures, biomarkers, and dietary, demographic and lifestyle factors. Stool samples were collected by giving willing participants a stool sampling kit with detailed instructions. These samples were mailed overnight between Monday and Thursday under Finnish winter conditions to the laboratory of the Finnish Institute for Health and Welfare, where they were stored at -20°C. In 2017, the samples were shipped still unthawed to University of California San Diego for microbiome sequencing.

Details of the FINRISK cohorts analyzed in this study are included in the supplementary files (**Table S1**). Further details and sampling have also been extensively covered in previous publications (see refs. ^24,73^). The Coordinating Ethics Committee of the Helsinki University Hospital District approved the study protocol for FINRISK 2002 (Ref. 558/E3/2001), and all participants have given their written informed consent.

### Stool DNA extraction and shallow shotgun metagenome sequencing

DNA extraction was performed according to the Earth Microbiome Project protocols, with the MagAttract PowerSoil DNA kit (Qiagen), as previously described.^74^ A miniaturized version of the Kapa HyperPlus Illumina-compatible library prep kit (Kapa Biosystems) was used for library generation, following the previously published protocol.^75^ DNA extracts were normalized to 5 ng total input per sample in an Echo 550 acoustic liquid handling robot (Labcyte Inc.). A Mosquito HV liquid-handling robot (TTP Labtech Inc.) was used for 1/10 scale enzymatic fragmentation, end-repair, and adapter-ligation reactions. Sequencing adapters were based on the iTru protocol,^76^ in which short universal adapter stubs are ligated first and then sample-specific barcoded sequences added in a subsequent PCR step. Amplified and barcoded libraries were then quantified by the PicoGreen assay and pooled in approximately equimolar ratios before being sequenced on an Illumina HiSeq 4000 instrument to an average read count of approximately 900,000 reads per sample.

### Taxonomic matching and phylogenetic transforms

We quality trimmed the sequences and removed the sequencing adapters with Atropos.^77^ Host reads were removed by mapping the reads against the human genome assembly GRCh38 with Bowtie2.^78^ To improve the taxonomic assignments of our reads, we used a custom index,^79^ based on the Genome Taxonomy Database (GTDB) release 89 taxonomic redefinitions,^28,80^ for read classification with default parameters in Centrifuge 1.0.4.^81^ Viral and eukaryotic sequences were removed in this step, as the database contains only bacterial and archaeal reference genomes. After read classification, all following steps were performed with R version 3.5.2,^82^ using phyloseq 1.30.0^83^ to manage the data. To reduce the number of spurious read assignments, and to facilitate more accurate phylogenetic transformations, only reads classified at the species level, matching individual GTDB reference genomes, were retained. Samples with less than 50,000 reads, from pregnant participants or recorded antibiotic use in the past 6 months were removed, resulting in a final number of 6,269 samples. We first filtered taxa not seen with more than 3 counts in at least 1% of samples and those with a coefficient of variation ≤ 3 across all samples, following McMurdie and Holmes^83^, with a slight adaption from 20% of samples to 1% of samples, because of our larger sample size. The complete bacterial and archaeal phylogenetic trees of the GTDB release 89 reference genomes, constructed from an alignment of 120 bacterial or 122 archaeal marker genes,^28^ were then combined with our taxa tables. The resulting trees were thus subset only to species which were observed in at least one sample in our data. The read counts were transformed to phylogenetic node balances in both trees with PhILR.^29^ The default method for PhILR inputs a pseudocount of 1 for taxa absent in an individual sample before the transform.

In this study, we did not specifically and solely use relative abundances at various taxonomic levels, as is common practice for microbiome studies. Instead, we applied a PhILR transformation to our microbial composition data,^29^ introducing the concept of microbial “balances”. Indeed, evolutionary relationships of all species harbored in each microbiome sample can be represented on a phylogenetic tree, with species typically shown as external nodes that are related to each other by multiple branches connected by internal nodes. In this context, the value of a given microbial “balance” is defined as the log-ratio of the geometric mean abundance between two groups of microbes descending from the same corresponding internal node on a microbial phylogenetic tree. This phylogenetic transform was used because it i) addresses the compositionality of the metagenomic read data,^84^ ii) permits simultaneous comparison of all clades without merging the taxa by predefined taxonomic levels, and iii) enables evolutionary insights into the microbial community. The links between microbes and their environment, such as the human gut, is mediated by their functions. Different functions are known to be conserved at different taxonomic resolutions, and most often at multiple different resolutions.^85^ Thus, associations between the microbes and the response variable are likely not best explained by predefined taxonomic levels. In the absence of functional data, concurrently analyzing all clades (partitioned by the nodes in the phylogenetic tree) would likely enable the detection of the associations at the appropriate resolution depending on the function and the local tree topography.

To further validate our approach, assess how the use of the GTDB taxonomic redefinitions and custom database affected our results, and to facilitate comparisons with previous results, we annotated our raw reads in FINRISK 2002 samples also with NCBI taxonomy and performed several additional analyses. For this comparison data, after quality trimming the FINRISK 2002 reads and removing host sequences as described above, SHOGUN v1.0.5^59^ was used for taxonomy assignments against the NCBI RefSeq version 82 (May 8, 2017) database containing complete bacterial, archaeal, and viral genomes. To facilitate comparisons between different annotations, we subset the samples included in the SHOGUN/NCBI annotated data to those included in the Centrifuge/GTDB data (for exclusion criteria, see above).

### Covariates

Because fatty liver disease is underdiagnosed at the population level,^25^ and our sampling did not have extensive coverage of liver fat measurements, we chose to use the Fatty Liver index as a proxy for fatty liver.^26^ Furthermore, the index performs well in cohorts of Caucasian ethnicity, such as ours, to diagnose the presence of NAFLD.^23^ We calculated FLI after Bedogni et al.^26^: 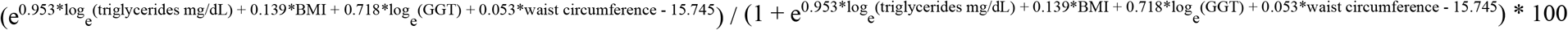. We chose the cutoff at FLI ≥ 60 to identify participants likely to be diagnosed with hepatic steatosis (positive likelihood ratio = 4.3 and negative likelihood ratio = 0.5, after Bedogni et al.^26^). Triglycerides, gamma-glutamyl transferase (GGT), BMI and waist circumference measurements had near complete coverage for the participants in our data. Self-reported alcohol use was calculated as grams of pure ethanol per week. Cases with missing values were omitted in linear regression models. At least one feature used for FLI calculation was missing for 20 participants in FINRISK 2002 (0.3%) and the self-reported alcohol use was missing for 247 participants (3.9%). In the machine learning framework, missing values for FLI and self-reported alcohol use were mean imputed. However, for the feature selection to identify liver function-specific balances, GGT, triglycerides and BMI were not imputed but observations where any of these were missing were simply removed.

### Taxa composition and alpha diversity

The baseline compositions of the microbial communities in the samples were summarized at phylum level in the different FLI groups (< 60 and ≥ 60 FLI) with the Centrifuge/GTDB data in FINRISK 2002 and 2007 (**Figure S1**), and with SHOGUN/NCBI data in FINRISK 2002 (**Figure S2**) by total sum scaling and merging taxa at phylum level, separately for bacteria and archaea.

Bacterial alpha diversity of each individual sample in FINRISK 2002 was estimated through Shannon diversity as the mean of 10 random rarefactions of raw annotated read counts (see ref. ^86^), separately in both the Centrifuge/GTDB and SHOGUN/NCBI data sets. Associations between the FLI group (< 60 or ≥ 60 FLI) and Shannon diversity in the data sets was modeled using binomial regression and adjusted for age, sex, and self-reported alcohol use, using “glm” in base R.^82^

### Beta diversity and linear modeling of FLI

In the Centrifuge/GTDB data, beta diversity was calculated as Euclidian distance of the PhILR balances through Principal Component Analysis (PCA) on bacterial and archaeal balances separately with “rda” in vegan 2.5.6.^87^ To calculate beta diversity with the SHOGUN/GTDB data, raw bacterial taxa counts were centered log-ratio (CLR) transformed with “transform” in microbiome 1.8.0,^88^ and their Euclidian distances were obtained similarly through PCA. Linear regression models were constructed for FLI with “lm” in base R^82^ with Centrifuge/GTDB data, and separately with SHOGUN/NCBI. Log_10_(FLI) was used as the dependent variable and the first three bacterial PCs, sex, age, and self-reported alcohol were used as the independent variables. Archaeal PCs were not included in the models because none of them were significantly correlated with FLI in Centrifuge/GTDB data (all *P* > 0.001). To visualize the association between beta diversity and FLI, the FLI of each participant was plotted against its quantiles along the three bacterial PC axes in Centrifuge/GTDB data (**Figure 1C**). A comparison of the associations with the alternative SHOGUN/NCBI annotated data was also included in the SI (**Figure S3**).

### Differential abundance of individual taxa between the FLI groups

To facilitate comparisons to previous studies, we assessed the associations between the FLI group (< 60 or ≥ 60 FLI) of the participants and Centrifuge/GTDB and SHOGUN/NCBI annotated individual taxa present in the samples. With both data sets, differential abundance of the bacterial taxa between the FLI groups was assessed with the ALDEx2 compositional data analysis tool.^89^ Briefly, significance of the abundance differences between the groups were estimated with a Welch’s t-test, and only taxa with (Benjamini Hochberg) false discovery rate - adjusted *P* values (or *Q* values) < 0.001 were retained. The associated taxa were then divided in each data set to those positively or negatively associated with the high FLI group and sorted based on effect sizes estimated from the median CLR differences between the groups.

### FLI modeling within a machine learning framework

In the machine learning framework, both regression and categorical models were constructed for FLI, using only the Centrifuge/GTDB data. The feature selection, hyperparameter optimization and internal cross-validation methods were identical for both approaches, unless otherwise stated. The continuous or categorical FLI (groups of FLI < 60 and FLI ≥ 60) were modeled with xgboost 0.90.0.2,^90^ by using both bacterial and archaeal balances, sex, age, and self-reported alcohol use as preliminary predictor features. We used FLI 60 as the cutoff for ruling in fatty liver (steatosis) for the classification, after Bedogni et al., (2006).^26^ The data was first split to 70% train and 30% test sets while preserving sex and region balance. To take into account geographical differences (see *e*.*g*., ref. ^35^) and to find robust patterns across all 6 sampled regions in Finland between the features and FLI group, we used Leave-One-Group-Out Cross-Validation (LOGOCV) inside the 70% train set to construct 6 separate models in each optimization step. Because of high dimensionality of the data (3,423 predictor features) feature selection by filtering was first performed inside the training data, based on random forest permutation as recommended by Bommert et al.^91^ Briefly, permutation importance is based on accuracy, or specifically the difference in accuracy between real and permuted (random) values of the specific variable, averaged in all trees across the whole random forest. The permutation importance in models based on the 6 LOGOCV subsets of the training data were calculated with mlr 2.16.0,^92^ and the simple intersect between the top 50 features in all LOGOCV subsets were retained as the final set of features. Thus, the feature selection was influenced by the training data from all 6 geographical regions, but this only serves to limit the number of chosen features because of the required simple intersect. This approach was used to obtain a set of core predictive features which would have potential for generalizability across the regions. The number of features included in the models by this approach was deemed appropriate, since the relative effect size of the last included predictor was very small (< 0.1 change in classification probability across its range).

Bayesian hyperparameter optimization of the xgboost models was then performed with only the selected features. An optimal set of parameters for the xgboost models were searched over all LOGOCV subsets with “mbo” in mlrMBO 1.1.3,^93^ using 30 preliminary rounds with randomized parameters, followed by 100 optimization rounds. Parameters in the xgboost models and their considered ranges were learning rate (eta) [0.001, 0.3], gamma [0.1, 5], maximum depth of a tree [2, 8], minimum child weight [1, 10], fraction of data subsampled per each iteration [0.2, 0.8], fraction of columns subsampled per tree [0.2, 0.9], and maximum number of iterations (nrounds) [50, 5000]. The parameters recommended by these searchers were as following for regression: eta=0.00889; gamma=2.08; max_depth=2; min_child_weight=8; subsample=0.783; colsample_bytree=0.672; nrounds=1,810, and for classification: eta=0.00107; gamma=0.137; max_depth=5; min_child_weight=9; subsample=0.207; colsample_bytree=0.793; nrounds=4,328. We used Root-Mean-Square Error (RMSE) for the regression models and Area Under the ROC Curve (AUC) for the classification models to measure model fit on the left-out data (region) in each LOGOCV subset. Receiver operating characteristic and precision-recall curves for these validation metrics were calculated with “evalmod” in precrec v0.11.2.^94^ The final models were trained on the LOGOCV subset training data, the data from one region thus omitted per model, and using the selected features and optimized hyperparameters. Internal validation of these models was conducted against participants only from the region omitted from each model, in the 30% test data which was not used in model training or optimization. Sensitivity analysis was conducted by using only the predictive covariates (sex and age) or balances separately, with the same hyperparameters, data partitions and cross-region internal validation as for the full models.

### Partial dependence interpretation of the FLI classification models

Because the classification models have a more clinically relevant modeling target for the difference between FLI < 60 and FLI ≥ 60, the latter used to rule in fatty liver,^26^ we further interpreted the partial dependence of their predictions. Partial dependence of the classification model predictions on individual features was calculated with “partial” in pdp 0.7.0.^95^ The partial dependence of the features on the model predictions was also plotted, overlaying the results from each of the 6 models. For each feature, its relative effect on the model prediction was estimated as medians of the minimum and maximum yhat (output probability of the model for the FLI ≥ 60 class), calculated at the minimum and maximum values of the feature separately in each of the 6 models. The relative effects of the balances were then overlaid as a heatmap on a genome cladogram which covers all balances in the model with ggtree 2.1.1.^96^

### Construction of alternative classification models to discern between FLI < 30 and FLI ≥ 60 groups

To assess robustness of the models and how removing the participants with intermediate FLI (between 30 and 60) affects model performance, we removed this group (*N* = 1910) and constructed alternative classification models to discern between the FLI < 30 and FLI ≥ 60 groups. Other than removing the intermediate FLI participants and resulting new random split to the train (70%) and test (30%) sets, these models were constructed identically to the main models, including LOGOCV design, feature selection, and hyperparameter optimization. The recommended parameters for this classification task were eta=0.00102; gamma=3.7; max_depth=2; min_child_weight=5; subsample=0.49; colsample_bytree=0.631; nrounds=3,119. Interpretation of partial dependence was also performed identically, but only the relative effects of the model features were plotted without a cladogram.

### Exclusion of validation region data from feature selection and hyperparameter optimization

Because training data from all 6 regions is used to inform the selection of optimal features and hyperparameters, the validation region data cannot be considered completely independent from the training of the LOGOCV models. Thus, we constructed a set of classification models for the FLI ≥ 60 and FLI < 60 groups, where all validation region samples also in the training data were excluded from the simple intercept of top 50 features in each LOGOCV set and from the subsequent hyperparameter optimization. These models with individualized features and hyperparameters were then tested on the validation region samples in the unseen test data to estimate how model performance was affected. The main test (70%) and train (30%) sets were identical to the main models, but additionally 6 randomized 70/30 splits nested inside the test set (excluding the validation region) were used in hyperparameter optimization to reduce overfitting. Average optimal hyperparameters in the 6 models were eta=0.00106; gamma=4.3; max_depth=2; min_child_weight=7; subsample=0.36; colsample_bytree=0.613; nrounds=1,772.

### External validation of the models in a separate population cohort

To further validate our models and results, we leveraged the data from a more recent population cohort in Finland, FINRISK 2007 (see **Table S1**). In this cohort, the choice of participants, sample collection, and related methods for the data used in the current study were similar to FINRISK 2002 to facilitate inter-cohort comparisons, and are reported elsewhere.^30^ The study protocol of FINRISK 2007 was approved by the Coordinating Ethical Committee of the Hospital District of Helsinki and Uusimaa (Ref. 229/EO/2006). All participants have signed an informed consent.

Briefly, compared to FINRISK 2002, FINRISK 2007 features a smaller number of participants who donated fecal samples (*N* = 258 after excluding pregnant individuals or antibiotic use in the last 6 months), they were younger on average, and a smaller proportion of them were in the high FLI group. To produce data for the validation, methods and quality control related to DNA extraction, sequencing, taxonomic assignments, and calculation of FLI values were identical to FINRISK 2002 data, as described above. For the phylogenetic transform (performed otherwise identically), only taxa passing the filtering in FINRISK 2002 bacterial data set were retained in FINRISK 2007 and a pseudo-count of 1 was used for taxa unobserved in the new data, to exactly match the node balance names. The FINRISK 2007 data was then subset to the model features of the main classification models (sex, age, and the 11 bacterial balances), and input in each of the 6 LOGOCV classification models. The results of these predictions were then compared against the true FLI groups (FLI ≥ 60 and FLI < 60) of the participants (**Table S5**). Receiver operating characteristic and precision-recall curves for the external validation were calculated similarly to the main models for the AUC and AUPRC metrics and plotted after averaging the predictions of the 6 models to obtain single curves (**Figure S6**).

### Identification of predictive features specific to liver function

Because the FLI also incorporates BMI and waist circumference, and they strongly contribute to the index,^26^ we deemed it necessary to further investigate which of the identified balances were specific to liver function. The participants were first grouped by age (< 40, 40 – 60, and 60 <), sex (female or male) and BMI (< 25, 25 – 30, and 30 <) into 18 categories (*N* = 105 ∼ 711 per category). We performed feature selection similarly to the FLI models by fitting random forest regressors for GGT and triglycerides with mlr 2.16.0.^92^ This was done separately in each of the 18 categories, and in each category, we again used LOGOCV with the regions to obtain 6 runs per category. Finally, the features predictive of GGT or triglycerides in each category were selected as the intersect of top 50 features in the 6 LOGOCV iterations by permutation importance. The intersect of features predictive of GGT or triglycerides in any of the categories and the features predictive of categorical FLI were identified as specific to liver function.

### Pathway inference for taxa associated with FLI

Our taxonomic matching of the reads is based on the genomes of GTDB (release 89),^28^ which are all complete or nearly complete and available in online databases. This enables us to estimate the likely genetic content, and thus, the metabolic potential of the microbes associated with FLI. We use this approach because the sequencing depth of our samples does not allow assembling contigs and (metagenome-assembled) genomes, required for pathway predictions. Because of the compositional phylogenetic transform, among other features of our data, previously developed approaches such as PICRUSt,^97^ could not be used here.

The genomes of all 336 bacteria under at least one of the predictive balances were downloaded from NCBI. 119 of these genomes were originally not annotated, which is a requirement for pathway prediction. Therefore, Prokka v1.14.6,^98^ was used to annotate the 119 unannotated genomes as a preliminary step. Pathway predictions were then performed for all 336 genomes with mpwt v0.5.3 multiprocessing tool,^99^ for the PathoLogic pipeline of Pathway Tools 23.0.^100^ Pathways for ethanol and short chain fatty acid (acetate, butyrate, propionate) production, bile acid metabolism, and choline degradation to trimethylamine were identified from MetaCyc pathway classifications (see ref. ^101^, and **Table S4**). The prevalence of these processes was then assessed in the analyzed genomes and summarized per process to consider the possible links of the taxa with fatty liver pathophysiology. Finally, the presence of individual pathways for acetate and ethanol production was also outlined for each genome.

## Data Availability

The datasets generated during and analyzed during the current study are not public, but are available based on a written application to the THL Biobank as instructed in: https://thl.fi/en/web/thl-biobank/for-researchers

## Data availability statement

The analysis code written for this study is included with the Supplementary Information. The datasets generated during and analyzed during the current study are not public, but are available based on a written application to the THL Biobank as instructed in: https://thl.fi/en/web/thl-biobank/for-researchers

## Authors’ contributions

M.R., F.Å., V.M., V.S., R.K, L.L and T.N designed the work. A.H., L.V., G.M., P.J., V.S., M.J and R.K. acquired the data. M.R., L.L. and T.N. analyzed the data. M.R. wrote the manuscript in consultation with all authors. M.I., P.J., V.S., R.K., L.L. and T.N. supervised the work. All authors gave final approval of the version to be published.

## Acknowledgements

We thank all participants of the FINRISK 2002 and FINRISK 2007 surveys for their contributions to this work, and Tara Schwartz for assistance with laboratory work. We also thank the editor and both anonymous reviewers for their constructive criticism.

