## Supplementary material for "Links between gut microbiome composition and fatty liver disease in a large population sample": Analysis_scripts_v1.3.R

```
#Analysis scripts for the manuscript: "Links between gut microbiome composition and fatty liver disease in a large population sample" by Ruuskanen et al.
#The datasets generated during and analyzed during the current study are not public, but are available based on a written application to the THL Biobank as instructed in the web-site of the biobank: https://thl.fi/en/web/thl-biobank/for-researchers 

#Required packages can be loaded with this simple script. Additional packages should be added to the "packages" variable from which they are automatically installed and loaded. Plotting theme is set to a minimalistic black and white for ggplot.

if (!requireNamespace("BiocManager")) {
  install.packages("BiocManager")
}

#devtools::install_github("clauswilke/ggtext")
#devtools::install_github("YuLab-SMU/treeio")

packages <- c("ggplot2", "mlr", "svglite", "ggthemes", "phyloseq", "vegan", "uwot", "dbscan", "mefa", "caret", "xgboost", "ape", "philr", "pdp", "mlrMBO", "ROCR", "pROC", "precrec", "treeio", "ggtree", "ggnewscale", "patchwork", "gtools", "ALDEx2", "BiocParallel", "microbiome", "dplyr")


is.installed <- function(pkg) {
  new.pkg <- pkg[!(pkg %in% installed.packages()[, "Package"])]
  if (length(new.pkg)) {
    BiocManager::install(new.pkg, ask=F)
  }
  sapply(pkg, require, character.only = TRUE)
}
is.installed(packages)


theme_set(theme_tufte(base_family = "sans", base_size = 18) + theme(panel.border = element_rect(colour = "black", fill = NA), axis.text = element_text(colour = "black", size = 18)))


#Phyloseq objects are constructed based on output from the Centrifuge runs against GTDB. This information will be highly useful later on for subsetting and pooling data on different taxonomic levels, and in the use of phylogenetically informed compositional transformation (PhILR).
#load the up-to-date phenotype data (objects FR02 and FR02names). Contact person: Aki Havulinna, aki.havulinna(at)helsinki.fi
load("FR_02_phenotype_data.RData")
#Subset to data which includes the fecal samples
FR02 <- FR02[!is.na(FR02$Barcode),]
row.names(FR02) <- FR02$Barcode
#load the microbiome data (objects gtdb_bac_data and gtdb_arch_data) and make them if they do not exist
if (file.exists("Liver_analysis_antibiotics/gtdb_bac_data.RDs") && file.exists("Liver_analysis_antibiotics/gtdb_arch_data.RDs")) {
  gtdb_bac_data <- readRDS("Liver_analysis_antibiotics/gtdb_bac_data.RDs")
  gtdb_arch_data <- readRDS("Liver_analysis_antibiotics/gtdb_arch_data.RDs")
} else {
  #Construct the primary phyloseq object and subset to FR02 samples.
  gtdb_data <- import_biom("Liver_analysis_antibiotics/table.biom", parallel=TRUE) #BIOM table was constructed from Centrifuge reports.
  samples_to_keep <- sample_names(gtdb_data)[which(grepl("-", sample_names(gtdb_data)))]
  samples_to_keep <- samples_to_keep[which(grepl("^8", samples_to_keep) | nchar(samples_to_keep) < 11)]
  gtdb_data <- prune_samples(samples_to_keep, gtdb_data)
  #Format the tax table.
  colnames(tax_table(gtdb_data)) <- c("Domain", "Phylum", "Class", "Order", "Family",  "Genus", "Species")
  #Format taxa names to the last defined rank to make them match the GTDB tree.
  gtdb_names <- as.vector(tax_table(gtdb_data)[,7])
  gtdb_names[which(gtdb_names != "s__")] <- paste0("s__", gsub("g__", "", as.vector(tax_table(gtdb_data)[which(gtdb_names != "s__"),6])), "_", gsub("s__", "", as.vector(tax_table(gtdb_data)[which(gtdb_names != "s__"),7])))
  gtdb_names[which(gtdb_names == "s__")] <- as.vector(tax_table(gtdb_data)[which(gtdb_names == "s__"),6])
  gtdb_names[which(gtdb_names == "g__")] <- as.vector(tax_table(gtdb_data)[which(gtdb_names == "g__"),5])
  gtdb_names[which(gtdb_names == "f__")] <- as.vector(tax_table(gtdb_data)[which(gtdb_names == "f__"),4])
  taxa_names(gtdb_data) <- gtdb_names
  #Also format species names in the tax table to match the new taxa names (to get the full species names)
  tax_table(gtdb_data)[,"Species"] <- taxa_names(gtdb_data)
  #Separate controls from samples in the sample data table
  sample_data(gtdb_data) <- data.frame(row.names=sample_names(gtdb_data), Name=sample_names(gtdb_data), Type=ifelse(nchar(sample_names(gtdb_data)) < 11, "Control", "Sample"))
  #Combine with gtdb bacterial and archaeal trees so that only species-level data is preserved. These files can be found in the GTDB repository: https://data.ace.uq.edu.au/public/gtdb/data/releases/release89/89.0/
  gtdb_bac_tree <- read_tree("Liver_analysis_antibiotics/bac120_r89.sp_labels_underscore.tree")
  gtdb_bac_data <- merge_phyloseq(gtdb_data, gtdb_bac_tree)
  gtdb_arch_tree <- read_tree("Liver_analysis_antibiotics/ar122_r89.sp_labels_underscore.tree")
  gtdb_arch_data <- merge_phyloseq(gtdb_data, gtdb_arch_tree)
  #Subset control data into separate objects
  gtdb_bac_ctrl <- subset_samples(gtdb_bac_data, Type=="Control")
  gtdb_arch_ctrl <- subset_samples(gtdb_arch_data, Type=="Control")
  #remove samples with less than 50k reads (total)
  to_be_pruned <- sample_sums(gtdb_bac_data) + sample_sums(gtdb_arch_data) > 50000
  gtdb_bac_data <- prune_samples(to_be_pruned, gtdb_bac_data)
  gtdb_arch_data <- prune_samples(to_be_pruned, gtdb_arch_data)
  #Combine the phenotype data with the bacterial and archaeal data (subsets the objects at the same time to samples)
  gtdb_bac_data <- phyloseq(otu_table(gtdb_bac_data), phy_tree(gtdb_bac_data), tax_table(gtdb_bac_data), sample_data(FR02))
  gtdb_arch_data <- phyloseq(otu_table(gtdb_arch_data), phy_tree(gtdb_arch_data), tax_table(gtdb_arch_data), sample_data(FR02))
  #remove pregnant (GRAVID==2) participants
  gtdb_bac_data <- subset_samples(gtdb_bac_data, GRAVID %in% c(1, NA))
  gtdb_arch_data <- subset_samples(gtdb_arch_data, GRAVID %in% c(1, NA))
  #remove participants who have used antibiotics in the last 6 months (BL_USE_RX_J01==1)
  gtdb_bac_data <- subset_samples(gtdb_bac_data, BL_USE_RX_J01 %in% c(0, NA))
  gtdb_arch_data <- subset_samples(gtdb_arch_data, BL_USE_RX_J01 %in% c(0, NA))
  #save the final objects
  saveRDS(gtdb_bac_data, "Liver_analysis_antibiotics/gtdb_bac_data.RDs")
  saveRDS(gtdb_arch_data, "Liver_analysis_antibiotics/gtdb_arch_data.RDs")
  saveRDS(gtdb_bac_ctrl, "Liver_analysis_antibiotics/gtdb_bac_ctrl.RDs")
  saveRDS(gtdb_arch_ctrl, "Liver_analysis_antibiotics/gtdb_arch_ctrl.RDs")
}

#Define functions for PhILR transforms, accessors, and calculation of Fatty Liver Index (FLI).
#Define functions to transform the taxa counts of a phyloseq object to PhILR balances. Adapted from: https://bioconductor.org/packages/release/bioc/vignettes/philr/inst/doc/philr-intro.html
philr_prepare <- function (phyloseq_obj) {
  #Filter taxa not seen with more than 3 counts in at least 1% of samples and, subsequently, those with a coefficient of variation = 3.
  new_obj <- filter_taxa(phyloseq_obj, function(x) sum(x > 3) > (0.01*length(x)), TRUE)
  new_obj <- filter_taxa(new_obj, function(x) sd(x)/mean(x) > 3.0, TRUE)
  #Add a pseudocount of 1 to the remaining taxa to avoid calculating log-rations involving zeros
  new_obj <- transform_sample_counts(new_obj, function(x) x+1)
  #Process the phylogenetic tree
  #Check that the tree is rooted and all multichotomies are resolved (the tree is binary)
  if (!is.rooted(phy_tree(new_obj))) {
    stop("Error: The tree is not rooted. Please root the tree in your phyloseq object.")
  } else if (!is.binary.tree(phy_tree(new_obj))) {
    stop("Error: There are unresolved multichotomies in the tree.")
  }
  #Rename the internal nodes so they are easier to work with.
  phy_tree(new_obj) <- makeNodeLabel(phy_tree(new_obj), method="number", prefix="n")
  #Check the root name balance of the tree and print it out.
  print(paste0("The root name balance of the tree is: ", name.balance(phy_tree(new_obj), tax_table(new_obj), "n1"), collapse=""))
  return(new_obj)
}
philr_transform <- function (phyloseq_obj) {
  #Check the phyloseq object
  if (!is.rooted(phy_tree(phyloseq_obj))) {
    stop("Error: The tree is not rooted. Please root the tree in your phyloseq object.")
  } else if (!is.binary.tree(phy_tree(phyloseq_obj))) {
    stop("Error: There are unresolved multichotomies in the tree.")
  } else if (!any(grepl("^n[0-9]+", phy_tree(phyloseq_obj)$node.label))) {
    stop("Error: Node labels have not been renamed. Please run philr_prepare() first.")
  }
  #OTU table needs to be transposed for PhILR
  t_otu_table <- t(otu_table(phyloseq_obj))
  #Perform the PhILR transform
  new_obj_philr <- philr(t_otu_table, phy_tree(phyloseq_obj), part.weights="enorm.x.gm.counts", ilr.weights="blw.sqrt")
  return(new_obj_philr)
}

calculate_FLI <- function (phyloseq_sample_data) {
  phyloseq_sample_data <- as.data.frame(phyloseq_sample_data)
  #convert triglycerides from mmolL-1 to mgdL-1
  triglycerides_mgdL <- phyloseq_sample_data$TRIG*88.57
  BMI <- phyloseq_sample_data$BMI
  GGT <- phyloseq_sample_data$GGT
  waist_circumference <- phyloseq_sample_data$VYOTARO
  FLI <- (exp(1)^(0.953*log(triglycerides_mgdL) + 0.139*BMI + 0.718*log(GGT) + 0.053*waist_circumference - 15.745)/
            (1 + exp(1)^(0.953*log(triglycerides_mgdL) + 0.139*BMI + 0.718*log(GGT) + 0.053*waist_circumference - 15.745))) * 100
  FLI <- round(FLI, 3)
  return(FLI)
}

#function for getting tip names from nodes
get.ud.tips <- function(tr,coord){
  l.tips <- list()
  child <- phangorn::Children(tr, name.to.nn(tr,coord))
  if (length(child) < 2) stop(paste0(coord,' is a tip'))
  if (length(child) > 2) stop("Tree is not soley binary.") #TODO: Bit of validation - consider better location
  l.tips[['up']] <- sapply(unlist(phangorn::Descendants(tr,child[1],type='tips')), function(x) nn.to.name(tr, x))
  l.tips[['down']] <- sapply(unlist(phangorn::Descendants(tr,child[2],type='tips')), function(x) nn.to.name(tr, x))
  return(l.tips)
}

#Calculate ordinations.
if (file.exists("Liver_analysis_antibiotics/gtdb_bac_pcoa.RDs") && file.exists("Liver_analysis_antibiotics/gtdb_arch_pcoa.RDs")) {
  gtdb_bac_data_tr <- readRDS("Liver_analysis_antibiotics/gtdb_bac_data_tr.RDs")
  gtdb_bac_philr <- readRDS("Liver_analysis_antibiotics/gtdb_bac_philr.RDs")
  gtdb_bac_dist <- readRDS("Liver_analysis_antibiotics/gtdb_bac_dist.RDs")
  gtdb_bac_pcoa <- readRDS("Liver_analysis_antibiotics/gtdb_bac_pcoa.RDs")
  gtdb_bac_pca <- readRDS("Liver_analysis_antibiotics/gtdb_bac_pca.RDs")
  gtdb_arch_data_tr <- readRDS("Liver_analysis_antibiotics/gtdb_arch_data_tr.RDs")
  gtdb_arch_philr <- readRDS("Liver_analysis_antibiotics/gtdb_arch_philr.RDs")
  gtdb_arch_dist <- readRDS("Liver_analysis_antibiotics/gtdb_arch_dist.RDs")
  gtdb_arch_pcoa <- readRDS("Liver_analysis_antibiotics/gtdb_arch_pcoa.RDs")
  gtdb_arch_pca <- readRDS("Liver_analysis_antibiotics/gtdb_arch_pca.RDs")
} else {
  gtdb_bac_data_tr <- philr_prepare(gtdb_bac_data)
  gtdb_bac_philr <- philr_transform(gtdb_bac_data_tr)
  gtdb_bac_dist <- dist(gtdb_bac_philr, method="euclidean")
  gtdb_bac_pcoa <- ordinate(gtdb_bac_data_tr, 'PCoA', distance=gtdb_bac_dist)
  gtdb_bac_pca <- rda(gtdb_bac_philr)
  saveRDS(gtdb_bac_data_tr, "Liver_analysis_antibiotics/gtdb_bac_data_tr.RDs")
  saveRDS(gtdb_bac_philr, "Liver_analysis_antibiotics/gtdb_bac_philr.RDs")
  saveRDS(gtdb_bac_dist, "Liver_analysis_antibiotics/gtdb_bac_dist.RDs")
  saveRDS(gtdb_bac_pcoa, "Liver_analysis_antibiotics/gtdb_bac_pcoa.RDs")
  saveRDS(gtdb_bac_pca, "Liver_analysis_antibiotics/gtdb_bac_pca.RDs")
  gtdb_arch_data_tr <- philr_prepare(gtdb_arch_data)
  gtdb_arch_philr <- philr_transform(gtdb_arch_data_tr)
  gtdb_arch_dist <- dist(gtdb_arch_philr, method="euclidean")
  gtdb_arch_pcoa <- ordinate(gtdb_arch_data_tr, 'PCoA', distance=gtdb_arch_dist)
  gtdb_arch_pca <- rda(gtdb_arch_philr)
  saveRDS(gtdb_arch_data_tr, "Liver_analysis_antibiotics/gtdb_arch_data_tr.RDs")
  saveRDS(gtdb_arch_philr, "Liver_analysis_antibiotics/gtdb_arch_philr.RDs")
  saveRDS(gtdb_arch_dist, "Liver_analysis_antibiotics/gtdb_arch_dist.RDs")
  saveRDS(gtdb_arch_pcoa, "Liver_analysis_antibiotics/gtdb_arch_pcoa.RDs")
  saveRDS(gtdb_arch_pca, "Liver_analysis_antibiotics/gtdb_arch_pca.RDs")
}

#Calculate the Fatty Liver Index.
sample_data(gtdb_arch_data_tr)$FLI <- calculate_FLI(sample_data(gtdb_arch_data_tr))
sample_data(gtdb_bac_data_tr)$FLI <- calculate_FLI(sample_data(gtdb_bac_data_tr))

#Plot FLI and its components.
p_TRIG <- ggplot(data=sample_data(gtdb_bac_data_tr), aes(x=TRIG)) + geom_histogram(bins=20, fill="gray50") + 
  scale_x_log10(breaks = scales::trans_breaks("log10", function(x) 10^x), labels = scales::trans_format("log10", scales::math_format(10^.x))) +
  scale_y_continuous(limits=c(0,1500)) + 
  xlab("Triglycerides (mmol/L)") + ylab("Count") + theme(axis.text.x = element_text(size=14)) + ggtitle("B")
p_BMI <- ggplot(data=sample_data(gtdb_bac_data_tr), aes(x=BMI)) + geom_histogram(bins=20, fill="gray50") +
  scale_y_continuous(limits=c(0,1500)) +
  xlab("BMI") + theme(axis.title.y = element_blank(), axis.text.y = element_blank(), axis.ticks.y = element_blank(), axis.text.x = element_text(size=14))
p_GGT <- ggplot(data=sample_data(gtdb_bac_data_tr), aes(x=GGT)) + geom_histogram(bins=20, fill="gray50") +
  scale_x_log10(breaks = scales::trans_breaks("log10", function(x) 10^x), labels = scales::trans_format("log10", scales::math_format(10^.x))) +
  scale_y_continuous(limits=c(0,1500)) +
  xlab("GGT (units/L)") + theme(axis.title.y = element_blank(), axis.text.y = element_blank(), axis.ticks.y = element_blank(), axis.text.x = element_text(size=14))
p_WAIST <- ggplot(data=sample_data(gtdb_bac_data_tr), aes(x=VYOTARO)) + geom_histogram(bins=20, fill="gray50") +
  scale_y_continuous(limits=c(0,1500)) + 
  xlab("Waist Circumference (cm)") + theme(axis.title.y = element_blank(), axis.text.y = element_blank(), axis.ticks.y = element_blank(), axis.text.x = element_text(size=14))
plot_FLI <- data.frame(FLI = sample_data(gtdb_bac_data_tr)$FLI)
plot_FLI_label <- paste0("FLI = 60\n", round(length(which(plot_FLI$FLI >= 60))/length(plot_FLI$FLI)*100, 1), "%")
p_FLI <- ggplot(data=plot_FLI, aes(x=FLI)) + geom_histogram(breaks=seq(0,100, by=5), fill="gray50") + geom_vline(aes(xintercept = 60), linetype = "dashed", size = 0.5) + 
  annotate("text", x = 80, y = 500, label = plot_FLI_label, size = 5) + xlab("Fatty Liver Index") + ylab("Count") + scale_x_continuous(breaks=c(-Inf, 0, 20, 40, 60, 80, 100, Inf)) +
  theme(axis.text.x = element_text(size=14)) + ggtitle("A")

#Plot beta-diversity (PCoA)
#gtdb_arch_beta_div <- plot_ordination(gtdb_arch_data_tr, gtdb_arch_pcoa, type="samples", justDF=TRUE)
#gtdb_bac_beta_div <- plot_ordination(gtdb_bac_data_tr, gtdb_bac_pcoa, type="samples", justDF=TRUE)
#ggplot(data=gtdb_bac_beta_div, aes(x=Axis.1, y=Axis.2)) + geom_point(aes(colour = FLI), size=0.25) + scale_colour_gradient(low="blue", high="orange") + labs(x=paste0("PCoA 1 (", round(gtdb_bac_pcoa$values[1,2]*100), "%)", collapse=""), y=paste0("PCoA 2 (", round(gtdb_bac_pcoa$values[2,2]*100), "%)", collapse=""), title="Overlay of Fatty Liver Index\non bacterial beta-diversity")
#ggplot(data=gtdb_arch_beta_div, aes(x=Axis.1, y=Axis.2)) + geom_point(aes(colour = FLI), size=0.25) + scale_colour_gradient(low="blue", high="orange") + labs(x=paste0("PCoA 1 (", round(gtdb_arch_pcoa$values[1,2]*100), "%)", collapse=""), y=paste0("PCoA 2 (", round(gtdb_arch_pcoa$values[2,2]*100), "%)", collapse=""), title="Overlay of Fatty Liver Index\non archaeal beta-diversity")

#Check if bacterial or archaeal PC axes are linearly correlated with FLI. Plot the correlated axes. 
PCA_lm_bac_data <- cbind(summary(gtdb_bac_pca)$sites, sample_data(gtdb_bac_data_tr))
colnames(PCA_lm_bac_data)[1:3] <- c("PC1_b", "PC2_b", "PC3_b") 
PCA_lm_bac_perc <- head(round(100*gtdb_bac_pca$CA$eig/sum(gtdb_bac_pca$CA$eig),2), 10)
PCA_lm_arch_data <- cbind(summary(gtdb_arch_pca)$sites, sample_data(gtdb_arch_data_tr))
colnames(PCA_lm_arch_data)[1:3] <- c("PC1_a", "PC2_a", "PC3_a") 
PCA_lm_arch_perc <- head(round(100*gtdb_arch_pca$CA$eig/sum(gtdb_arch_pca$CA$eig),2), 10)
PCA_lm_data <- cbind(PCA_lm_bac_data, PCA_lm_arch_data[,1:3])
FLI_lm_model <- lm(log10(FLI) ~ PC1_b + PC2_b + PC3_b + MEN + BL_AGE + ALKI2_FR02, data=PCA_lm_data)
summary(FLI_lm_model)
#plot(residuals(FLI_lm_model))
FLI_arrow <- data.frame(t(matrix(as.numeric(coef(FLI_lm_model)[c("PC1_b", "PC3_b")]))))
PCA_lm_data$FLI_cat <- ifelse(PCA_lm_data$FLI >= 60, "FLI = 60", "FLI < 60")
p <- ggplot(data=PCA_lm_data, aes(x=PC1_b, y=PC3_b)) + geom_point(aes(colour = FLI_cat), size=0.4) + #geom_segment(aes(x=0, y=0, xend=FLI_arrow$X1, yend=FLI_arrow$X2), arrow=arrow(length=unit(0.2, "cm"))) +
  scale_colour_manual(values=c("slateblue1", "red"), name="FLI category", limits=c( "FLI < 60", "FLI = 60")) + labs(x=paste0("PC1 (", PCA_lm_bac_perc[1], "%)", collapse=""), y=paste0("PC3 (", PCA_lm_bac_perc[3], "%)", collapse=""))
ggsave(file = "Liver_analysis_antibiotics/FLI_PC_axes.svg", plot=p, units="cm", width=18, height=12)
plot_lm_data <- reshape2::melt(PCA_lm_data[,c("FLI", "PC1_b", "PC2_b", "PC3_b")], measure.vars=c("PC1_b", "PC2_b", "PC3_b"))
plot_lm_data$FLI_groups <- cut(plot_lm_data$FLI, c(-Inf, 20, 40, 60, 80, Inf), labels=c("0-20", "20-40", "40-60", "60-80", "80-100"))
plot_lm_data <- plot_lm_data[which(complete.cases(plot_lm_data)),]
plot_lm_data2 <- data.frame(PCA_lm_data["FLI"], lapply(PCA_lm_data[c("PC1_b", "PC2_b", "PC3_b")], function(x) quantcut(x=x, q=4, na.rm=T, labels=c("0-.25", ".25-.5", ".5-.75", ".75-1")))) 
plot_lm_data2 <- reshape2::melt(plot_lm_data2, measure.vars=c("PC1_b", "PC2_b", "PC3_b"))
plot_lm_data2$value <- factor(plot_lm_data2$value, levels=c("0-.25", ".25-.5", ".5-.75", ".75-1"))
PC_labs <- c("Bacterial PC1", "Bacterial PC2", "Bacterial PC3")
names(PC_labs) <- c("PC1_b", "PC2_b", "PC3_b")
p_PC_boxplots <- ggplot(data=plot_lm_data2, aes(value, FLI)) +
  geom_boxplot() +
  geom_jitter(alpha=0.1, size=0.5) +
  geom_hline(yintercept=60, alpha=0.75, linetype="dashed") +
  facet_wrap(~variable, labeller=labeller(variable=PC_labs)) +
  ylab("Fatty Liver Index") +
  xlab("PC quantiles") +
  theme(axis.text.x = element_text(size=14), strip.text.x = element_text(size=20)) +
  ggtitle("C")
p <- (p_FLI | p_TRIG | p_BMI | p_GGT | p_WAIST) / p_PC_boxplots
ggsave(file = "Liver_analysis_antibiotics/FLI_boxplots_Fig1_alt.svg", plot=p, units="cm", width=42, height=24)

#Plot the loadings of the top 3 bacterial PCs.
gtdb_bac_pca_top_scores <- data.frame(scores(gtdb_bac_pca, choices=c(1:3), display="species"))
gtdb_bac_PC1_top_scores <- head(gtdb_bac_pca_top_scores[order(-abs(gtdb_bac_pca_top_scores$PC1)),], 20)
gtdb_bac_PC1_top_scores$balance <- paste0(unlist(lapply(rownames(gtdb_bac_PC1_top_scores), function (x, y) name.balance(phy_tree(y), tax_table(y), x), y=gtdb_bac_data_tr)), ": ", rownames(gtdb_bac_PC1_top_scores))
#Flip the data to have the balances in the correct direction in the loading plot (since the PC1~FLI connection is positive)
gtdb_bac_PC1_top_scores$PC_value <- -gtdb_bac_PC1_top_scores$PC1
gtdb_bac_PC1_top_scores$PC <- "PC1"
gtdb_bac_PC2_top_scores <- head(gtdb_bac_pca_top_scores[order(-abs(gtdb_bac_pca_top_scores$PC2)),], 20)
gtdb_bac_PC2_top_scores$balance <- paste0(unlist(lapply(rownames(gtdb_bac_PC2_top_scores), function (x, y) name.balance(phy_tree(y), tax_table(y), x), y=gtdb_bac_data_tr)), ": ", rownames(gtdb_bac_PC2_top_scores))
#Flip the data to have the balances in the correct direction in the loading plot (since the PC2~FLI connection is positive)
gtdb_bac_PC2_top_scores$PC_value <- -gtdb_bac_PC2_top_scores$PC2
gtdb_bac_PC2_top_scores$PC <- "PC2"
gtdb_bac_PC3_top_scores <- head(gtdb_bac_pca_top_scores[order(-abs(gtdb_bac_pca_top_scores$PC3)),], 20)
gtdb_bac_PC3_top_scores$balance <- paste0(unlist(lapply(rownames(gtdb_bac_PC3_top_scores), function (x, y) name.balance(phy_tree(y), tax_table(y), x), y=gtdb_bac_data_tr)), ": ", rownames(gtdb_bac_PC3_top_scores))
gtdb_bac_PC3_top_scores$PC_value <- gtdb_bac_PC3_top_scores$PC3
gtdb_bac_PC3_top_scores$PC <- "PC3"
PC_loadings_df <- as.data.frame(mapply(c, gtdb_bac_PC1_top_scores[c("PC_value","balance", "PC")], gtdb_bac_PC2_top_scores[c("PC_value","balance", "PC")], gtdb_bac_PC3_top_scores[c("PC_value","balance", "PC")]))
PC_loadings_df$PC_value <- as.numeric(as.character(PC_loadings_df$PC_value))

PC_loadings_df$balance <- gsub("[A-Z]{1}[a-z]+_([a-z]__.*)", "\\1" , PC_loadings_df$balance)
PC_loadings_df$balance <- gsub("[A-Z]{1}[a-z]+_([a-z]__.*)", "\\1" , PC_loadings_df$balance)
PC_loadings_df$balance <- gsub("k__", "k__Bacteria", PC_loadings_df$balance)
PC_loadings_df$balance <- ifelse(PC_loadings_df$PC_value > 0, paste0(gsub(".*/(.*):.*", "\\1", PC_loadings_df$balance), " / ", gsub("(.*)/.*", "\\1", PC_loadings_df$balance), ": ", gsub(".*: (.*)", "\\1", PC_loadings_df$balance)), gsub("/", " / ", PC_loadings_df$balance))
PC_loadings_df$PC_value <- abs(PC_loadings_df$PC_value)
PC1_loadings_df <- PC_loadings_df[PC_loadings_df$PC == "PC1",]
PC1_loadings_df$balance <- factor(PC1_loadings_df$balance, levels=rev(PC1_loadings_df$balance))
PC2_loadings_df <- PC_loadings_df[PC_loadings_df$PC == "PC2",]
PC2_loadings_df$balance <- factor(PC2_loadings_df$balance, levels=rev(PC2_loadings_df$balance))
PC3_loadings_df <- PC_loadings_df[PC_loadings_df$PC == "PC3",]
PC3_loadings_df$balance <- factor(PC3_loadings_df$balance, levels=rev(PC3_loadings_df$balance))
#In the figure, negative loadings denote a positive connection between the clade in the numerator (on the left) and FLI (through the primary component)
p1 <- ggplot(data=PC1_loadings_df, aes(x=balance, y=PC_value)) + geom_bar(stat="identity", width=0.5, color="black", fill="gray") + coord_flip() + scale_y_continuous(limits=c(0,10)) +
  theme(axis.title.y=element_blank(), axis.text=element_text(size=10), axis.title.x=element_text(size=12)) + ylab("Loading for PC1 (13.52%)")
p2 <- ggplot(data=PC2_loadings_df, aes(x=balance, y=PC_value)) + geom_bar(stat="identity", width=0.5, color="black", fill="gray") + coord_flip() + scale_y_continuous(limits=c(0,10)) +
  theme(axis.title.y=element_blank(), axis.text=element_text(size=10), axis.title.x=element_text(size=12)) + ylab("Loading for PC2 (7.09%)")
p3 <- ggplot(data=PC3_loadings_df, aes(x=balance, y=PC_value)) + geom_bar(stat="identity", width=0.5, color="black", fill="gray") + coord_flip() + scale_y_continuous(limits=c(0,10)) +
  theme(axis.title.y=element_blank(), axis.text=element_text(size=10), axis.title.x=element_text(size=12)) + ylab("Loading for PC3 (6.13%)")
p <- p1 / p2 / p3
ggsave(file = "Liver_analysis_antibiotics/FLI_PC_loadings.svg", plot=p, units="cm", width=20, height=30)

#Next stage of the analysis is predicting FLI based on the composition of the microbial community. Using xgboost to model FLI with PhILR balances and covariates enables the identification of likely key players in fatty liver disease.
#Collect the data into one object by concatenating FLI, covariates, and the bacterial and archaeal PhILR balances.
FLI_ML_data <- cbind(data.frame(sample_data(gtdb_bac_data_tr)[,c("FLI", "MEN", "BL_AGE", "ALKI2_FR02")]), as(otu_table(gtdb_bac_philr, taxa_are_rows = FALSE), "matrix"))
FLI_ML_arch_data <- as(otu_table(gtdb_arch_philr, taxa_are_rows = FALSE), "matrix")
colnames(FLI_ML_arch_data) <- paste0(colnames(FLI_ML_arch_data), "_a")
FLI_ML_data <- cbind(FLI_ML_data, FLI_ML_arch_data)
#Replace zeros
FLI_ML_data$FLI[is.na(FLI_ML_data$FLI)] <- mean(FLI_ML_data$FLI, na.rm = TRUE)
FLI_ML_data$ALKI2_FR02[is.na(FLI_ML_data$ALKI2_FR02)] <- mean(FLI_ML_data$ALKI2_FR02, na.rm = TRUE)
#Divide the data first to 70% train 30% test preserving sex and area class balance
part_df <- data.frame(sample_data(gtdb_bac_data_tr))
part_criteria <- paste0(part_df$MEN, part_df$ALUE)
set.seed(42)
trainIndex <- createDataPartition(part_criteria, p = .7, list = FALSE, times = 1)
FLI_ML_train <- FLI_ML_data[trainIndex,]
FLI_ML_test <- FLI_ML_data[-trainIndex,]
train_groups <- data.frame(sample_data(gtdb_bac_data_tr))$ALUE[trainIndex]
cv_folds <- groupKFold(train_groups, k = length(unique(train_groups)))
train_data <- as.matrix(FLI_ML_train)

#Perform feature selection and hyperparameter optimization of the main models.
#Find the optimal predictors by random forest permutation importance in mlr and hyperparameters through Bayesian optimization (adapted from https://www.simoncoulombe.com/2019/01/bayesian/)
obj.fun <- smoof::makeSingleObjectiveFunction(
  name = "xgb_cv_bayes",
  fn = function(x) {
    train_mbo <- dtrain[cv_folds[[as.numeric(x["fold"])]],]
    test_idx <- setdiff(c(1:dim(dtrain)[1]), cv_folds[[as.numeric(x["fold"])]])
    test_mbo <- dtrain[test_idx,]
    watchlist <- list(train=train_mbo, test=test_mbo)
    set.seed(42)
    cv <- xgb.train(data = train_mbo,
      watchlist = watchlist,
      params = list(
      booster          = "gbtree",
      eta              = x["eta"],
      max_depth        = x["max_depth"],
      min_child_weight = x["min_child_weight"],
      gamma            = x["gamma"],
      subsample        = x["subsample"],
      colsample_bytree = x["colsample_bytree"],
      objective        = "reg:squarederror", 
      eval_metric      = "rmse"),
      nrounds          = x["nrounds"],
      showsd = TRUE,
      verbose = FALSE)
    
    as.numeric(cv$evaluation_log[max(iter),3])
  },
  par.set = makeParamSet(
    makeNumericParam("eta",              lower = 0.001,  upper = 0.3),
    makeNumericParam("gamma",            lower = 0.1,   upper = 5),
    makeIntegerParam("max_depth",        lower = 2,     upper = 8),
    makeIntegerParam("min_child_weight", lower = 1,     upper = 10),
    makeNumericParam("subsample",        lower = 0.2,   upper = 0.8), #set maximum to 80%
    makeNumericParam("colsample_bytree", lower = 0.2,   upper = 0.9),  #set maximum to 90%
    makeIntegerParam("nrounds",          lower = 50,    upper = 5000),
    makeIntegerParam("fold",             lower = 1,     upper = 6, tunable = FALSE)
  ),
  minimize = TRUE
)

obj.fun_class <- smoof::makeSingleObjectiveFunction(
  name = "xgb_cv_bayes",
  fn = function(x) {
    train_mbo <- dtrain_class[cv_folds[[as.numeric(x["fold"])]],]
    test_idx <- setdiff(c(1:dim(dtrain_class)[1]), cv_folds[[as.numeric(x["fold"])]])
    test_mbo <- dtrain_class[test_idx,]
    watchlist <- list(train=train_mbo, test=test_mbo)
    set.seed(42)
    cv <- xgb.train(data = train_mbo,
      watchlist = watchlist,
      params = list(
      booster          = "gbtree",
      eta              = x["eta"],
      max_depth        = x["max_depth"],
      min_child_weight = x["min_child_weight"],
      gamma            = x["gamma"],
      subsample        = x["subsample"],
      colsample_bytree = x["colsample_bytree"],
      objective        = "binary:logistic", 
      eval_metric      = "auc"),
      nrounds          = x["nrounds"],
      showsd = TRUE,
      verbose = FALSE)
    
    as.numeric(cv$evaluation_log[max(iter),3])
  },
  par.set = getParamSet(obj.fun),
  minimize = FALSE
)
#Generate random sets of the parameter space
set.seed(42)
des <- generateDesign(n=50, par.set = getParamSet(obj.fun), fun = lhs::randomLHS)

#Gather a set of consensus most important features present in all models: intersect of top 50 predictors (ranger permutation importance) in all folds
if (file.exists("Liver_analysis_antibiotics/top_predictors.RDs") && file.exists("Liver_analysis_antibiotics/top_predictors_class.RDs")) {
  top_predictors <- readRDS("Liver_analysis_antibiotics/top_predictors.RDs")
  top_predictors_class <- readRDS("Liver_analysis_antibiotics/top_predictors_class.RDs")
} else {
  top_predictors <- list(NULL)
  top_predictors_class <- list(NULL)
  for (fold in 1:length(cv_folds)) {
	#First do feature selection for regression
	fold_regr_task <- makeRegrTask(data = as.data.frame(train_data[cv_folds[[fold]],]), target = "FLI")
	set.seed(42)
	fv <- generateFilterValuesData(fold_regr_task, method = "ranger_permutation")
	fold_predictors <- data.frame(fv$data[order(-fv$data$value),])[1:50,]
  top_predictors[[fold]] <- fold_predictors$name
  #Then for classification
	class_fold_data <- as.data.frame(train_data[cv_folds[[fold]],])
	class_fold_data$FLI <- as.factor(ifelse(class_fold_data$FLI >= 60, 1, 0))
	fold_class_task <- makeClassifTask(data = class_fold_data, target = "FLI", positive = "1")
	set.seed(42)
	fv_class <- generateFilterValuesData(fold_class_task, method = "ranger_permutation")
	fold_predictors_class <- data.frame(fv_class$data[order(-fv_class$data$value),])[1:50,]
  top_predictors_class[[fold]] <- fold_predictors_class$name
  }
  print(top_predictors)
  saveRDS(top_predictors, "Liver_analysis_antibiotics/top_predictors.RDs")
  print(top_predictors_class)
  saveRDS(top_predictors_class, "Liver_analysis_antibiotics/top_predictors_class.RDs")
}
top_predictors <- Reduce(intersect, top_predictors)
top_predictors_class <- Reduce(intersect, top_predictors_class)

#Subset features in the data based on results of the feature selection
train_data_subset <- as.matrix(FLI_ML_train[,colnames(FLI_ML_train) %in% c("FLI", top_predictors)])
dtrain <- xgb.DMatrix(data = train_data_subset[,-1], label = train_data_subset[,1])
train_data_subset_class <- as.matrix(FLI_ML_train[,colnames(FLI_ML_train) %in% c("FLI", top_predictors_class)])
dtrain_class <- xgb.DMatrix(data = train_data_subset_class[,-1], label = ifelse(train_data_subset_class[,1] >= 60, 1, 0))

#Run hyperparameter optimization with the feature selected models
if (file.exists("Liver_analysis_antibiotics/xgboost_parameters.RDs")) {
  run <- readRDS("Liver_analysis_antibiotics/xgboost_parameters.RDs")
  run_class <- readRDS("Liver_analysis_antibiotics/xgboost_parameters_class.RDs")
} else {
  control <- makeMBOControl()
  control <- setMBOControlTermination(control, iters = 100)
  run <- mbo(fun = obj.fun, design = des[21:50,], control = control, show.info = TRUE)
  saveRDS(run, "Liver_analysis_antibiotics/xgboost_parameters.RDs")
  run_class <- mbo(fun = obj.fun_class, design = des[21:50,], control = control, show.info = TRUE)
  saveRDS(run_class, "Liver_analysis_antibiotics/xgboost_parameters_class.RDs")
}

#Construct the main models for FLI.
#Define regions
test_fold_region_codes <- data.frame(c(2, 7, 3, 6, 4, 5), c("North Karelia", "Lapland", "North Savonia", "Oulu province", "Turku/Loimaa", "Helsinki/Vantaa"))
test_fold_regions <- part_df$ALUE[-trainIndex]
test_data <- as.matrix(FLI_ML_test[,colnames(FLI_ML_test) %in% c("FLI", top_predictors)])
dtest <- xgb.DMatrix(data = test_data[,-1], label = test_data[,1])
#For sensitivity analysis, optionally test covariates and balances separately
train_data_subset_class <- list(as.matrix(FLI_ML_train[,colnames(FLI_ML_train) %in% c("FLI", top_predictors_class)]), as.matrix(FLI_ML_train[,colnames(FLI_ML_train) %in% c("FLI", c("MEN", "BL_AGE"))]), as.matrix(FLI_ML_train[,colnames(FLI_ML_train) %in% c("FLI", top_predictors_class[-which(top_predictors_class %in% c("MEN", "BL_AGE"))])]))
test_data_class <- list(as.matrix(FLI_ML_test[,colnames(FLI_ML_test) %in% c("FLI", top_predictors_class)]), as.matrix(FLI_ML_test[,colnames(FLI_ML_test) %in% c("FLI", c("MEN", "BL_AGE"))]), as.matrix(FLI_ML_test[,colnames(FLI_ML_test) %in% c("FLI", top_predictors_class[-which(top_predictors_class %in% c("MEN", "BL_AGE"))])]))
sens_codes <- c("covariates + balances", "covariates only", "balances only")
validation_stats <- list(NULL)
validation_stats_class <- list(NULL)
validation_predictors <- list(NULL)
validation_predictors_class <- list(NULL)
validation_predictions <- list(NULL)
validation_predictions_class <- list(NULL)
pd_plot_data <- list(NULL)
for (fold in 1:length(cv_folds)) {
    train_fold <- dtrain[cv_folds[[fold]],]
    test_fold <- dtest[which(test_fold_regions == test_fold_region_codes[fold,1]),]
    print(paste0("Regression model omitting region ", unique(train_groups[-cv_folds[[fold]]]), " tested on region ", test_fold_region_codes[fold,1], " (", test_fold_region_codes[fold,2], ")"))
    set.seed(42)
    fold_model <- xgboost(params = run$x,
      objective = "reg:squarederror", 
      eval_metric = "rmse",
      data = train_fold,
      nrounds = run$x$nrounds,
      verbose = 0)
    print(paste0("Regression model trained in fold ", fold))
    predicted <- predict(fold_model, test_fold)
    validation_predictions[[fold]] <- predicted
    y_test <- getinfo(test_fold, "label")
    y_test_mean <- mean(y_test)
    # Calculate total sum of squares
    tss <- sum((y_test - y_test_mean)^2)
    # Calculate residual sum of squares
    residuals <- y_test - predicted
    rss <- sum(residuals^2)
    # Calculate R-squared
    validation_stats[[fold]] <- round(1 - (rss/tss),3)
    xgb.save(fold_model, paste0("Liver_analysis_antibiotics/fold_model_", fold, ".model"))
    validation_predictors[[fold]] <- data.frame(xgb.importance(colnames(test_data[,-1]), model=fold_model))
    validation_stats_sensset <- list(NULL)
    for (sensset in 1:3) {
      dtrain_class <- xgb.DMatrix(data = train_data_subset_class[[sensset]][,-1], label = ifelse(train_data_subset_class[[sensset]][,1] >= 60, 1, 0))
      train_fold_class <- dtrain_class[cv_folds[[fold]],]
      dtest_class <- xgb.DMatrix(data = test_data_class[[sensset]][,-1], label = ifelse(test_data_class[[sensset]][,1] >= 60, 1, 0))
      test_fold_class <- dtest_class[which(test_fold_regions == test_fold_region_codes[fold,1]),]
      print(paste0("Classification model omitting region ", unique(train_groups[-cv_folds[[fold]]]), " tested on region ", test_fold_region_codes[fold,1], " (", test_fold_region_codes[fold,2], "), including ", sens_codes[sensset]))
      set.seed(42)
      fold_model_class <- xgboost(params = run_class$x,
        objective = "binary:logistic", 
        eval_metric = "auc",
        data = train_fold_class,
        nrounds = run_class$x$nrounds,
        verbose = F)
      predicted_class <- predict(fold_model_class, test_fold_class)
      if (sensset==1) {
        validation_predictions_class[[fold]] <- predicted_class
      }
      aucs <- evalmod(scores=predicted_class, labels=getinfo(test_fold_class, "label"))
      auc_values <- c(as.numeric(attr(aucs, "aucs")$aucs), as.numeric(attr(aucs, "data_info")$np/(attr(aucs, "data_info")$np+attr(aucs, "data_info")$nn)))
      validation_stats_sensset[[sensset]] <- auc_values
      print(paste0("Model AUC: ", round(auc_values[1], 4), " and AUPRC: ", round(auc_values[2], 4), " at baseline of ", round(auc_values[3], 2)*100, "% positive cases in ", dim(test_fold_class)[1], " participants, when geograpically extrapolating to ", test_fold_region_codes[fold,2]))
      xgb.save(fold_model_class, paste0("Liver_analysis_antibiotics/fold_model_class", fold, "_", sensset, ".model"))
    }
    validation_stats_class[[fold]] <- as.data.frame(validation_stats_sensset, col.names=c(1:3))
    fold_model_class <- xgb.load(paste0("Liver_analysis_antibiotics/fold_model_class", fold, "_1.model"))
    validation_predictors_class[[fold]] <- data.frame(xgb.importance(colnames(test_data_class[[1]][,-1]), model=fold_model_class))
    #Gather partial dependence of the variables from the model (takes quite long to compute)
    #Retrain the full model (partial doesn't work with loaded xgboost models)
    dtrain_class <- xgb.DMatrix(data = train_data_subset_class[[1]][,-1], label = ifelse(train_data_subset_class[[1]][,1] >= 60, 1, 0))
    train_fold_class <- dtrain_class[cv_folds[[fold]],]
    dtest_class <- xgb.DMatrix(data = test_data_class[[1]][,-1], label = ifelse(test_data_class[[1]][,1] >= 60, 1, 0))
    test_fold_class <- dtest_class[which(test_fold_regions == test_fold_region_codes[fold,1]),]
    set.seed(42)
    fold_model_class <- xgboost(params = run_class$x,
                                objective = "binary:logistic", 
                                eval_metric = "auc",
                                data = train_fold_class,
                                nrounds = run_class$x$nrounds,
                                verbose = F)
    pd_train <- train_data_subset_class[[1]][cv_folds[[fold]], -1]
    pd_data <- list(NULL)
    for (predictor in 1:ncol(pd_train)) {
      pd_df <- partial(fold_model_class, train = pd_train, pred.var = colnames(pd_train)[predictor], plot=FALSE)
      print(paste0("Partial dependence of ", colnames(pd_train)[predictor], " calculated in fold ", fold))
      pd_df$predictor <- colnames(pd_train)[predictor]
      colnames(pd_df)[1] <- "value"
      pd_data[[predictor]] <- pd_df
    }
    pd_data <- do.call(rbind, pd_data)
    pd_data$fold <- fold
    pd_plot_data[[fold]] <- pd_data
}
validation_stats_class_df <- data.frame(CV_region=c(as.character(test_fold_region_codes[,2]), "Mean"), Main_model=round(c(unlist(lapply(validation_stats_class, `[[`, 1))[c(T,F,F)], mean(unlist(lapply(validation_stats_class, `[[`, 1))[c(T,F,F)])),3), Main_model_covariates=round(c(unlist(lapply(validation_stats_class, `[[`, 2))[c(T,F,F)], mean(unlist(lapply(validation_stats_class, `[[`, 2))[c(T,F,F)])),3), Main_model_balances=round(c(unlist(lapply(validation_stats_class, `[[`, 3))[c(T,F,F)], mean(unlist(lapply(validation_stats_class, `[[`, 3))[c(T,F,F)])),3), Main_model_test_set_N=c(as.numeric(table(test_fold_regions)[as.factor(test_fold_region_codes[,1])]), round(mean(table(test_fold_regions)),1)))
validation_stats_class_auprc_df <- data.frame(CV_region=c(as.character(test_fold_region_codes[,2]), "Mean"), Main_model=round(c(unlist(lapply(validation_stats_class, `[[`, 1))[c(F,T,F)], mean(unlist(lapply(validation_stats_class, `[[`, 1))[c(F,T,F)])),3), Main_model_covariates=round(c(unlist(lapply(validation_stats_class, `[[`, 2))[c(F,T,F)], mean(unlist(lapply(validation_stats_class, `[[`, 2))[c(F,T,F)])),3), Main_model_balances=round(c(unlist(lapply(validation_stats_class, `[[`, 3))[c(F,T,F)], mean(unlist(lapply(validation_stats_class, `[[`, 3))[c(F,T,F)])),3), Main_model_test_set_N=c(as.numeric(table(test_fold_regions)[as.factor(test_fold_region_codes[,1])]), round(mean(table(test_fold_regions)),1)), Main_model_test_set_positives=round(c(unlist(lapply(validation_stats_class, `[[`, 1))[c(F,F,T)], mean(unlist(lapply(validation_stats_class, `[[`, 1))[c(F,F,T)])),3))
validation_predictors <- do.call(rbind, validation_predictors)
validation_predictions <- do.call(cbind, validation_predictions)
#validation_predictions_compare <- cbind(getinfo(dtest, "label"), rowMeans(validation_predictions))
validation_importances <- do.call(data.frame, aggregate(. ~ Feature, validation_predictors, function(x) c(mean = mean(x), sd = sd(x))))
validation_importances <- validation_importances[order(-validation_importances$Gain.mean),]
predictor_balances <- list(NULL)
for (balance in validation_importances$Feature) { 
  if (balance %in% c("MEN", "BL_AGE", "ALKI2_FR02")) {
  predictor_balances <- append(predictor_balances, NA)
  } else {
  predictor_balances <- append(predictor_balances, name.balance(phy_tree(gtdb_bac_data_tr), tax_table(gtdb_bac_data_tr), balance))
  }
}
validation_importances$balance <- unlist(predictor_balances)
validation_predictors_class <- do.call(rbind, validation_predictors_class)
validation_predictions_class <- do.call(cbind, validation_predictions_class)
#validation_predictions_class_compare <- cbind(getinfo(dtest_class, "label"), rowMeans(validation_predictions_class))
validation_importances_class <- do.call(data.frame, aggregate(. ~ Feature, validation_predictors_class, function(x) c(mean = mean(x), sd = sd(x))))
validation_importances_class <- validation_importances_class[order(-validation_importances_class$Gain.mean),]
predictor_balances_class <- list(NULL)
for (balance in validation_importances_class$Feature) { 
  if (balance %in% c("MEN", "BL_AGE", "ALKI2_FR02")) {
  predictor_balances_class <- append(predictor_balances_class, NA)
  } else {
  predictor_balances_class <- append(predictor_balances_class, name.balance(phy_tree(gtdb_bac_data_tr), tax_table(gtdb_bac_data_tr), balance))
  }
}
validation_importances_class$balance <- unlist(predictor_balances_class)
pd_plot_data2 <- do.call(rbind, pd_plot_data)
predictor_ranges <- list(NULL)
for (fold in 1:length(cv_folds)) {
  fold_pd_data <- pd_plot_data2[which(pd_plot_data2$fold == fold),]
  fold_predictor_ranges <- aggregate(fold_pd_data["value"], by=list(fold_pd_data$predictor), FUN=range)
  fold_yhat_1_values <- list(NULL)
  for (predictor in 1:length(fold_predictor_ranges$Group.1)){
    min_yhat_1 <- fold_pd_data[which(fold_pd_data$value == fold_predictor_ranges$value[predictor,1] & fold_pd_data$predictor == fold_predictor_ranges$Group.1[predictor]),"yhat"]
    max_yhat_1 <- fold_pd_data[which(fold_pd_data$value == fold_predictor_ranges$value[predictor,2] & fold_pd_data$predictor == fold_predictor_ranges$Group.1[predictor]),"yhat"]
    fold_yhat_1_values[[predictor]] <- data.frame(min_yhat_1 = min_yhat_1, max_yhat_1 = max_yhat_1)
    }
  fold_predictor_ranges <- cbind(fold_predictor_ranges, do.call(rbind, fold_yhat_1_values))
  fold_predictor_ranges$fold <- fold
  predictor_ranges[[fold]] <- fold_predictor_ranges
}
predictor_ranges <- do.call(rbind, predictor_ranges)
predictor_ranges$min_value <- predictor_ranges$value[,1]
predictor_ranges$max_value <- predictor_ranges$value[,2]
median_predictor_ranges <- aggregate(predictor_ranges[-c(1,2,5)], by=list(predictor_ranges$Group.1), FUN=median)

#Plot partial dependence of the main models.
pd_plot_data2$predictor <- factor(pd_plot_data2$predictor, levels=median_predictor_ranges$Group.1)
pd_plot_data2$yhat_1 <- 1 + pd_plot_data2$yhat
pd_plot_data_age <- pd_plot_data2[which(pd_plot_data2$predictor %in% "BL_AGE"),]
pd_plot_data_sex <- pd_plot_data2[which(pd_plot_data2$predictor %in% "MEN"),]
pd_plot_data_sex$value<- ifelse(pd_plot_data_sex$value == 1, "Male", "Female")
pd_plot_data_bal <- pd_plot_data2[-which(pd_plot_data2$predictor %in% c("BL_AGE", "MEN")),]
median_predictor_ranges_plot <- data.frame(predictor=rep(median_predictor_ranges$Group.1, 2), yhat_1=1 + unlist(median_predictor_ranges[,2:3]), value=unlist(median_predictor_ranges[,4:5]))
median_predictor_ranges_plot_age <- median_predictor_ranges_plot[which(median_predictor_ranges_plot$predictor %in% "BL_AGE"),]
median_predictor_ranges_plot_sex <- median_predictor_ranges_plot[which(median_predictor_ranges_plot$predictor %in% "MEN"),]
median_predictor_ranges_plot_sex$value <- ifelse(median_predictor_ranges_plot_sex$value == 1, "Male", "Female")
median_predictor_ranges_plot_bal <- median_predictor_ranges_plot[-which(median_predictor_ranges_plot$predictor %in% c("BL_AGE", "MEN")),]
#flip balances so that all connections are positive
for (predictor in unique(median_predictor_ranges_plot_bal$predictor)) {
  if (median_predictor_ranges[which(median_predictor_ranges$Group.1 %in% predictor),"min_yhat_1"] > median_predictor_ranges[which(median_predictor_ranges$Group.1 %in% predictor),"max_yhat_1"]) {
  median_predictor_ranges_plot_bal[which(median_predictor_ranges_plot_bal$predictor %in% predictor),"value"] <- median_predictor_ranges_plot_bal[which(median_predictor_ranges_plot_bal$predictor %in% predictor),"value"] * -1
  pd_plot_data_bal[which(pd_plot_data_bal$predictor %in% predictor),"value"] <- pd_plot_data_bal[which(pd_plot_data_bal$predictor %in% predictor),"value"] * -1
  }
}
p1 <- ggplot(data=pd_plot_data_age, aes(x=value, y=yhat_1, group=fold)) + geom_line(alpha=0.25, color="black") + scale_y_continuous(limits=c(0.05,0.9), breaks=seq(0.2,0.8,0.2)) +
  geom_point(data=median_predictor_ranges_plot_age, aes(x=value, y=yhat_1), size=2, color="black", shape=4) + theme(axis.title.y=element_blank()) + xlab("Age at baseline")
p2 <- ggplot(data=pd_plot_data_sex, aes(x=value, y=yhat_1, group=fold)) + geom_line(alpha=0.25, color="black") + scale_y_continuous(limits=c(0.05,0.9), breaks=seq(0.2,0.8,0.2)) +
  geom_point(data=median_predictor_ranges_plot_sex, aes(x=value, y=yhat_1), size=2, color="black", shape=4) + theme(axis.title.y=element_blank()) + xlab("Sex")
p3 <- ggplot(data=pd_plot_data_bal, aes(x=value, y=yhat_1, group=fold)) + geom_line(alpha=0.25, color="black") + facet_wrap(~predictor, scales="free_x", ncol=4) + scale_y_continuous(limits=c(0.2,0.8), breaks=seq(0.2,0.8,0.2)) +
  geom_point(data=median_predictor_ranges_plot_bal, aes(x=value, y=yhat_1), size=2, color="black", shape=4) + scale_x_continuous(limits=c(-10.05,10), breaks=seq(-10,10,5))+ geom_vline(xintercept=0, color="gray") + 
  theme(axis.title.y=element_blank()) + xlab("PhILR balance value")
p <- ((p1 / p2) | p3) + plot_layout(widths=c(1,3))
p0 <- wrap_plots(grid::textGrob("Relative predicted score", rot=90, gp=grid::gpar(fontsize = 20)), p) + plot_layout(widths=c(1,40))
ggsave(file = "Liver_analysis_antibiotics/classification_pd_plot.svg", plot=p0, units="cm", width=40, height=25)

#Check how excluding the validation region from feature selection and hyperparameter optimization affects the performance of the models.
#Read the top50 features in classification for each region separately
top_predictors_class <- readRDS("Liver_analysis_antibiotics/top_predictors_class.RDs")

#Include all unique features in the top50 of each region as possible features in the test data
top50_predictors_class <- unique(unlist(top_predictors_class))
test_data <- as.matrix(FLI_ML_test[,colnames(FLI_ML_test) %in% c("FLI", top50_predictors_class)])
dtest <- xgb.DMatrix(data = test_data[,-1], label = test_data[,1])
#For sensitivity analysis, optionally test covariates and balances separately
train_data_subset_class <- list(as.matrix(FLI_ML_train[,colnames(FLI_ML_train) %in% c("FLI", top50_predictors_class)]), as.matrix(FLI_ML_train[,colnames(FLI_ML_train) %in% c("FLI", c("MEN", "BL_AGE", "ALKI2_FR02"))]), as.matrix(FLI_ML_train[,colnames(FLI_ML_train) %in% c("FLI", top50_predictors_class[-which(top50_predictors_class %in% c("MEN", "BL_AGE", "ALKI2_FR02"))])]))
test_data_class <- list(as.matrix(FLI_ML_test[,colnames(FLI_ML_test) %in% c("FLI", top50_predictors_class)]), as.matrix(FLI_ML_test[,colnames(FLI_ML_test) %in% c("FLI", c("MEN", "BL_AGE", "ALKI2_FR02"))]), as.matrix(FLI_ML_test[,colnames(FLI_ML_test) %in% c("FLI", top50_predictors_class[-which(top50_predictors_class %in% c("MEN", "BL_AGE", "ALKI2_FR02"))])]))

#Generate new optimal, individual hyperparameters for each LOGOCV model, leaving out data and features of one region at a time, for full separation between train and validation regions
run_class_folds <- list(NULL)
for (fold in 1:length(cv_folds)) {
obj.fun_sep_class <- smoof::makeSingleObjectiveFunction(
  name = "xgb_cv_bayes",
  fn = function(x) {
    train_sep_fold_class <- xgb.DMatrix(data = train_data_subset_class[[1]][,Reduce(intersect, top_predictors_class[-fold])], label = ifelse(train_data_subset_class[[1]][,1] >= 60, 1, 0))
    LOGOCV_mbo <- train_sep_fold_class[cv_folds[[fold]],]
    #Here, instead of testing in the region cross-validation setting, create a nested 70/30 split in the 5 included regions in the "train" set of the LOGOCV model. 
    set.seed(as.numeric(x["fold"])) #Use the "fold" variable in the parameter list as a repeatable random seed for the 70/30 nested splits
    train_idx <- createDataPartition(getinfo(LOGOCV_mbo, "label"), p = .7, list = FALSE, times = 1)
    test_idx <- setdiff(c(1:dim(LOGOCV_mbo)[1]), train_idx)
    train_mbo <- LOGOCV_mbo[train_idx,]
    test_mbo <- LOGOCV_mbo[test_idx,]
    watchlist <- list(train=train_mbo, test=test_mbo)
    set.seed(42)
    cv <- xgb.train(data = train_mbo,
      watchlist = watchlist,
      params = list(
      booster          = "gbtree",
      eta              = x["eta"],
      max_depth        = x["max_depth"],
      min_child_weight = x["min_child_weight"],
      gamma            = x["gamma"],
      subsample        = x["subsample"],
      colsample_bytree = x["colsample_bytree"],
      objective        = "binary:logistic", 
      eval_metric      = "auc"),
      nrounds          = x["nrounds"],
      showsd = TRUE,
      verbose = FALSE)
    
    as.numeric(cv$evaluation_log[max(iter),3])
  },
  par.set = getParamSet(obj.fun),
  minimize = FALSE
)

#Run hyperparameter optimization for the models, leaving out features and data of one region at a time
if (file.exists(paste0("Liver_analysis_antibiotics/xgboost_sep_parameters_class_", fold, ".RDs"))) {
  run_class <- readRDS(paste0("Liver_analysis_antibiotics/xgboost_sep_parameters_class_", fold, ".RDs"))
} else {
  control <- makeMBOControl()
  control <- setMBOControlTermination(control, iters = 100)
  run_class <- mbo(fun = obj.fun_sep_class, design = des[21:50,], control = control, show.info = TRUE)
  saveRDS(run_class, paste0("Liver_analysis_antibiotics/xgboost_sep_parameters_class_", fold, ".RDs"))
}
run_class_folds[[fold]] <- run_class
}

sens_codes <- c("covariates + balances", "covariates only", "balances only")
validation_stats_class <- list(NULL)
validation_predictors_class <- list(NULL)
validation_predictions_class <- list(NULL)
for (fold in 1:length(cv_folds)) {
    #For each LOGOCV model, omit the top50 features calculated in the test region and take the intersect of the other regions
    fold_features <- Reduce(intersect, top_predictors_class[-fold])
    validation_stats_sensset <- list(NULL)
    for (sensset in 1:3) {
      dtrain_class <- xgb.DMatrix(data = train_data_subset_class[[sensset]][,colnames(train_data_subset_class[[sensset]]) %in% fold_features], label = ifelse(train_data_subset_class[[sensset]][,1] >= 60, 1, 0))
      train_fold_class <- dtrain_class[cv_folds[[fold]],]
      dtest_class <- xgb.DMatrix(data = test_data_class[[sensset]][,colnames(test_data_class[[sensset]]) %in% fold_features], label = ifelse(test_data_class[[sensset]][,1] >= 60, 1, 0))
      test_fold_class <- dtest_class[which(test_fold_regions == test_fold_region_codes[fold,1]),]
      print(paste0("Classification model omitting region ", unique(train_groups[-cv_folds[[fold]]]), " tested on region ", test_fold_region_codes[fold,1], " (", test_fold_region_codes[fold,2], "), including ", sens_codes[sensset]))
      set.seed(42)
      fold_model_class <- xgboost(params = run_class_folds[[fold]]$x,
        objective = "binary:logistic", 
        eval_metric = "auc",
        data = train_fold_class,
        nrounds = run_class_folds[[fold]]$x$nrounds,
        verbose = F)
      predicted_class <- predict(fold_model_class, test_fold_class)
      if (sensset==1) {
        validation_predictions_class[[fold]] <- predicted_class
      }
      aucs <- evalmod(scores=predicted_class, labels=getinfo(test_fold_class, "label"))
      auc_values <- c(as.numeric(attr(aucs, "aucs")$aucs), as.numeric(attr(aucs, "data_info")$np/(attr(aucs, "data_info")$np+attr(aucs, "data_info")$nn)))
      validation_stats_sensset[[sensset]] <- auc_values
      print(paste0("Model AUC: ", round(auc_values[1], 4), " and AUPRC: ", round(auc_values[2], 4), " at baseline of ", round(auc_values[3], 2)*100, "% positive cases in ", dim(test_fold_class)[1], " participants, when geograpically extrapolating to ", test_fold_region_codes[fold,2]))
      xgb.save(fold_model_class, paste0("Liver_analysis_antibiotics/fold_model_class_sep", fold, "_", sensset, ".model"))
    }
    validation_stats_class[[fold]] <- as.data.frame(validation_stats_sensset, col.names=c(1:3))
    fold_model_class <- xgb.load(paste0("Liver_analysis_antibiotics/fold_model_class_sep", fold, "_1.model"))
    validation_predictors_class[[fold]] <- data.frame(xgb.importance(colnames(test_data_class[[1]][,-1]), model=fold_model_class))
}
validation_stats_class_sep_df <- data.frame(CV_region=c(as.character(test_fold_region_codes[,2]), "Mean"), Main_sep_model_AUC=round(c(unlist(lapply(validation_stats_class, `[[`, 1))[c(T,F,F)], mean(unlist(lapply(validation_stats_class, `[[`, 1))[c(T,F,F)])),3), Main_sep_model_covariates_AUC=round(c(unlist(lapply(validation_stats_class, `[[`, 2))[c(T,F,F)], mean(unlist(lapply(validation_stats_class, `[[`, 2))[c(T,F,F)])),3), Main_sep_model_balances_AUC=round(c(unlist(lapply(validation_stats_class, `[[`, 3))[c(T,F,F)], mean(unlist(lapply(validation_stats_class, `[[`, 3))[c(T,F,F)])),3), Main_sep_model_AUPPRC=round(c(unlist(lapply(validation_stats_class, `[[`, 1))[c(F,T,F)], mean(unlist(lapply(validation_stats_class, `[[`, 1))[c(F,T,F)])),3), Main_sep_model_covariates_AUPPRC=round(c(unlist(lapply(validation_stats_class, `[[`, 2))[c(F,T,F)], mean(unlist(lapply(validation_stats_class, `[[`, 2))[c(F,T,F)])),3), Main_sep_model_balances_AUPPRC=round(c(unlist(lapply(validation_stats_class, `[[`, 3))[c(F,T,F)], mean(unlist(lapply(validation_stats_class, `[[`, 3))[c(F,T,F)])),3), Test_set_N=c(as.numeric(table(test_fold_regions)[as.factor(test_fold_region_codes[,1])]), round(mean(table(test_fold_regions)),1)), Test_set_positives=round(c(unlist(lapply(validation_stats_class, `[[`, 1))[c(F,F,T)], mean(unlist(lapply(validation_stats_class, `[[`, 1))[c(F,F,T)])),3))
print(validation_stats_class_sep_df)
write.table(validation_stats_class_sep_df, "Liver_analysis_antibiotics/model_performance_sep_auc_auprc_N.tsv", sep="\t", row.names=F)

#Perform feature selection for triglycerides and GGT in BMI, age and sex groups to detect "liver-specific" features in the main models. 
#Group BMI and age
grouping_FLI_ML_data <- cbind(FLI_ML_data, data.frame(sample_data(gtdb_bac_data_tr)[,c("BMI", "TRIG", "GGT", "ALUE")]))
grouping_FLI_ML_data$TRIG <- grouping_FLI_ML_data$TRIG*88.57
grouping_FLI_ML_data$BMI[is.na(grouping_FLI_ML_data$BMI)] <- mean(grouping_FLI_ML_data$BMI, na.rm = TRUE)
grouping_FLI_ML_data$TRIG[is.na(grouping_FLI_ML_data$TRIG)] <- mean(grouping_FLI_ML_data$TRIG, na.rm = TRUE)
grouping_FLI_ML_data$GGT[is.na(grouping_FLI_ML_data$GGT)] <- mean(grouping_FLI_ML_data$GGT, na.rm = TRUE)
grouping_FLI_ML_data$BMI_category <- cut(grouping_FLI_ML_data$BMI, breaks=c(0, 25, 30, Inf), labels=c("underweight_normal", "overweight", "obese"))
grouping_FLI_ML_data$AGE_category <- cut(grouping_FLI_ML_data$BL_AGE, breaks=c(0, 40, 60, Inf), labels=c("<40", "40-60", "60<"))
grouping_FLI_ML_data$category <- factor(paste(grouping_FLI_ML_data$BMI_category, grouping_FLI_ML_data$AGE_category, ifelse(grouping_FLI_ML_data$MEN == 1, "MALE", "FEMALE"), sep="."))
grouping_folds <- groupKFold(grouping_FLI_ML_data$ALUE, k = length(unique(grouping_FLI_ML_data$ALUE)))
grouping_FLI_ML_data <- grouping_FLI_ML_data[,-which(colnames(grouping_FLI_ML_data) %in% c("FLI", "BMI", "BMI_category", "BL_AGE", "AGE_category", "MEN", "ALUE"))]
all_predictors_TRIG_GGT <- list(NULL)
for (fold in 1:length(grouping_folds)) {
	fold_data_TRIG <- grouping_FLI_ML_data[grouping_folds[[fold]], !names(grouping_FLI_ML_data) %in% "GGT"]
	fold_data_GGT <- grouping_FLI_ML_data[grouping_folds[[fold]], !names(grouping_FLI_ML_data) %in% "TRIG"]
  fold_predictors_TRIG_GGT <- list(NULL)
	for (grouping in levels(grouping_FLI_ML_data$category)) {
		grouping_data_TRIG <- fold_data_TRIG[fold_data_TRIG$category %in% grouping, !names(fold_data_TRIG) %in% "category"]
		fold_regr_task_TRIG <- makeRegrTask(data = grouping_data_TRIG, target = "TRIG")
		set.seed(42)
		fv_TRIG <- generateFilterValuesData(fold_regr_task_TRIG, method = "ranger_permutation")
		fold_predictors_TRIG <- data.frame(fv_TRIG$data[order(-fv_TRIG$data$value),])[1:50,] 
    fold_predictors_TRIG_GGT <- append(fold_predictors_TRIG_GGT, list(data.frame(features = fold_predictors_TRIG$name, type="TRIG", group = grouping, fold = fold)))
		grouping_data_GGT <- fold_data_GGT[fold_data_GGT$category %in% grouping, !names(fold_data_GGT) %in% "category"]
		fold_regr_task_GGT <- makeRegrTask(data = grouping_data_GGT, target = "GGT")
		set.seed(42)
		fv_GGT <- generateFilterValuesData(fold_regr_task_GGT, method = "ranger_permutation")
		fold_predictors_GGT <- data.frame(fv_GGT$data[order(-fv_GGT$data$value),])[1:50,] 
		fold_predictors_TRIG_GGT <- append(fold_predictors_TRIG_GGT, list(data.frame(features = fold_predictors_GGT$name, type="GGT", group = grouping, fold = fold)))
	}
  all_predictors_TRIG_GGT[[fold]] <- do.call(rbind, fold_predictors_TRIG_GGT)
  saveRDS(all_predictors_TRIG_GGT, "Liver_analysis_antibiotics/all_predictors_TRIG_GGT.RDs")
}
all_predictors_TRIG_GGT_df <- do.call(rbind, all_predictors_TRIG_GGT)
all_predictors_TRIG_GGT_df$type_group <- paste(all_predictors_TRIG_GGT_df$type, all_predictors_TRIG_GGT_df$group, sep="_")
all_predictors_TRIG_GGT_df <- table(all_predictors_TRIG_GGT_df$features, all_predictors_TRIG_GGT_df$type_group)
top_predictors_TRIG_GGT <- all_predictors_TRIG_GGT_df[which(apply(all_predictors_TRIG_GGT_df, 1, function (r) any(r == 6))),which(apply(all_predictors_TRIG_GGT_df, 2, function (r) any(r == 6)))]

#Prepare phylogenetic trees
new_ggtree <- as_tibble(phy_tree(gtdb_bac_data_tr))
all_taxonomy <- as.data.frame(gtdb_bac_data_tr@tax_table@.Data)
all_taxonomy$label <- rownames(all_taxonomy)
new_ggtree <- full_join(new_ggtree, all_taxonomy, by="label")
new_ggtree$Order_grp <- as.character(new_ggtree$Order)
new_ggtree$Order_grp <- gsub("o__", "", new_ggtree$Order_grp)
new_ggtree$label <- gsub("s__", "", new_ggtree$label)
new_ggtree$label <- gsub("_", " ", new_ggtree$label)
new_ggtree2 <- as.treedata(new_ggtree)@phylo
new_ggtree2_subset <- tree_subset(new_ggtree2, "n336", levels_back = 8)
new_ggtree_subset <- new_ggtree[which(new_ggtree$label %in% c(new_ggtree2_subset$tip.label, new_ggtree2_subset$node.label)),]

new_ggtree_groupinfo <- split(new_ggtree_subset$label, new_ggtree_subset$Order_grp)
new_ggtree2_subset <- ggtree::groupOTU(new_ggtree2_subset, new_ggtree_groupinfo)


#Perform pathway analysis of the genomes under the predictive balances.
GTDB_genome_metadata <- read.table("Liver_analysis_antibiotics/sp_clusters_r89.tsv", sep="\t", nrows=24707, header=T)
GTDB_genome_names <- GTDB_genome_metadata[,1:2]
GTDB_genome_names$Simp.name <- gsub("_", " ", gsub("s__", "", GTDB_genome_names$GTDB.species))
GTDB_genome_names$Simp.accession <- gsub("^[A-Z]{2}_", "", GTDB_genome_names$Representative.genome)
GTDB_genome_names <- GTDB_genome_names[which(GTDB_genome_names$Simp.name %in% new_ggtree2_subset$tip.label),c(3,4)]
#Subset a list of NCBI genomes to the target GTDB genomes
GTDB_genome_files <- readLines("Liver_analysis_antibiotics/assembly_summary_09032020.txt")[-c(1:2)]
GTDB_genome_files <- as.data.frame(do.call("rbind", lapply(strsplit(GTDB_genome_files, split="\t", fixed=TRUE), "[", c(1,18,20))))
GTDB_genome_files <- GTDB_genome_files[which(GTDB_genome_files$V1 %in% GTDB_genome_names$Simp.accession | GTDB_genome_files$V2 %in% GTDB_genome_names$Simp.accession),]
GTDB_genome_files[] <- lapply(GTDB_genome_files, as.character)
#Match the target genomes with the correct NCBI accession names
GTDB_genome_names$NCBI.accession <- NA
for (i in 1:nrow(GTDB_genome_names)) {
  NCBI.accession <- GTDB_genome_files[which(GTDB_genome_files$V1 %in% GTDB_genome_names$Simp.accession[i] | GTDB_genome_files$V2 %in% GTDB_genome_names$Simp.accession[i]),3]
  GTDB_genome_names$NCBI.accession[i] <- gsub("\\.", "_", gsub("^.*/(.*\\.[0-9]*)_.*$", "\\1", NCBI.accession))
}
GTDB_genome_files <- as.character(GTDB_genome_files[which(GTDB_genome_files$V1 %in% GTDB_genome_names$Simp.accession | GTDB_genome_files$V2 %in% GTDB_genome_names$Simp.accession),3])
for (genome in GTDB_genome_files){
  genome_name <- gsub("\\.", "_", gsub("^.*/(.*\\.[0-9]*)_.*$", "\\1", genome))
  genome_destination_dir <- paste0("Liver_analysis_antibiotics/genomes/", genome_name, "/")
  dir.create(genome_destination_dir)
  genome_gbff_destination <- paste0(genome_destination_dir, genome_name, ".gbk")
  if (!file.exists(genome_gbff_destination)) {
    genome_gbff_source <- paste0(genome, "/", gsub("^.*/(.*)$", "\\1", genome), "_genomic.gbff.gz")
    genome_gbff_destination <- paste0(genome_gbff_destination, ".gz")
    download.file(url=genome_gbff_source, destfile=genome_gbff_destination, method="wget")
    system(paste0("gunzip ", genome_gbff_destination))
  }
}

##########

###Following, the genomes are analyzed for MetaCyc pathways with:

#mpwt -f Liver_analysis_antibiotics/genomes/ --patho --clean --ignore-error --cpu=2 -v

#Genomes which have not been annotated need to be reanalyzed after annotation with Prokka:

#for genome in $(grep "build-pathologic-kb: Error, no proteins found. Pathologic build aborted." Liver_analysis_antibiotics/genomes/*/pathologic.log | awk -v FS="/" '{print $6}')
#do
#  genome_lower=$(echo ${genome} | awk '{print tolower($0)}')
#  cat pathwaytools/confdata/ptools-local/pgdbs/user/${genome_lower}cyc/1.0/input/*.fsa > Liver_analysis_antibiotics/genomes/${genome}/${genome}.fsa
#  prokka --force --cpus 0 --kingdom Bacteria --gcode 11 --gram pos --norrna --notrna --prefix ${genome} --outdir Liver_analysis_antibiotics/genomes/prokka/${genome}/ Liver_analysis_antibiotics/genomes/${genome}/$#{genome}.fsa
#done

###NCBI taxon ID 186801 (Class Clostridia) was used in the PathoLogic pathway inference
#grep "build-pathologic-kb: Error, no proteins found. Pathologic build aborted." Liver_analysis_antibiotics/genomes/*/pathologic.log | awk -v FS="/" -v OFS="\t" '{print $6, "186801"}' > Liver_analysis_antibiotics/genomes/prokka/taxon_id.tsv

###The .gff files created by Prokka need to be renamed to be able to use mpwt
#for file in Liver_analysis_antibiotics/genomes/prokka/*/*.gff; do mv ${file} ${file/.gff/_redundant.gff}; done

###Run mpwt for PathoLogic pathway inference
#mpwt -f Liver_analysis_antibiotics/genomes/prokka/ --clean --patho --ignore-error --cpu=2 -v --taxon-file

#cat Liver_analysis_antibiotics/liver_metacyc_pathways.tsv | while read line 
#do
# pathway=$(echo $line | awk '{print $1}')
# process=$(echo $line | cut -f 1 -d ' ' --complement)
# grep ${pathway} pathwaytools/confdata/ptools-local/pgdbs/user/*cyc/1.0/reports/pathways-report*.txt | awk -v OFS="\t" -v FS="_|cyc" -v pathway="${pathway}" -v process="${process}" '{print "GCA_" $2 "_" $3, pathway, process}'
#done > Liver_analysis_antibiotics/genome_pathway_inference_results.tsv

##########

#Collect results of the pathway analysis.
genome_pwy_results <- read.table("Liver_analysis_antibiotics/genome_pathway_inference_results.tsv", sep = "\t")
genome_pwy_results_table <- as.data.frame.matrix(table(genome_pwy_results[c(1,3)]))
missing_genomes <- data.frame(row.names=GTDB_genome_names$NCBI.accession[-which(GTDB_genome_names$NCBI.accession %in% row.names(genome_pwy_results_table))], matrix(0, ncol=ncol(genome_pwy_results_table), nrow=nrow(GTDB_genome_names)-nrow(genome_pwy_results_table)))
colnames(missing_genomes) <- colnames(genome_pwy_results_table)
genome_pwy_results_table <- rbind(genome_pwy_results_table, missing_genomes)
genome_pwy_results_table <- merge(GTDB_genome_names, genome_pwy_results_table, by.x="NCBI.accession", by.y="row.names")
write.csv(genome_pwy_results_table, "Liver_analysis_antibiotics/genome_pathway_inference_results_table.csv", row.names=F)
genome_pwy_results_table2 <- as.data.frame.matrix(table(genome_pwy_results[which(genome_pwy_results[,3] %in% c("Fermentation to Acetate", "Fermentation to Ethanol")),c(1,2)]))
missing_genomes2 <- data.frame(row.names=GTDB_genome_names$NCBI.accession[-which(GTDB_genome_names$NCBI.accession %in% row.names(genome_pwy_results_table2))], matrix(0, ncol=ncol(genome_pwy_results_table2), nrow=nrow(GTDB_genome_names)-nrow(genome_pwy_results_table2)))
colnames(missing_genomes2) <- colnames(genome_pwy_results_table2)
genome_pwy_results_table2 <- rbind(genome_pwy_results_table2, missing_genomes2)
genome_pwy_results_table2 <- merge(GTDB_genome_names, genome_pwy_results_table2, by.x="NCBI.accession", by.y="row.names")
write.csv(genome_pwy_results_table2, "Liver_analysis_antibiotics/genome_pathway_inference_results_table2.csv", row.names=F)

#Plot figures for the manuscript.
order_palette <- c("#000000","#8d70c9","#67a64e","#c8588c","#49adad","#cb5b42","#b69340")
tc_colors <- c("#9983b6","#c8119e","#019eba","#d55886","#0289d1","#fe91b4","#8882ff","#ff8de3","#484d8c","#96b4ff","#7d2f94")

top_balances <- as.character(validation_importances_class$Feature[grep("n", validation_importances_class$Feature)])
top_balance_labels <- as.character(validation_importances_class$balance[grep("n", validation_importances_class$Feature)])
top_balance_labels <- gsub("[a-z]__", "", top_balance_labels)
top_balance_labels <- gsub("_", " ", top_balance_labels)
top_balance_labels <- strsplit(top_balance_labels, "/")
tc_nn <- name.to.nn(new_ggtree2_subset, top_balances)

#formulate table for median yhat_1 values for each balance
median_predictor_ranges <- median_predictor_ranges[order(abs(median_predictor_ranges$max_yhat_1-median_predictor_ranges$min_yhat_1), decreasing = T),]
predictor_nodes <- grep("n[0-9]", median_predictor_ranges$Group.1, value=T)
balances_heatmap <- data.frame(matrix(nrow=length(new_ggtree2_subset$tip.label), ncol=length(predictor_nodes)))
rownames(balances_heatmap) <- new_ggtree2_subset$tip.label
colnames(balances_heatmap) <- predictor_nodes 
for (node in predictor_nodes){
   up_down_tips <- get.ud.tips(new_ggtree2_subset, node)
   balances_heatmap[rownames(balances_heatmap) %in% up_down_tips$up,node] <- median_predictor_ranges[median_predictor_ranges$Group.1 %in% node,"min_yhat_1"]
   balances_heatmap[rownames(balances_heatmap) %in% up_down_tips$down,node] <- median_predictor_ranges[median_predictor_ranges$Group.1 %in% node,"max_yhat_1"]
}
balances_heatmap2 <- round(1+balances_heatmap, 1)
rownames(balances_heatmap2) <- rownames(balances_heatmap)

#formulate table for number of pathways inferred for genomes, per each process
pathway_heatmap <- data.frame(row.names=genome_pwy_results_table$Simp.name, genome_pwy_results_table[-c(1:3)])
pathway_heatmap[pathway_heatmap == 0] <- NA
pathway_heatmap <- pathway_heatmap[,c(3,4,6,5,1,2)]

pathway_heatmap2 <- data.frame(row.names=genome_pwy_results_table2$Simp.name, genome_pwy_results_table2[-c(1:3)])
#Convert number of pathways to 1 or NA
pathway_heatmap2[pathway_heatmap2 == 0] <- NA
pathway_heatmap2[pathway_heatmap2 > 1] <- 1
#remove non-acetate and ethanol pathways
acetate_ethanol_pwys <- c("FERMENTATION.PWY", "P122.PWY", "PWY.5480", "PWY.6587", "P161.PWY", "P162.PWY", "P163.PWY", "PWY.5482", "PWY0.1312")
pathway_heatmap2 <- pathway_heatmap2[,acetate_ethanol_pwys]

p <- ggtree(tr = new_ggtree2_subset, ladderize = T, mapping = aes(color = group), branch.length ='none') + 
  scale_color_manual(values = order_palette, name = "Order", breaks = c("Oscillospirales", "4C28d-15", "Christensenellales", "TANB77", "Saccharofermentanales", "Lachnospirales")) + 
  geom_tiplab(aes(label=new_ggtree_subset$label), align=T, size = 3, show.legend=F) + geom_rootedge()
#p <- scaleClade(p, 572, .05) %>% scaleClade(551, .05) %>% scaleClade(604, .05) %>% scaleClade(667, .05) %>% scaleClade(467, .05) %>% scaleClade(473, .05) %>% scaleClade(515, .05) %>% scaleClade(451, .05) %>% scaleClade(425, .05) %>% scaleClade(421, .05) %>% scaleClade(395, .05) %>% scaleClade(344, .05)
#p <- p %>% collapse(572, 'min', fill=order_palette[4]) %>% collapse(551, 'min', fill=order_palette[4]) %>% collapse(604, 'min', fill=order_palette[4]) %>% collapse(667, 'min', fill=order_palette[4]) %>% collapse(467, 'min', fill=order_palette[3]) %>% collapse(473, 'min', fill=order_palette[2]) %>% collapse(515, 'min', fill=order_palette[7]) %>% collapse(451, 'min', fill=order_palette[5]) %>% collapse(425, 'min', fill=order_palette[5]) %>% collapse(421, 'min', fill=order_palette[5]) %>% collapse(395, 'min', fill=order_palette[5]) %>% collapse(344, 'min', fill=order_palette[5])
p <- gheatmap(p, balances_heatmap2, width=1.1, offset=22.5, colnames=T, color="black", colnames_position="top", font.size=3, colnames_angle=90, hjust=0) + scale_fill_distiller(palette="RdBu", na.value="gray50", values=scales::rescale(c(0.3, 0.4, 0.5, 0.6, 0.7)), limits=c(0.3, 0.7), breaks = c(0.3, 0.4, 0.5, 0.6, 0.7), guide="legend", name="Relative\npredicted score")
p2 <- p + new_scale_fill()
p2 <- gheatmap(p2, pathway_heatmap, width=0.6, offset=42, colnames=T, color="black", colnames_position="top", font.size=3, colnames_angle=90, hjust=0) + scale_fill_gradient(low = "yellow", high = "red", na.value = 0, guide = "legend", name="Number of\npathways")
p3 <- p2 + new_scale_fill()
p3 <- gheatmap(p3, pathway_heatmap2, width=0.9, offset=53, colnames=T, color="black", colnames_position="top", font.size=3, colnames_angle=90, hjust=0) + scale_fill_gradient(high = "black", na.value = 0, guide = "legend", name="Presence of\npathway")
p3 <- p3 + xlim_tree(50) + expand_limits(y=363)
p3 <- p3 + geom_nodepoint(aes(subset=node %in% tc_nn), shape=18, size=3, color="black") + geom_nodelab(aes(subset=node %in% tc_nn, label=label), color="black", vjust=-.5, hjust=1.1, size=3)

ggsave(file = "Liver_analysis_antibiotics/pathway_tree.pdf", plot=p3, units="cm", width=27, height=80, limitsize=F)

#Construct alternative models, where participants with FLI between 30 and 60 are excluded.
#Collect the data into one object by concatenating FLI, covariates, and the bacterial and archaeal PhILR balances.
FLI_ML_data <- cbind(data.frame(sample_data(gtdb_bac_data_tr)[,c("FLI", "MEN", "BL_AGE", "ALKI2_FR02")]), as(otu_table(gtdb_bac_philr, taxa_are_rows = FALSE), "matrix"))
FLI_ML_arch_data <- as(otu_table(gtdb_arch_philr, taxa_are_rows = FALSE), "matrix")
colnames(FLI_ML_arch_data) <- paste0(colnames(FLI_ML_arch_data), "_a")
FLI_ML_data <- cbind(FLI_ML_data, FLI_ML_arch_data)
#Replace zeros
FLI_ML_data$FLI[is.na(FLI_ML_data$FLI)] <- mean(FLI_ML_data$FLI, na.rm = TRUE)
FLI_ML_data$ALKI2_FR02[is.na(FLI_ML_data$ALKI2_FR02)] <- mean(FLI_ML_data$ALKI2_FR02, na.rm = TRUE)
#Remove FLI 30 => 60
FLI_ML_data <- FLI_ML_data[which(FLI_ML_data$FLI >= 60 | FLI_ML_data$FLI < 30),]
#Divide the data first to 70% train 30% test preserving sex and area class balance
part_df <- data.frame(sample_data(gtdb_bac_data_tr))
part_df <- part_df[which(part_df$FLI >= 60 | part_df$FLI < 30),]
part_criteria <- paste0(part_df$MEN, part_df$ALUE)
set.seed(42)
trainIndex <- createDataPartition(part_criteria, p = .7, list = FALSE, times = 1)
FLI_ML_train <- FLI_ML_data[trainIndex,]
FLI_ML_test <- FLI_ML_data[-trainIndex,]
train_groups <- part_df$ALUE[trainIndex]
cv_folds <- groupKFold(train_groups, k = length(unique(train_groups)))
train_data <- as.matrix(FLI_ML_train)

#Perform feature selection and hyperparameter optimization for the alternative models.
#Find the optimal predictors by random forest permutation importance in mlr and hyperparameters through Bayesian optimization (adapted from https://www.simoncoulombe.com/2019/01/bayesian/)
obj.fun <- smoof::makeSingleObjectiveFunction(
  name = "xgb_cv_bayes",
  fn = function(x) {
    train_mbo <- dtrain[cv_folds[[as.numeric(x["fold"])]],]
    test_idx <- setdiff(c(1:dim(dtrain)[1]), cv_folds[[as.numeric(x["fold"])]])
    test_mbo <- dtrain[test_idx,]
    watchlist <- list(train=train_mbo, test=test_mbo)
    set.seed(42)
    cv <- xgb.train(data = train_mbo,
      watchlist = watchlist,
      params = list(
      booster          = "gbtree",
      eta              = x["eta"],
      max_depth        = x["max_depth"],
      min_child_weight = x["min_child_weight"],
      gamma            = x["gamma"],
      subsample        = x["subsample"],
      colsample_bytree = x["colsample_bytree"],
      objective        = "reg:squarederror", 
      eval_metric      = "rmse"),
      nrounds          = x["nrounds"],
      showsd = TRUE,
      verbose = FALSE)
    
    as.numeric(cv$evaluation_log[max(iter),3])
  },
  par.set = makeParamSet(
    makeNumericParam("eta",              lower = 0.001,  upper = 0.3),
    makeNumericParam("gamma",            lower = 0.1,   upper = 5),
    makeIntegerParam("max_depth",        lower = 2,     upper = 8),
    makeIntegerParam("min_child_weight", lower = 1,     upper = 10),
    makeNumericParam("subsample",        lower = 0.2,   upper = 0.8), #set maximum to 80%
    makeNumericParam("colsample_bytree", lower = 0.2,   upper = 0.9),  #set maximum to 90%
    makeIntegerParam("nrounds",          lower = 50,    upper = 5000),
    makeIntegerParam("fold",             lower = 1,     upper = 6, tunable = FALSE)
  ),
  minimize = TRUE
)
obj.fun_class <- smoof::makeSingleObjectiveFunction(
  name = "xgb_cv_bayes",
  fn = function(x) {
    train_mbo <- dtrain_class[cv_folds[[as.numeric(x["fold"])]],]
    test_idx <- setdiff(c(1:dim(dtrain_class)[1]), cv_folds[[as.numeric(x["fold"])]])
    test_mbo <- dtrain_class[test_idx,]
    watchlist <- list(train=train_mbo, test=test_mbo)
    set.seed(42)
    cv <- xgb.train(data = train_mbo,
      watchlist = watchlist,
      params = list(
      booster          = "gbtree",
      eta              = x["eta"],
      max_depth        = x["max_depth"],
      min_child_weight = x["min_child_weight"],
      gamma            = x["gamma"],
      subsample        = x["subsample"],
      colsample_bytree = x["colsample_bytree"],
      objective        = "binary:logistic", 
      eval_metric      = "auc"),
      nrounds          = x["nrounds"],
      showsd = TRUE,
      verbose = FALSE)
    
    as.numeric(cv$evaluation_log[max(iter),3])
  },
  par.set = getParamSet(obj.fun),
  minimize = FALSE
)
#Generate random sets of the parameter space
set.seed(42)
des <- generateDesign(n=50, par.set = getParamSet(obj.fun), fun = lhs::randomLHS)
#Gather a set of consensus most important features present in all models: intersect of top 50 predictors (ranger permutation importance) in all folds
if (file.exists("Liver_analysis_30_60/top_predictors_class.RDs")) {
  top_predictors_class <- readRDS("Liver_analysis_30_60/top_predictors_class.RDs")
} else {
  top_predictors_class <- list(NULL)
  for (fold in 1:length(cv_folds)) {
  #Feature selection for classification
class_fold_data <- as.data.frame(train_data[cv_folds[[fold]],])
class_fold_data$FLI <- as.factor(ifelse(class_fold_data$FLI >= 60, 1, 0))
fold_class_task <- makeClassifTask(data = class_fold_data, target = "FLI", positive = "1")
set.seed(42)
fv_class <- generateFilterValuesData(fold_class_task, method = "ranger_permutation")
fold_predictors_class <- data.frame(fv_class$data[order(-fv_class$data$value),])[1:50,]
  top_predictors_class[[fold]] <- fold_predictors_class$name
  }
  print(top_predictors_class)
  saveRDS(top_predictors_class, "Liver_analysis_30_60/top_predictors_class.RDs")
}
top_predictors_class <- Reduce(intersect, top_predictors_class)

#Subset features in the data based on results of the feature selection
train_data_subset_class <- as.matrix(FLI_ML_train[,colnames(FLI_ML_train) %in% c("FLI", top_predictors_class)])
dtrain_class <- xgb.DMatrix(data = train_data_subset_class[,-1], label = ifelse(train_data_subset_class[,1] >= 60, 1, 0))

#Run hyperparameter optimization with the feature selected models
if (file.exists("Liver_analysis_30_60/xgboost_parameters_class.RDs")) {
  run_class <- readRDS("Liver_analysis_30_60/xgboost_parameters_class.RDs")
} else {
  control <- makeMBOControl()
  control <- setMBOControlTermination(control, iters = 100)
  run_class <- mbo(fun = obj.fun_class, design = des[21:50,], control = control, show.info = TRUE)
  saveRDS(run_class, "Liver_analysis_30_60/xgboost_parameters_class.RDs")
}

#Train the alternative models.
test_fold_regions <- part_df$ALUE[-trainIndex]
#For sensitivity analysis, optionally test covariates and balances separately
train_data_subset_class <- list(as.matrix(FLI_ML_train[,colnames(FLI_ML_train) %in% c("FLI", top_predictors_class)]), as.matrix(FLI_ML_train[,colnames(FLI_ML_train) %in% c("FLI", c("MEN", "BL_AGE"))]), as.matrix(FLI_ML_train[,colnames(FLI_ML_train) %in% c("FLI", top_predictors_class[-which(top_predictors_class %in% c("MEN", "BL_AGE"))])]))
test_data_class <- list(as.matrix(FLI_ML_test[,colnames(FLI_ML_test) %in% c("FLI", top_predictors_class)]), as.matrix(FLI_ML_test[,colnames(FLI_ML_test) %in% c("FLI", c("MEN", "BL_AGE"))]), as.matrix(FLI_ML_test[,colnames(FLI_ML_test) %in% c("FLI", top_predictors_class[-which(top_predictors_class %in% c("MEN", "BL_AGE"))])]))
validation_stats_class <- list(NULL)
validation_predictors_class <- list(NULL)
validation_predictions_class <- list(NULL)
pd_plot_data <- list(NULL)
for (fold in 1:length(cv_folds)) {
    validation_stats_sensset <- list(NULL)
    for (sensset in 1:3) {
      dtrain_class <- xgb.DMatrix(data = train_data_subset_class[[sensset]][,-1], label = ifelse(train_data_subset_class[[sensset]][,1] >= 60, 1, 0))
      train_fold_class <- dtrain_class[cv_folds[[fold]],]
      dtest_class <- xgb.DMatrix(data = test_data_class[[sensset]][,-1], label = ifelse(test_data_class[[sensset]][,1] >= 60, 1, 0))
      test_fold_class <- dtest_class[which(test_fold_regions == test_fold_region_codes[fold,1]),]
      print(paste0("Classification model omitting region ", unique(train_groups[-cv_folds[[fold]]]), " tested on region ", test_fold_region_codes[fold,1], " (", test_fold_region_codes[fold,2], "), including ", sens_codes[sensset]))
      set.seed(42)
      fold_model_class <- xgboost(params = run_class$x,
        objective = "binary:logistic", 
        eval_metric = "auc",
        data = train_fold_class,
        nrounds = run_class$x$nrounds,
        verbose = F)
      predicted_class <- predict(fold_model_class, test_fold_class)
      if (sensset==1) {
        validation_predictions_class[[fold]] <- predicted_class
      }
      aucs <- evalmod(scores=predicted_class, labels=getinfo(test_fold_class, "label"))
      auc_values <- c(as.numeric(attr(aucs, "aucs")$aucs), as.numeric(attr(aucs, "data_info")$np/(attr(aucs, "data_info")$np+attr(aucs, "data_info")$nn)))
      validation_stats_sensset[[sensset]] <- auc_values
      print(paste0("Model AUC: ", round(auc_values[1], 4), " and AUPRC: ", round(auc_values[2], 4), " at baseline of ", round(auc_values[3], 2)*100, "% positive cases in ", dim(test_fold_class)[1], " participants, when geographically extrapolating to ", test_fold_region_codes[fold,2]))
      xgb.save(fold_model_class, paste0("Liver_analysis_30_60/fold_model_class", fold, "_", sensset, ".model"))
    }
    validation_stats_class[[fold]] <- as.data.frame(validation_stats_sensset, col.names=c(1:3))
    fold_model_class <- xgb.load(paste0("Liver_analysis_30_60/fold_model_class", fold, "_1.model"))
    validation_predictors_class[[fold]] <- data.frame(xgb.importance(colnames(test_data_class[[1]][,-1]), model=fold_model_class))
    #Gather partial dependence of the variables from the model (takes quite long to compute)
    #Recreate the full model (partial doesn't work with loaded xgboost models)
    dtrain_class <- xgb.DMatrix(data = train_data_subset_class[[1]][,-1], label = ifelse(train_data_subset_class[[1]][,1] >= 60, 1, 0))
    train_fold_class <- dtrain_class[cv_folds[[fold]],]
    dtest_class <- xgb.DMatrix(data = test_data_class[[1]][,-1], label = ifelse(test_data_class[[1]][,1] >= 60, 1, 0))
    test_fold_class <- dtest_class[which(test_fold_regions == test_fold_region_codes[fold,1]),]
    print(paste0("Classification model omitting region ", unique(train_groups[-cv_folds[[fold]]]), " tested on region ", test_fold_region_codes[fold,1], " (", test_fold_region_codes[fold,2], "), including ", sens_codes[sensset]))
    set.seed(42)
    fold_model_class <- xgboost(params = run_class$x,
                                objective = "binary:logistic", 
                                eval_metric = "auc",
                                data = train_fold_class,
                                nrounds = run_class$x$nrounds,
                                verbose = F)
    pd_train <- train_data_subset_class[[1]][cv_folds[[fold]], -1]
    pd_data <- list(NULL)
    for (predictor in 1:ncol(pd_train)) {
      pd_df <- partial(fold_model_class, train = pd_train, pred.var = colnames(pd_train)[predictor], plot=FALSE)
      print(paste0("Partial dependence of ", colnames(pd_train)[predictor], " calculated in fold ", fold))
      pd_df$predictor <- colnames(pd_train)[predictor]
      colnames(pd_df)[1] <- "value"
      pd_data[[predictor]] <- pd_df
    }
    pd_data <- do.call(rbind, pd_data)
    pd_data$fold <- fold
    pd_plot_data[[fold]] <- pd_data
}
validation_stats_class_df <- cbind(validation_stats_class_df, data.frame(Alt_model=round(c(unlist(lapply(validation_stats_class, `[[`, 1))[c(T,F,F)], mean(unlist(lapply(validation_stats_class, `[[`, 1))[c(T,F,F)])),3), Alt_model_covariates=round(c(unlist(lapply(validation_stats_class, `[[`, 2))[c(T,F,F)], mean(unlist(lapply(validation_stats_class, `[[`, 2))[c(T,F,F)])),3), Alt_model_balances=round(c(unlist(lapply(validation_stats_class, `[[`, 3))[c(T,F,F)], mean(unlist(lapply(validation_stats_class, `[[`, 3))[c(T,F,F)])),3), Alt_model_test_set_N=c(as.numeric(table(test_fold_regions)[as.factor(test_fold_region_codes[,1])]), round(mean(table(test_fold_regions)),1))))
validation_stats_class_auprc_df <- cbind(validation_stats_class_auprc_df, data.frame(Alt_model=round(c(unlist(lapply(validation_stats_class, `[[`, 1))[c(F,T,F)], mean(unlist(lapply(validation_stats_class, `[[`, 1))[c(F,T,F)])),3), Alt_model_covariates=round(c(unlist(lapply(validation_stats_class, `[[`, 2))[c(F,T,F)], mean(unlist(lapply(validation_stats_class, `[[`, 2))[c(F,T,F)])),3), Alt_model_balances=round(c(unlist(lapply(validation_stats_class, `[[`, 3))[c(F,T,F)], mean(unlist(lapply(validation_stats_class, `[[`, 3))[c(F,T,F)])),3), Alt_model_test_set_N=c(as.numeric(table(test_fold_regions)[as.factor(test_fold_region_codes[,1])]), round(mean(table(test_fold_regions)),1)), Alt_model_test_set_positives=round(c(unlist(lapply(validation_stats_class, `[[`, 1))[c(F,F,T)], mean(unlist(lapply(validation_stats_class, `[[`, 1))[c(F,F,T)])),3)))
write.table(validation_stats_class_df, "Liver_analysis_antibiotics/model_performance_auc_N.tsv", sep="\t", row.names=F)
write.table(validation_stats_class_auprc_df, "Liver_analysis_antibiotics/model_performance_auprc_N.tsv", sep="\t", row.names=F)
validation_predictors_class <- do.call(rbind, validation_predictors_class)
validation_predictions_class <- do.call(cbind, validation_predictions_class)
#validation_predictions_class_compare <- cbind(getinfo(dtest_class, "label"), rowMeans(validation_predictions_class))
validation_importances_class <- do.call(data.frame, aggregate(. ~ Feature, validation_predictors_class, function(x) c(mean = mean(x), sd = sd(x))))
validation_importances_class <- validation_importances_class[order(-validation_importances_class$Gain.mean),]
predictor_balances_class <- list(NULL)
for (balance in validation_importances_class$Feature) { 
  if (balance %in% c("MEN", "BL_AGE", "ALKI2_FR02")) {
  predictor_balances_class <- append(predictor_balances_class, NA)
  } else {
  predictor_balances_class <- append(predictor_balances_class, name.balance(phy_tree(gtdb_bac_data_tr), tax_table(gtdb_bac_data_tr), balance))
  }
}
validation_importances_class$balance <- unlist(predictor_balances_class)
pd_plot_data2 <- do.call(rbind, pd_plot_data)
predictor_ranges <- list(NULL)
for (fold in 1:length(cv_folds)) {
  fold_pd_data <- pd_plot_data2[which(pd_plot_data2$fold == fold),]
  fold_predictor_ranges <- aggregate(fold_pd_data["value"], by=list(fold_pd_data$predictor), FUN=range)
  fold_yhat_1_values <- list(NULL)
  for (predictor in 1:length(fold_predictor_ranges$Group.1)){
    min_yhat_1 <- fold_pd_data[which(fold_pd_data$value == fold_predictor_ranges$value[predictor,1] & fold_pd_data$predictor == fold_predictor_ranges$Group.1[predictor]),"yhat"]
    max_yhat_1 <- fold_pd_data[which(fold_pd_data$value == fold_predictor_ranges$value[predictor,2] & fold_pd_data$predictor == fold_predictor_ranges$Group.1[predictor]),"yhat"]
    fold_yhat_1_values[[predictor]] <- data.frame(min_yhat_1 = min_yhat_1, max_yhat_1 = max_yhat_1)
    }
  fold_predictor_ranges <- cbind(fold_predictor_ranges, do.call(rbind, fold_yhat_1_values))
  fold_predictor_ranges$fold <- fold
  predictor_ranges[[fold]] <- fold_predictor_ranges
}
predictor_ranges <- do.call(rbind, predictor_ranges)
predictor_ranges$min_value <- predictor_ranges$value[,1]
predictor_ranges$max_value <- predictor_ranges$value[,2]
median_predictor_ranges <- aggregate(predictor_ranges[-c(1,2,5)], by=list(predictor_ranges$Group.1), FUN=median)

#Plot partial dependence of the features in the alternative models.
pd_plot_data2$predictor <- factor(pd_plot_data2$predictor, levels=as.character(median_predictor_ranges$Group.1))
pd_plot_data2$yhat_1 <- 1 + pd_plot_data2$yhat
pd_plot_data_age <- pd_plot_data2[which(pd_plot_data2$predictor %in% "BL_AGE"),]
pd_plot_data_sex <- pd_plot_data2[which(pd_plot_data2$predictor %in% "MEN"),]
pd_plot_data_sex$value<- ifelse(pd_plot_data_sex$value == 1, "Male", "Female")
pd_plot_data_bal <- pd_plot_data2[-which(pd_plot_data2$predictor %in% c("BL_AGE", "MEN")),]
median_predictor_ranges_plot <- data.frame(predictor=rep(median_predictor_ranges$Group.1, 2), yhat_1=1 + unlist(median_predictor_ranges[,2:3]), value=unlist(median_predictor_ranges[,4:5]))
median_predictor_ranges_plot_age <- median_predictor_ranges_plot[which(median_predictor_ranges_plot$predictor %in% "BL_AGE"),]
median_predictor_ranges_plot_sex <- median_predictor_ranges_plot[which(median_predictor_ranges_plot$predictor %in% "MEN"),]
median_predictor_ranges_plot_sex$value <- ifelse(median_predictor_ranges_plot_sex$value == 1, "Male", "Female")
median_predictor_ranges_plot_bal <- median_predictor_ranges_plot[-which(median_predictor_ranges_plot$predictor %in% c("BL_AGE", "MEN")),]
#flip balances so that all connections are positive
for (predictor in unique(median_predictor_ranges_plot_bal$predictor)) {
  if (median_predictor_ranges[which(median_predictor_ranges$Group.1 %in% predictor),"min_yhat_1"] > median_predictor_ranges[which(median_predictor_ranges$Group.1 %in% predictor),"max_yhat_1"]) {
  median_predictor_ranges_plot_bal[which(median_predictor_ranges_plot_bal$predictor %in% predictor),"value"] <- median_predictor_ranges_plot_bal[which(median_predictor_ranges_plot_bal$predictor %in% predictor),"value"] * -1
  pd_plot_data_bal[which(pd_plot_data_bal$predictor %in% predictor),"value"] <- pd_plot_data_bal[which(pd_plot_data_bal$predictor %in% predictor),"value"] * -1
  }
}
p1 <- ggplot(data=pd_plot_data_age, aes(x=value, y=yhat_1, group=fold)) + geom_line(alpha=0.25, color="black") + scale_y_continuous(limits=c(0.3,1.1), breaks=seq(0.4,1,0.2)) +
  geom_point(data=median_predictor_ranges_plot_age, aes(x=value, y=yhat_1), size=2, color="black", shape=4) + theme(axis.title.y=element_blank()) + xlab("Age at baseline")
p2 <- ggplot(data=pd_plot_data_sex, aes(x=value, y=yhat_1, group=fold)) + geom_line(alpha=0.25, color="black") + scale_y_continuous(limits=c(0.3,1.1), breaks=seq(0.4,1,0.2)) +
  geom_point(data=median_predictor_ranges_plot_sex, aes(x=value, y=yhat_1), size=2, color="black", shape=4) + theme(axis.title.y=element_blank()) + xlab("Sex")
p3 <- ggplot(data=pd_plot_data_bal, aes(x=value, y=yhat_1, group=fold)) + geom_line(alpha=0.25, color="black") + facet_wrap(~predictor, scales="free_x", ncol=4) + scale_y_continuous(limits=c(0.7,1), breaks=seq(0.8,1,0.1)) +
  geom_point(data=median_predictor_ranges_plot_bal, aes(x=value, y=yhat_1), size=2, color="black", shape=4) + scale_x_continuous(limits=c(-16.2,26.05), breaks=seq(-10,20,10))+ geom_vline(xintercept=0, color="gray") + 
  theme(axis.title.y=element_blank()) + xlab("PhILR balance value")
p <- ((p1 / p2) | p3) + plot_layout(widths=c(1,3))
p0 <- wrap_plots(grid::textGrob("Relative predicted score", rot=90, gp=grid::gpar(fontsize = 20)), p) + plot_layout(widths=c(1,40))
ggsave(file = "Liver_analysis_30_60/classification_pd_plot_alt.svg", plot=p0, units="cm", width=40, height=25)

#preprocess validation data similarly to the main FR02 data
#load the up-to-date phenotype data (object FR07). Contact person: Aki Havulinna, aki.havulinna(at)helsinki.fi
load("FR07_phenotype_data.Rdata")
FR07 <- FR07[!is.na(FR07$STOOL_ID),]
FR07$STOOL_ID <- gsub("(\\d{9})(\\d{1})$", "\\1-\\2", FR07$STOOL_ID)
row.names(FR07) <- FR07$STOOL_ID

if (file.exists("Liver_analysis_antibiotics/gtdb_bac_data_valid.RDs") && file.exists("Liver_analysis_antibiotics/gtdb_arch_data_valid.RDs")) {
  gtdb_bac_data_valid <- readRDS("Liver_analysis_antibiotics/gtdb_bac_data_valid.RDs")
  gtdb_arch_data_valid <- readRDS("Liver_analysis_antibiotics/gtdb_arch_data_valid.RDs")
} else {
  gtdb_data_valid <- import_biom("Liver_analysis_antibiotics/table.biom", parallel=TRUE)
  gtdb_data_valid <- prune_samples(row.names(FR07), gtdb_data_valid)
  #Format the tax table.
  colnames(tax_table(gtdb_data_valid)) <- c("Domain", "Phylum", "Class", "Order", "Family",  "Genus", "Species")
  #Format taxa names to the last defined rank to make them match the GTDB tree.
  gtdb_names_valid <- as.vector(tax_table(gtdb_data_valid)[,7])
  gtdb_names_valid[which(gtdb_names_valid != "s__")] <- paste0("s__", gsub("g__", "", as.vector(tax_table(gtdb_data_valid)[which(gtdb_names_valid != "s__"),6])), "_", gsub("s__", "", as.vector(tax_table(gtdb_data_valid)[which(gtdb_names_valid != "s__"),7])))
  gtdb_names_valid[which(gtdb_names_valid == "s__")] <- as.vector(tax_table(gtdb_data_valid)[which(gtdb_names_valid == "s__"),6])
  gtdb_names_valid[which(gtdb_names_valid == "g__")] <- as.vector(tax_table(gtdb_data_valid)[which(gtdb_names_valid == "g__"),5])
  gtdb_names_valid[which(gtdb_names_valid == "f__")] <- as.vector(tax_table(gtdb_data_valid)[which(gtdb_names_valid == "f__"),4])
  taxa_names(gtdb_data_valid) <- gtdb_names_valid
  #Also format species names in the tax table to match the new taxa names (to get the full species names)
  tax_table(gtdb_data_valid)[,"Species"] <- taxa_names(gtdb_data_valid)
  #Separate controls from samples in the sample data table
  sample_data(gtdb_data_valid) <- data.frame(row.names=sample_names(gtdb_data_valid), Name=sample_names(gtdb_data_valid), Type=ifelse(nchar(sample_names(gtdb_data_valid)) < 11, "Control", "Sample"))
  #Combine with gtdb bacterial and archaeal trees so that only species-level data is preserved. These files can be found in the GTDB repository: https://data.ace.uq.edu.au/public/gtdb/data/releases/release89/89.0/
  gtdb_bac_tree <- read_tree("Liver_analysis_antibiotics/bac120_r89.sp_labels_underscore.tree")
  gtdb_bac_data_valid <- merge_phyloseq(gtdb_data_valid, gtdb_bac_tree)
  gtdb_arch_tree <- read_tree("Liver_analysis_antibiotics/ar122_r89.sp_labels_underscore.tree")
  gtdb_arch_data_valid <- merge_phyloseq(gtdb_data_valid, gtdb_arch_tree)
  #remove samples with less than 50k reads (total)
  to_be_pruned_valid <- sample_sums(gtdb_bac_data_valid) + sample_sums(gtdb_arch_data_valid) > 50000
  gtdb_bac_data_valid <- prune_samples(to_be_pruned_valid, gtdb_bac_data_valid)
  gtdb_arch_data_valid <- prune_samples(to_be_pruned_valid, gtdb_arch_data_valid)
  #Combine the phenotype data with the bacterial and archaeal data (subsets the objects at the same time to samples)
  gtdb_bac_data_valid <- phyloseq(otu_table(gtdb_bac_data_valid), phy_tree(gtdb_bac_data_valid), tax_table(gtdb_bac_data_valid), sample_data(FR07))
  gtdb_arch_data_valid <- phyloseq(otu_table(gtdb_arch_data_valid), phy_tree(gtdb_arch_data_valid), tax_table(gtdb_arch_data_valid), sample_data(FR07))
  #remove pregnant (GRAVID==2) participants
  gtdb_bac_data_valid <- subset_samples(gtdb_bac_data_valid, GRAVID %in% c(1, NA))
  gtdb_arch_data_valid <- subset_samples(gtdb_arch_data_valid, GRAVID %in% c(1, NA))
  #remove participants who have used antibiotics in the last 6 months (BL_USE_RX_J01==1)
  gtdb_bac_data_valid <- subset_samples(gtdb_bac_data_valid, BL_USE_RX_J01 %in% c(0, NA))
  gtdb_arch_data_valid <- subset_samples(gtdb_arch_data_valid, BL_USE_RX_J01 %in% c(0, NA))
  #save the final objects
  saveRDS(gtdb_bac_data_valid, "Liver_analysis_antibiotics/gtdb_bac_data_valid.RDs")
  saveRDS(gtdb_arch_data_valid, "Liver_analysis_antibiotics/gtdb_arch_data_valid.RDs")
}

#fit the validation data to the main data with a modified philr_prepare function
philr_prepare_bac_valid <- function (phyloseq_obj) {
  #Retain only the taxa passing the filtering in the main bacterial data set (to gain matching node balances in philr)
  new_obj <- prune_taxa(taxa_names(gtdb_bac_data_tr), phyloseq_obj)
  #Add a pseudocount of 1 to the remaining taxa to avoid calculating log-rations involving zeros
  new_obj <- transform_sample_counts(new_obj, function(x) x+1)
  #Process the phylogenetic tree
  #Check that the tree is rooted and all multichotomies are resolved (the tree is binary)
  if (!is.rooted(phy_tree(new_obj))) {
    stop("Error: The tree is not rooted. Please root the tree in your phyloseq object.")
  } else if (!is.binary.tree(phy_tree(new_obj))) {
    stop("Error: There are unresolved multichotomies in the tree.")
  }
  #Rename the internal nodes so they are easier to work with.
  phy_tree(new_obj) <- makeNodeLabel(phy_tree(new_obj), method="number", prefix="n")
  #Check the root name balance of the tree and print it out.
  print(paste0("The root name balance of the tree is: ", name.balance(phy_tree(new_obj), tax_table(new_obj), "n1"), collapse=""))
  return(new_obj)
}

gtdb_bac_data_tr_valid <- philr_prepare_bac_valid(gtdb_bac_data_valid)
gtdb_bac_philr_valid <- philr_transform(gtdb_bac_data_tr_valid)
#Calculate the Fatty Liver Index.
sample_data(gtdb_bac_data_tr_valid)$FLI <- calculate_FLI(sample_data(gtdb_bac_data_tr_valid))

#plot FLI and its components in the validation data
p_TRIG_valid <- ggplot(data=sample_data(gtdb_bac_data_tr_valid), aes(x=TRIG)) + geom_histogram(bins=20, fill="gray50") + 
  scale_x_log10(breaks = scales::trans_breaks("log10", function(x) 10^x), labels = scales::trans_format("log10", scales::math_format(10^.x))) +
  scale_y_continuous(limits=c(0,50)) + 
  xlab("Triglycerides (mmol/L)") + ylab("Count") + theme(axis.text.x = element_text(size=14)) + ggtitle("B")
p_BMI_valid <- ggplot(data=sample_data(gtdb_bac_data_tr_valid), aes(x=BMI)) + geom_histogram(bins=20, fill="gray50") +
  scale_y_continuous(limits=c(0,50)) +
  xlab("BMI") + theme(axis.title.y = element_blank(), axis.text.y = element_blank(), axis.ticks.y = element_blank(), axis.text.x = element_text(size=14))
p_GGT_valid <- ggplot(data=sample_data(gtdb_bac_data_tr_valid), aes(x=GGT)) + geom_histogram(bins=20, fill="gray50") +
  scale_x_log10(breaks = scales::trans_breaks("log10", function(x) 10^x), labels = scales::trans_format("log10", scales::math_format(10^.x))) +
  scale_y_continuous(limits=c(0,50)) +
  xlab("GGT (units/L)") + theme(axis.title.y = element_blank(), axis.text.y = element_blank(), axis.ticks.y = element_blank(), axis.text.x = element_text(size=14))
p_WAIST_valid <- ggplot(data=sample_data(gtdb_bac_data_tr_valid), aes(x=VYOTARO)) + geom_histogram(bins=20, fill="gray50") +
  scale_y_continuous(limits=c(0,50)) + 
  xlab("Waist Circumference (cm)") + theme(axis.title.y = element_blank(), axis.text.y = element_blank(), axis.ticks.y = element_blank(), axis.text.x = element_text(size=14))
plot_FLI_valid <- data.frame(FLI = sample_data(gtdb_bac_data_tr_valid)$FLI)
plot_FLI_label_valid <- paste0("FLI = 60\n", round(length(which(plot_FLI_valid$FLI >= 60))/length(plot_FLI_valid$FLI)*100, 1), "%")
p_FLI_valid <- ggplot(data=plot_FLI_valid, aes(x=FLI)) + geom_histogram(breaks=seq(0,100, by=5), fill="gray50") + geom_vline(aes(xintercept = 60), linetype = "dashed", size = 0.5) + 
  annotate("text", x = 80, y = 40, label = plot_FLI_label_valid, size = 5) + xlab("Fatty Liver Index") + ylab("Count") + scale_x_continuous(breaks=c(-Inf, 0, 20, 40, 60, 80, 100, Inf)) +
  theme(axis.text.x = element_text(size=14)) + ggtitle("A")
p <- (p_FLI_valid | p_TRIG_valid | p_BMI_valid | p_GGT_valid | p_WAIST_valid)
ggsave(file = "Liver_analysis_antibiotics/FLI_boxplots_valid.svg", plot=p, units="cm", width=42, height=12)

#prepare validation data for model validation
FLI_ML_data_valid <- cbind(data.frame(sample_data(gtdb_bac_data_tr_valid)[,c("FLI", "MEN", "BL_AGE")]), as(otu_table(gtdb_bac_philr_valid, taxa_are_rows = FALSE), "matrix"))
#Replace zeros
FLI_ML_data_valid$FLI[is.na(FLI_ML_data_valid$FLI)] <- mean(FLI_ML_data_valid$FLI, na.rm = TRUE)
#subset validation data to the predictor balances in the main models
top_predictors_class <- readRDS("Liver_analysis_antibiotics/top_predictors_class.RDs")
top_predictors_class <- Reduce(intersect, top_predictors_class)
validation_data_class <- list(as.matrix(FLI_ML_data_valid[,colnames(FLI_ML_test) %in% c("FLI", top_predictors_class)]), as.matrix(FLI_ML_data_valid[,colnames(FLI_ML_data_valid) %in% c("FLI", c("MEN", "BL_AGE"))]), as.matrix(FLI_ML_data_valid[,colnames(FLI_ML_data_valid) %in% c("FLI", top_predictors_class[-which(top_predictors_class %in% c("MEN", "BL_AGE"))])]))
validation_stats_class_external <- list(NULL)
auc_curves <- list(NULL)
all_predictions <- data.frame(matrix(nrow=nrow(FLI_ML_data_valid), ncol=6))
for (fold in 1:length(cv_folds)) {
	validation_stats_sensset <- list(NULL)
    for (sensset in 1:3) {
		print(paste0("Regression model omitting region ", test_fold_region_codes[fold,1], " (", test_fold_region_codes[fold,2], "), including ", sens_codes[sensset], ", tested on validation data"))
		dvalid_class <- xgb.DMatrix(data = validation_data_class[[sensset]][,-1], label = ifelse(validation_data_class[[sensset]][,1] >= 60, 1, 0))
		fold_model_class <- xgb.load(paste0("Liver_analysis_antibiotics/fold_model_class", fold, "_", sensset, ".model"))
		predicted_class <- predict(fold_model_class, dvalid_class)
		aucs <- evalmod(scores=predicted_class, labels=getinfo(dvalid_class, "label"))
		auc_values <- c(as.numeric(attr(aucs, "aucs")$aucs), as.numeric(attr(aucs, "data_info")$np/(attr(aucs, "data_info")$np+attr(aucs, "data_info")$nn)))
		validation_stats_sensset[[sensset]] <- auc_values
		if (sensset == 1) {
			all_predictions[,fold] <- predicted_class
			auc_curves[[fold]] <- rbind(data.frame(fold = fold, type = "roc", x = aucs$rocs[[1]]$x, y = aucs$rocs[[1]]$y), data.frame(fold = fold, type = "prc", x = aucs$prcs[[1]]$x, y = aucs$prcs[[1]]$y))
		}
		print(paste0("Model AUC: ", round(auc_values[1], 4), " and AUPRC: ", round(auc_values[2], 4), " at baseline of ", round(auc_values[3], 2)*100, "% positive cases in ", dim(dvalid_class)[1], " participants, when extrapolating to validation data"))
	}
validation_stats_class_external[[fold]] <- as.data.frame(validation_stats_sensset, col.names=c(1:3))
}

validation_stats_class_df_external <- data.frame(CV_region=c(as.character(test_fold_region_codes[,2]), "Mean"), Main_model=round(c(unlist(lapply(validation_stats_class_external, `[[`, 1))[c(T,F,F)], mean(unlist(lapply(validation_stats_class_external, `[[`, 1))[c(T,F,F)])),3), Main_model_covariates=round(c(unlist(lapply(validation_stats_class_external, `[[`, 2))[c(T,F,F)], mean(unlist(lapply(validation_stats_class_external, `[[`, 2))[c(T,F,F)])),3), Main_model_balances=round(c(unlist(lapply(validation_stats_class_external, `[[`, 3))[c(T,F,F)], mean(unlist(lapply(validation_stats_class_external, `[[`, 3))[c(T,F,F)])),3), Main_model_test_set_N=rep(258, 7))
validation_stats_class_auprc_df_external <- data.frame(CV_region=c(as.character(test_fold_region_codes[,2]), "Mean"), Main_model=round(c(unlist(lapply(validation_stats_class_external, `[[`, 1))[c(F,T,F)], mean(unlist(lapply(validation_stats_class_external, `[[`, 1))[c(F,T,F)])),3), Main_model_covariates=round(c(unlist(lapply(validation_stats_class_external, `[[`, 2))[c(F,T,F)], mean(unlist(lapply(validation_stats_class_external, `[[`, 2))[c(F,T,F)])),3), Main_model_balances=round(c(unlist(lapply(validation_stats_class_external, `[[`, 3))[c(F,T,F)], mean(unlist(lapply(validation_stats_class_external, `[[`, 3))[c(F,T,F)])),3), Main_model_test_set_N=rep(258, 7), Main_model_test_set_positives=round(c(unlist(lapply(validation_stats_class_external, `[[`, 1))[c(F,F,T)], mean(unlist(lapply(validation_stats_class_external, `[[`, 1))[c(F,F,T)])),3))
write.table(validation_stats_class_df_external, "Liver_analysis_antibiotics/model_performance_auc_N_external.tsv", sep="\t", row.names=F)
write.table(validation_stats_class_auprc_df_external, "Liver_analysis_antibiotics/model_performance_auprc_N_external.tsv", sep="\t", row.names=F)

#Average predictions of the 6 LOGOCV models on the external validation data and calculate metrics
ensemble_aucs <- evalmod(scores=rowMeans(all_predictions), labels=ifelse(validation_data_class[[1]][,1] >= 60, 1, 0))
ensemble_auc_values <- c(as.numeric(attr(ensemble_aucs, "aucs")$aucs), as.numeric(attr(ensemble_aucs, "data_info")$np/(attr(ensemble_aucs, "data_info")$np+attr(ensemble_aucs, "data_info")$nn)))

#Plot receiver-operator and precision-recall curves
#Single curves based on averaged predictions of the 6 models
svg("Liver_analysis_antibiotics/curves.svg")
autoplot(ensemble_aucs)
dev.off()
#Alternatively, plot curves for all models separately
curves_df <- do.call("rbind", auc_curves)
image <- ggplot(data=curves_df, aes(x=x, y=y, group=fold)) + geom_line(alpha=0.25, color="black") + scale_y_continuous(limits=c(0.0,1.0), breaks=seq(0.0,1.0,0.2)) +
  theme(axis.title.y=element_blank()) + facet_wrap(.~type)
ggsave(file = "Liver_analysis_antibiotics/curves_alt.svg", plot=image, units="cm", width=25, height=10)

#Comparison with NCBI data from SHOGUN
if (file.exists("Liver_analysis_antibiotics/ncbi_bac_data.RDs") && file.exists("Liver_analysis_antibiotics/ncbi_arch_data.RDs")) {
  ncbi_bac_data <- readRDS("Liver_analysis_antibiotics/ncbi_bac_data.RDs")
  ncbi_arch_data <- readRDS("Liver_analysis_antibiotics/ncbi_arch_data.RDs")
} else {
  #Construct the primary phyloseq object and subset to FR02 samples.
  ncbi_data <- biomformat::read_biom("SHOGUN.species.biom") #BIOM table from the SHOGUN output
  ncbi_data <- biomformat::biom_data(ncbi_data)
  ncbi_tax_table <- strsplit(row.names(as.matrix(ncbi_data)), ";")
  ncbi_tax_table <- matrix(unlist(ncbi_tax_table), nrow=length(ncbi_tax_table), byrow=T)
  row.names(ncbi_data) <- row.names(ncbi_tax_table)
  ncbi_data <- phyloseq(otu_table(as.matrix(ncbi_data), taxa_are_rows=T), tax_table(ncbi_tax_table))
  #subset NCBI data to the same samples with GTDB data (all QC already performed)
  ncbi_data <- prune_samples(sample_names(gtdb_bac_data), ncbi_data)
  #Format the tax table.
  colnames(tax_table(ncbi_data)) <- c("Domain", "Phylum", "Class", "Order", "Family",  "Genus", "Species")
  #Subset to archaeal and bacterial sets
  ncbi_bac_data <- subset_taxa(ncbi_data, Domain=="k__Bacteria")
  ncbi_arch_data <- subset_taxa(ncbi_data, Domain=="k__Archaea")
  #Combine the phenotype data with the bacterial and archaeal data
  ncbi_bac_data <- phyloseq(otu_table(ncbi_bac_data), tax_table(ncbi_bac_data), sample_data(FR02))
  ncbi_arch_data <- phyloseq(otu_table(ncbi_arch_data), tax_table(ncbi_arch_data), sample_data(FR02))
  #save the final objects
  saveRDS(ncbi_bac_data, "Liver_analysis_antibiotics/ncbi_bac_data.RDs")
  saveRDS(ncbi_arch_data, "Liver_analysis_antibiotics/ncbi_arch_data.RDs")
}

#Plot Phylum-level community compositions as bars
phyla_bac_FR02 <- phyloseq(otu_table(gtdb_bac_data), tax_table(gtdb_bac_data), sample_data(gtdb_bac_data_tr))
phyla_arch_FR02 <- phyloseq(otu_table(gtdb_arch_data), tax_table(gtdb_arch_data), sample_data(gtdb_arch_data_tr))
phyla_bac_FR07 <- phyloseq(otu_table(gtdb_bac_data_valid), tax_table(gtdb_bac_data_valid), sample_data(gtdb_bac_data_tr_valid))
phyla_arch_FR07 <- phyloseq(otu_table(gtdb_arch_data_valid), tax_table(gtdb_arch_data_valid), sample_data(gtdb_bac_data_tr_valid))
phyla_bac_ncbi <- phyloseq(otu_table(ncbi_bac_data), tax_table(ncbi_bac_data), sample_data(gtdb_bac_data_tr))
phyla_arch_ncbi <- phyloseq(otu_table(ncbi_arch_data), tax_table(ncbi_arch_data), sample_data(gtdb_arch_data_tr))
phyla_bac_FR02 <- tax_glom(phyla_bac_FR02, "Phylum")
phyla_arch_FR02 <- tax_glom(phyla_arch_FR02, "Phylum")
phyla_bac_FR07 <- tax_glom(phyla_bac_FR07, "Phylum")
phyla_arch_FR07 <- tax_glom(phyla_arch_FR07, "Phylum")
phyla_bac_ncbi <- tax_glom(phyla_bac_ncbi, "Phylum")
phyla_arch_ncbi <- tax_glom(phyla_arch_ncbi, "Phylum")
least_abundant_phyla_bac_FR02 <- names(taxa_sums(phyla_bac_FR02))[taxa_sums(phyla_bac_FR02)/sum(taxa_sums(phyla_bac_FR02))*100 < 1]
least_abundant_phyla_arch_FR02 <- names(taxa_sums(phyla_arch_FR02))[taxa_sums(phyla_arch_FR02)/sum(taxa_sums(phyla_arch_FR02))*100 < 1]
least_abundant_phyla_bac_FR07 <- names(taxa_sums(phyla_bac_FR07))[taxa_sums(phyla_bac_FR07)/sum(taxa_sums(phyla_bac_FR07))*100 < 1]
least_abundant_phyla_arch_FR07 <- names(taxa_sums(phyla_arch_FR07))[taxa_sums(phyla_arch_FR07)/sum(taxa_sums(phyla_arch_FR07))*100 < 1]
least_abundant_phyla_bac_ncbi <- names(taxa_sums(phyla_bac_ncbi))[taxa_sums(phyla_bac_ncbi)/sum(taxa_sums(phyla_bac_ncbi))*100 < 1]
least_abundant_phyla_arch_ncbi <- names(taxa_sums(phyla_arch_ncbi))[taxa_sums(phyla_arch_ncbi)/sum(taxa_sums(phyla_arch_ncbi))*100 < 1]
phyla_bac_FR02 <- merge_taxa(phyla_bac_FR02, least_abundant_phyla_bac_FR02, 1)
phyla_arch_FR02 <- merge_taxa(phyla_arch_FR02, least_abundant_phyla_arch_FR02, 1)
phyla_bac_FR07 <- merge_taxa(phyla_bac_FR07, least_abundant_phyla_bac_FR07, 1)
phyla_arch_FR07 <- merge_taxa(phyla_arch_FR07, least_abundant_phyla_arch_FR07, 1)
phyla_bac_ncbi <- merge_taxa(phyla_bac_ncbi, least_abundant_phyla_bac_ncbi, 1)
phyla_arch_ncbi <- merge_taxa(phyla_arch_ncbi, least_abundant_phyla_arch_ncbi, 1)
tax_table(phyla_bac_FR02)[tax_table(phyla_bac_FR02)[,2] %in% NA,2] <- "p__Other_Bacteria"
tax_table(phyla_arch_FR02)[tax_table(phyla_arch_FR02)[,2] %in% NA,2] <- "p__Other_Archaea"
tax_table(phyla_bac_FR07)[tax_table(phyla_bac_FR07)[,2] %in% NA,2] <- "p__Other_Bacteria"
tax_table(phyla_arch_FR07)[tax_table(phyla_arch_FR07)[,2] %in% NA,2] <- "p__Other_Archaea"
tax_table(phyla_bac_ncbi)[tax_table(phyla_bac_ncbi)[,2] %in% NA,2] <- "p__Other_Bacteria"
tax_table(phyla_arch_ncbi)[tax_table(phyla_arch_ncbi)[,2] %in% NA,2] <- "p__Other_Archaea"
#substitute NA values in FLI with mean and group into low and high FLI groups
sample_data(phyla_bac_FR02)$FLI[is.na(sample_data(phyla_bac_FR02)$FLI)] <- mean(sample_data(phyla_bac_FR02)$FLI, na.rm = TRUE)
sample_data(phyla_bac_FR02)$FLI_group <- ifelse(sample_data(phyla_bac_FR02)$FLI < 60, "FLI < 60", "FLI ≥ 60")
sample_data(phyla_arch_FR02)$FLI[is.na(sample_data(phyla_arch_FR02)$FLI)] <- mean(sample_data(phyla_arch_FR02)$FLI, na.rm = TRUE)
sample_data(phyla_arch_FR02)$FLI_group <- ifelse(sample_data(phyla_arch_FR02)$FLI < 60, "FLI < 60", "FLI ≥ 60")
sample_data(phyla_bac_FR07)$FLI[is.na(sample_data(phyla_bac_FR07)$FLI)] <- mean(sample_data(phyla_bac_FR07)$FLI, na.rm = TRUE)
sample_data(phyla_bac_FR07)$FLI_group <- ifelse(sample_data(phyla_bac_FR07)$FLI < 60, "FLI < 60", "FLI ≥ 60")
sample_data(phyla_arch_FR07)$FLI[is.na(sample_data(phyla_arch_FR07)$FLI)] <- mean(sample_data(phyla_arch_FR07)$FLI, na.rm = TRUE)
sample_data(phyla_arch_FR07)$FLI_group <- ifelse(sample_data(phyla_arch_FR07)$FLI < 60, "FLI < 60", "FLI ≥ 60")
sample_data(phyla_bac_ncbi)$FLI[is.na(sample_data(phyla_bac_ncbi)$FLI)] <- mean(sample_data(phyla_bac_ncbi)$FLI, na.rm = TRUE)
sample_data(phyla_bac_ncbi)$FLI_group <- ifelse(sample_data(phyla_bac_ncbi)$FLI < 60, "FLI < 60", "FLI ≥ 60")
sample_data(phyla_arch_ncbi)$FLI[is.na(sample_data(phyla_arch_ncbi)$FLI)] <- mean(sample_data(phyla_arch_ncbi)$FLI, na.rm = TRUE)
sample_data(phyla_arch_ncbi)$FLI_group <- ifelse(sample_data(phyla_arch_ncbi)$FLI < 60, "FLI < 60", "FLI ≥ 60")
#scale counts per total sum per sample and group by FLI
phyla_bac_FR02 <- transform_sample_counts(phyla_bac_FR02, function(x) 100 * x/sum(x))
phyla_arch_FR02 <- transform_sample_counts(phyla_arch_FR02, function(x) 100 * x/sum(x))
phyla_bac_FR07 <- transform_sample_counts(phyla_bac_FR07, function(x) 100 * x/sum(x))
phyla_arch_FR07 <- transform_sample_counts(phyla_arch_FR07, function(x) 100 * x/sum(x))
phyla_bac_ncbi <- transform_sample_counts(phyla_bac_ncbi, function(x) 100 * x/sum(x))
phyla_arch_ncbi <- transform_sample_counts(phyla_arch_ncbi, function(x) 100 * x/sum(x))
bars_bac_FR02 <- data.frame(t(otu_table(phyla_bac_FR02)))
bars_arch_FR02 <- data.frame(t(otu_table(phyla_arch_FR02)))
bars_bac_FR07 <- data.frame(t(otu_table(phyla_bac_FR07)))
bars_arch_FR07 <- data.frame(t(otu_table(phyla_arch_FR07)))
bars_bac_ncbi <- data.frame(t(otu_table(phyla_bac_ncbi)))
bars_arch_ncbi <- data.frame(t(otu_table(phyla_arch_ncbi)))
colnames(bars_bac_FR02) <- c(tax_table(phyla_bac_FR02)[,2])
colnames(bars_arch_FR02) <- c(tax_table(phyla_arch_FR02)[,2])
colnames(bars_bac_FR07) <- c(tax_table(phyla_bac_FR07)[,2])
colnames(bars_arch_FR07) <- c(tax_table(phyla_arch_FR07)[,2])
colnames(bars_bac_ncbi) <- c(tax_table(phyla_bac_ncbi)[,2])
colnames(bars_arch_ncbi) <- c(tax_table(phyla_arch_ncbi)[,2])
bars_bac_FR02$FLI_group <- sample_data(phyla_bac_FR02)$FLI_group
bars_arch_FR02$FLI_group <- sample_data(phyla_arch_FR02)$FLI_group
bars_bac_FR07$FLI_group <- sample_data(phyla_bac_FR07)$FLI_group
bars_arch_FR07$FLI_group <- sample_data(phyla_arch_FR07)$FLI_group
bars_bac_ncbi$FLI_group <- sample_data(phyla_bac_ncbi)$FLI_group
bars_arch_ncbi$FLI_group <- sample_data(phyla_arch_ncbi)$FLI_group
bars_bac_FR02$type <- "Bacteria"
bars_bac_FR02$set <- "FINRISK 2002"
bars_arch_FR02$type <- "Archaea"
bars_arch_FR02$set <- "FINRISK 2002"
bars_bac_FR07$type <- "Bacteria"
bars_bac_FR07$set <- "FINRISK 2007"
bars_arch_FR07$type <- "Archaea"
bars_arch_FR07$set <- "FINRISK 2007"
bars_bac_ncbi$type <- "Bacteria"
bars_bac_ncbi$set <- "FINRISK 2002 NCBI"
bars_arch_ncbi$type <- "Archaea"
bars_arch_ncbi$set <- "FINRISK 2002 NCBI"
bars_all <- dplyr::bind_rows(bars_bac_FR02, bars_arch_FR02, bars_bac_FR07, bars_arch_FR07)
bars_all_melt <- reshape2::melt(bars_all)
bars_all_melt <- aggregate(bars_all_melt$value, by=list(FLI_group = bars_all_melt$FLI_group, set = bars_all_melt$set, type = bars_all_melt$type, variable = bars_all_melt$variable), function (x) c(mean = mean(x, na.rm=T), sem = sqrt(var(x)/length(x))))
bars_all_melt$variable <- factor(bars_all_melt$variable, levels=c("p__Firmicutes_A", "p__Firmicutes_C", "p__Firmicutes", "p__Bacteroidota", "p__Actinobacteriota", "p__Proteobacteria", "p__Other_Bacteria", "p__Halobacterota", "p__Euryarchaeota", "p__Thermoplasmatota", "p__Crenarchaeota", "p__Nanoarchaeota", "p__Asgardarchaeota", "p__Altiarchaeota", "p__Other_Archaea"))
#plot
phylum_palette <- c("#be8d60","#663d0e","#aa6e00","#b2a600","#817d00","#bdbd4d","gray","#4a8f00","#9dc377","#445b3a","#006b1c","#006d3a","#6ac9ad","#018e86","gray")
image <- ggplot(bars_all_melt, aes(x=factor(variable), y=x[,1], fill=factor(variable))) + scale_x_discrete(limits=rev(levels(bars_all_melt$variable))) + scale_y_continuous(limits=c(0,50)) + geom_bar(stat="identity") + 
  scale_fill_manual(values = phylum_palette, name = "Phylum") + 
  theme(axis.text.y = element_text(hjust = 0, size = 22), strip.text.x = element_text(size = 20), axis.ticks = element_blank(), panel.border = element_blank(), axis.title.y = element_blank(), legend.position="none", plot.title = element_text(size = 28, hjust = 0.5), panel.grid.major.x = element_line(colour="gray", size=0.5)) + facet_wrap(set~FLI_group, scales = "free_x") + 
  xlab("Sample") + ylab("Abundance (%)") + coord_flip()
ggsave(file = "Liver_analysis_antibiotics/phyla_bars_FLI_groups.svg", plot=image, units="cm", width=42, height=42)

bars_ncbi <- dplyr::bind_rows(bars_bac_ncbi, bars_arch_ncbi)
bars_ncbi_melt <- reshape2::melt(bars_ncbi)
bars_ncbi_melt <- aggregate(bars_ncbi_melt$value, by=list(FLI_group = bars_ncbi_melt$FLI_group, set = bars_ncbi_melt$set, type = bars_ncbi_melt$type, variable = bars_ncbi_melt$variable), function (x) c(mean = mean(x, na.rm=T), sem = sqrt(var(x)/length(x))))
bars_ncbi_melt$variable <- factor(bars_ncbi_melt$variable, levels=c("p__Firmicutes", "p__Bacteroidetes", "p__Actinobacteria", "p__Proteobacteria", "p__Other_Bacteria", "p__Euryarchaeota", "p__Other_Archaea"))
#plot
phylum_ncbi_palette <- c("#aa6e00","#b2a600","#817d00","#bdbd4d","gray","#9dc377","gray")
image <- ggplot(bars_ncbi_melt, aes(x=factor(variable), y=x[,1], fill=factor(variable))) + scale_x_discrete(limits=rev(levels(bars_ncbi_melt$variable))) + scale_y_continuous(limits=c(0,100)) + geom_bar(stat="identity") + 
  scale_fill_manual(values = phylum_ncbi_palette, name = "Phylum") + 
  theme(axis.text.y = element_text(hjust = 0, size = 22), strip.text.x = element_text(size = 20), axis.ticks = element_blank(), panel.border = element_blank(), axis.title.y = element_blank(), legend.position="none", plot.title = element_text(size = 28, hjust = 0.5), panel.grid.major.x = element_line(colour="gray", size=0.5)) + facet_wrap(set~FLI_group, scales = "free_x") + 
  xlab("Sample") + ylab("Abundance (%)") + coord_flip()
ggsave(file = "Liver_analysis_antibiotics/phyla_bars_FLI_groups_ncbi.svg", plot=image, units="cm", width=42, height=21)

#Check connections between SHOGUN/NCBI analyzed data and high FLI
otu_table_ncbi <- as.data.frame(as(otu_table(ncbi_bac_data), "matrix"))
#mean substitute missing FLI values and convert to FLI groups
FLI_cat <- sample_data(gtdb_bac_data_tr)$FLI
FLI_cat[is.na(FLI_cat)] <- mean(FLI_cat, na.rm = TRUE)
FLI_cat <- ifelse(FLI_cat < 60, "low_FLI60", "high_FLI60")
FLI_cat <- factor(FLI_cat, levels = c("low_FLI60", "high_FLI60"))
#format covariates
FLI_covariates <- data.frame(FLI = sample_data(gtdb_bac_data_tr)$FLI, FLI_cat = FLI_cat, MEN = as.factor(sample_data(gtdb_bac_data_tr)$MEN), BL_AGE = as.numeric(sample_data(gtdb_bac_data_tr)$BL_AGE), ALKI2_FR02 = as.numeric(sample_data(gtdb_bac_data_tr)$ALKI2_FR02))

#Calculate alpha-diversity for each sample from 10 random rarefactions and plot and test differences between FLI groups
#Do this for both the GTDB and NCBI data sets
gtdb_avgrichness <- list(NULL)
for (j in 1:10) {
	set.seed(42*j)
	rarefied <- rarefy_even_depth(gtdb_bac_data)
	richness <- estimate_richness(rarefied)
	Goods.cover <- NULL
	for (k in 1:nsamples(rarefied)) {
		Goods.cover[k] <- 1-(length(which(otu_table(rarefied)[,k] == 1))/sample_sums(rarefied)[k])
	}
	richness <- cbind(richness, Goods.cover=Goods.cover)
	richness$sample <- sample_names(gtdb_bac_data)
	rownames(richness) <- seq(1+nrow(richness)*(j-1),nrow(richness)*j)
	gtdb_avgrichness[[j]] <- richness
}
gtdb_richness <- do.call("rbind",gtdb_avgrichness)
gtdb_richness_chao <- aggregate(Chao1 ~ sample, gtdb_richness, mean)
gtdb_richness_chao <- cbind(gtdb_richness_chao, aggregate(Chao1 ~ sample, gtdb_richness, sd)[2])
gtdb_richness_shannon <- aggregate(Shannon ~ sample, gtdb_richness, mean)[2]
gtdb_richness_shannon <- cbind(gtdb_richness_shannon, aggregate(Shannon ~ sample, gtdb_richness, sd)[2])
gtdb_richness_InvSimpson <- aggregate(InvSimpson ~ sample, gtdb_richness, mean)[2]
gtdb_richness_InvSimpson <- cbind(gtdb_richness_InvSimpson, aggregate(InvSimpson ~ sample, gtdb_richness, sd)[2])
gtdb_richness_Goodscover <- aggregate(Goods.cover ~ sample, gtdb_richness, mean)[2]
gtdb_richness_Goodscover <- cbind(gtdb_richness_Goodscover, aggregate(Goods.cover ~ sample, gtdb_richness, sd)[2])
gtdb_richness2 <- cbind(gtdb_richness_chao, gtdb_richness_shannon, gtdb_richness_InvSimpson, gtdb_richness_Goodscover, FLI_covariates)
colnames(gtdb_richness2) <- c("sample", "Chao1", "Chao1.sd", "Shannon", "Shannon.sd", "InvSimpson", "InvSimpson.sd", "Goods.cover", "Goods.cover.sd", colnames(FLI_covariates))
gtdb_richness2$FLI_cat <- factor(gtdb_richness2$FLI_cat, levels = c("low_FLI60", "high_FLI60"))
#Run a logit model, while adjusting for covariates
gtdb_alpha_diversity_model <- glm(FLI_cat ~ Shannon + MEN + BL_AGE + ALKI2_FR02, data = gtdb_richness2, family = "binomial")
summary(gtdb_alpha_diversity_model)
#Calculate effect size for a-diversity as %
print(paste0("High FLI group Shannon mean is ", round(coef(gtdb_alpha_diversity_model)["Shannon"]/aggregate(gtdb_richness2$Shannon ~ FLI_cat, gtdb_richness2, mean)[1,2]*100, 2), " +-", round(coef(summary(gtdb_alpha_diversity_model))[2,2]/aggregate(gtdb_richness2$Shannon ~ FLI_cat, gtdb_richness2, mean)[1,2]*100, 2), " % of low group FLI Shannon mean, when analyzing Centrifuge/GTDB data."))

ncbi_avgrichness <- list(NULL)
for (j in 1:10) {
	set.seed(42*j)
	rarefied <- rarefy_even_depth(ncbi_bac_data)
	richness <- estimate_richness(rarefied)
	Goods.cover <- NULL
	for (k in 1:nsamples(rarefied)) {
		Goods.cover[k] <- 1-(length(which(otu_table(rarefied)[,k] == 1))/sample_sums(rarefied)[k])
	}
	richness <- cbind(richness, Goods.cover=Goods.cover)
	richness$sample <- sample_names(ncbi_bac_data)
	rownames(richness) <- seq(1+nrow(richness)*(j-1),nrow(richness)*j)
	ncbi_avgrichness[[j]] <- richness
}
ncbi_richness <- do.call("rbind",ncbi_avgrichness)
ncbi_richness_chao <- aggregate(Chao1 ~ sample, ncbi_richness, mean)
ncbi_richness_chao <- cbind(ncbi_richness_chao, aggregate(Chao1 ~ sample, ncbi_richness, sd)[2])
ncbi_richness_shannon <- aggregate(Shannon ~ sample, ncbi_richness, mean)[2]
ncbi_richness_shannon <- cbind(ncbi_richness_shannon, aggregate(Shannon ~ sample, ncbi_richness, sd)[2])
ncbi_richness_InvSimpson <- aggregate(InvSimpson ~ sample, ncbi_richness, mean)[2]
ncbi_richness_InvSimpson <- cbind(ncbi_richness_InvSimpson, aggregate(InvSimpson ~ sample, ncbi_richness, sd)[2])
ncbi_richness_Goodscover <- aggregate(Goods.cover ~ sample, ncbi_richness, mean)[2]
ncbi_richness_Goodscover <- cbind(ncbi_richness_Goodscover, aggregate(Goods.cover ~ sample, ncbi_richness, sd)[2])
ncbi_richness2 <- cbind(ncbi_richness_chao, ncbi_richness_shannon, ncbi_richness_InvSimpson, ncbi_richness_Goodscover, FLI_covariates)
colnames(ncbi_richness2) <- c("sample", "Chao1", "Chao1.sd", "Shannon", "Shannon.sd", "InvSimpson", "InvSimpson.sd", "Goods.cover", "Goods.cover.sd", colnames(FLI_covariates))
ncbi_richness2$FLI_cat <- factor(ncbi_richness2$FLI_cat, levels = c("low_FLI60", "high_FLI60"))
#Run a logit model, while adjusting for covariates
ncbi_alpha_diversity_model <- glm(FLI_cat ~ Shannon + MEN + BL_AGE + ALKI2_FR02, data = ncbi_richness2, family = "binomial")
summary(ncbi_alpha_diversity_model)
#Calculate effect size for a-diversity as %
print(paste0("High FLI group Shannon mean is ", round(coef(ncbi_alpha_diversity_model)["Shannon"]/aggregate(ncbi_richness2$Shannon ~ FLI_cat, ncbi_richness2, mean)[1,2]*100, 2), " +-", round(coef(summary(ncbi_alpha_diversity_model))[2,2]/aggregate(ncbi_richness2$Shannon ~ FLI_cat, ncbi_richness2, mean)[1,2]*100, 2), " % of low group FLI Shannon mean, when analyzing SHOGUN/NCBI data."))


#Calculate beta-diversity (between-samples) from CLR + PCA
ncbi_bac_CLR <- microbiome::transform(ncbi_bac_data, 'clr')
ncbi_bac_pca <- rda(as.matrix(t(otu_table(ncbi_bac_CLR))))
ncbi_PCA_lm_bac_data <- cbind(summary(ncbi_bac_pca)$sites, FLI_covariates)
colnames(ncbi_PCA_lm_bac_data)[1:3] <- c("PC1_b", "PC2_b", "PC3_b") 
ncbi_FLI_lm_model <- lm(log10(FLI) ~ PC1_b + PC2_b + PC3_b + MEN + BL_AGE + ALKI2_FR02, data=ncbi_PCA_lm_bac_data)
summary(ncbi_FLI_lm_model)

#Plot beta-diversity comparison of NCBI to GTDB
#invert PC1 in NCBI data to match the direction of the associations in GTDB
ncbi_invPC1 <- ncbi_PCA_lm_bac_data[c("PC1_b", "PC2_b", "PC3_b")]
ncbi_invPC1$PC1_b <- -1*ncbi_invPC1$PC1_b
#plot rest normally
ncbi_plot_lm_data2 <- data.frame(ncbi_PCA_lm_bac_data["FLI"], lapply(ncbi_invPC1, function(x) quantcut(x=x, q=4, na.rm=T, labels=c("0-.25", ".25-.5", ".5-.75", ".75-1")))) 
ncbi_plot_lm_data2 <- reshape2::melt(ncbi_plot_lm_data2, measure.vars=c("PC1_b", "PC2_b", "PC3_b"))
ncbi_plot_lm_data2$value <- factor(ncbi_plot_lm_data2$value, levels=c("0-.25", ".25-.5", ".5-.75", ".75-1"))
p_gtdb_PC_boxplots <- ggplot(data=plot_lm_data2, aes(value, FLI)) +
  geom_boxplot() +
  geom_jitter(alpha=0.1, size=0.5) +
  geom_hline(yintercept=60, alpha=0.75, linetype="dashed") +
  facet_wrap(~variable, labeller=labeller(variable=PC_labs)) +
  ylab("Fatty Liver Index") +
  xlab("PC quantiles") +
  theme(axis.text.x = element_text(size=14), strip.text.x = element_text(size=20)) +
  ggtitle("A")
p_ncbi_PC_boxplots <- ggplot(data=ncbi_plot_lm_data2, aes(value, FLI)) +
  geom_boxplot() +
  geom_jitter(alpha=0.1, size=0.5) +
  geom_hline(yintercept=60, alpha=0.75, linetype="dashed") +
  facet_wrap(~variable, labeller=labeller(variable=PC_labs)) +
  ylab("Fatty Liver Index") +
  xlab("PC quantiles") +
  theme(axis.text.x = element_text(size=14), strip.text.x = element_text(size=20)) +
  ggtitle("B")
p <- p_gtdb_PC_boxplots / p_ncbi_PC_boxplots
ggsave(file = "Liver_analysis_antibiotics/FLI_boxplot_comparison.svg", plot=p, units="cm", width=42, height=24)

#run ALDEx2 models for both GTDB and NCBI data (https://github.com/ggloor/CoDa_microbiome_tutorial/wiki/Part-2:-ALDEx2-for-differential-expression-analysis)
alde_taxa_FLI_group <- aldex.clr(otu_table_ncbi, FLI_cat, mc.samples=128, verbose=TRUE)
alde_taxa_FLI_group.tt <- aldex.ttest(alde_taxa_FLI_group, FLI_cat, paired.test=FALSE)
otu_table_ncbi_sig <- otu_table_ncbi[which(rownames(otu_table_ncbi) %in% rownames(alde_taxa_FLI_group.tt)[which(alde_taxa_FLI_group.tt$we.eBH < 0.001)]),]
#reanalyze only taxa with p < 0.001 in the full model (to reduce computational effort)
alde_taxa_FLI_group_sig <- aldex.clr(otu_table_ncbi_sig, FLI_cat, mc.samples=128, verbose=TRUE)
alde_taxa_FLI_group_sig.tt <- aldex.ttest(alde_taxa_FLI_group_sig, FLI_cat, paired.test=FALSE)
alde_taxa_FLI_group_sig.effect <- aldex.effect(alde_taxa_FLI_group_sig, FLI_cat, include.sample.summary=FALSE, verbose=TRUE, useMC=TRUE)
alde_taxa_FLI_group_sig.all <- data.frame(alde_taxa_FLI_group_sig.tt, alde_taxa_FLI_group_sig.effect, stringsAsFactors=FALSE)
alde_taxa_FLI_group_sig.all2 <- alde_taxa_FLI_group_sig.all[c("rab.win.high_FLI60", "rab.win.low_FLI60", "diff.btw", "effect", "effect.low", "effect.high", "overlap")]
alde_taxa_FLI_group_sig.all2$BH_p <- "<0.001"
alde_taxa_FLI_group_sig.all2 <- alde_taxa_FLI_group_sig.all2 %>% 
 mutate_if(is.numeric, round, digits=3)
rownames(alde_taxa_FLI_group_sig.all2) <- rownames(alde_taxa_FLI_group_sig.all)
ncbi_taxa_FLI_sig_neg <- rownames(alde_taxa_FLI_group_sig.all2)[which(alde_taxa_FLI_group_sig.all2$effect < 0)]
ncbi_taxa_FLI_sig_pos <- rownames(alde_taxa_FLI_group_sig.all2)[which(alde_taxa_FLI_group_sig.all2$effect > 0)]
ncbi_bac_data_sig_neg <- tax_table(ncbi_bac_data)[which(taxa_names(ncbi_bac_data) %in% ncbi_taxa_FLI_sig_neg),]
ncbi_bac_data_sig_neg <- cbind(ncbi_bac_data_sig_neg, alde_taxa_FLI_group_sig.all2[match(rownames(ncbi_bac_data_sig_neg), rownames(alde_taxa_FLI_group_sig.all2)),])
ncbi_bac_data_sig_neg <- ncbi_bac_data_sig_neg[order(ncbi_bac_data_sig_neg$effect),]
write.table(ncbi_bac_data_sig_neg, "Liver_analysis_antibiotics/ncbi_neg_taxa.tsv", sep="\t", row.names=FALSE)
ncbi_bac_data_sig_pos <- tax_table(ncbi_bac_data)[which(taxa_names(ncbi_bac_data) %in% ncbi_taxa_FLI_sig_pos),]
ncbi_bac_data_sig_pos <- cbind(ncbi_bac_data_sig_pos, alde_taxa_FLI_group_sig.all2[match(rownames(ncbi_bac_data_sig_pos), rownames(alde_taxa_FLI_group_sig.all2)),])
ncbi_bac_data_sig_pos <- ncbi_bac_data_sig_pos[order(-ncbi_bac_data_sig_pos$effect),]
write.table(ncbi_bac_data_sig_pos, "Liver_analysis_antibiotics/ncbi_pos_taxa.tsv", sep="\t", row.names=FALSE)

otu_table_gtdb <- as.data.frame(as(otu_table(gtdb_bac_data), "matrix"))
alde_taxa_FLI_group <- aldex.clr(otu_table_gtdb, FLI_cat, mc.samples=32, verbose=TRUE)
alde_taxa_FLI_group.tt <- aldex.ttest(alde_taxa_FLI_group, FLI_cat, paired.test=FALSE)
otu_table_gtdb_sig <- otu_table_gtdb[which(rownames(otu_table_gtdb) %in% rownames(alde_taxa_FLI_group.tt)[which(alde_taxa_FLI_group.tt$we.eBH < 0.001)]),]
#reanalyze only taxa with p < 0.001 in the full model (to reduce computational effort)
gtdb_alde_taxa_FLI_group_sig <- aldex.clr(otu_table_gtdb_sig, FLI_cat, mc.samples=32, verbose=TRUE)
gtdb_alde_taxa_FLI_group_sig.tt <- aldex.ttest(gtdb_alde_taxa_FLI_group_sig, FLI_cat, paired.test=FALSE)
gtdb_alde_taxa_FLI_group_sig.effect <- aldex.effect(gtdb_alde_taxa_FLI_group_sig, FLI_cat, include.sample.summary=FALSE, verbose=TRUE, useMC=TRUE)
gtdb_alde_taxa_FLI_group_sig.all <- data.frame(gtdb_alde_taxa_FLI_group_sig.tt, gtdb_alde_taxa_FLI_group_sig.effect, stringsAsFactors=FALSE)
gtdb_alde_taxa_FLI_group_sig.all2 <- gtdb_alde_taxa_FLI_group_sig.all[c("rab.win.high_FLI60", "rab.win.low_FLI60", "diff.btw", "effect", "effect.low", "effect.high", "overlap")]
gtdb_alde_taxa_FLI_group_sig.all2$BH_p <- "<0.001"
gtdb_alde_taxa_FLI_group_sig.all2 <- gtdb_alde_taxa_FLI_group_sig.all2 %>% 
 mutate_if(is.numeric, round, digits=3)
rownames(gtdb_alde_taxa_FLI_group_sig.all2) <- rownames(gtdb_alde_taxa_FLI_group_sig.all)
gtdb_taxa_FLI_sig_neg <- rownames(gtdb_alde_taxa_FLI_group_sig.all2)[which(gtdb_alde_taxa_FLI_group_sig.all2$effect < 0)]
gtdb_taxa_FLI_sig_pos <- rownames(gtdb_alde_taxa_FLI_group_sig.all2)[which(gtdb_alde_taxa_FLI_group_sig.all2$effect > 0)]
gtdb_bac_data_sig_neg <- tax_table(gtdb_bac_data)[which(taxa_names(gtdb_bac_data) %in% gtdb_taxa_FLI_sig_neg),]
gtdb_bac_data_sig_neg <- cbind(gtdb_bac_data_sig_neg, gtdb_alde_taxa_FLI_group_sig.all2[match(rownames(gtdb_bac_data_sig_neg), rownames(gtdb_alde_taxa_FLI_group_sig.all2)),])
gtdb_bac_data_sig_neg <- gtdb_bac_data_sig_neg[order(gtdb_bac_data_sig_neg$effect),]
write.table(gtdb_bac_data_sig_neg, "Liver_analysis_antibiotics/gtdb_neg_taxa.tsv", sep="\t", row.names=FALSE)
gtdb_bac_data_sig_pos <- tax_table(gtdb_bac_data)[which(taxa_names(gtdb_bac_data) %in% gtdb_taxa_FLI_sig_pos),]
gtdb_bac_data_sig_pos <- cbind(gtdb_bac_data_sig_pos, gtdb_alde_taxa_FLI_group_sig.all2[match(rownames(gtdb_bac_data_sig_pos), rownames(gtdb_alde_taxa_FLI_group_sig.all2)),])
gtdb_bac_data_sig_pos <- gtdb_bac_data_sig_pos[order(-gtdb_bac_data_sig_pos$effect),]
write.table(gtdb_bac_data_sig_pos, "Liver_analysis_antibiotics/gtdb_pos_taxa.tsv", sep="\t", row.names=FALSE)

save.image("Liver_analysis_antibiotics/Liver_analysis_antibiotics_notebook.RData") #Save the data
```
