## Supplementary material for "Links between gut microbiome composition and fatty liver disease in a large population sample": Ruuskanen_FLI_microbiome_SI.docx

**Supplementary figures and tables**

Matti O. Ruuskanen^1,2^*, Fredrik Åberg^3,4^, Ville Männistö^5,6^, Aki S. Havulinna^2,7^, Guillaume Méric^8,9^, Yang Liu^8,10^, Rohit Loomba^11,12^, Yoshiki Vázquez-Baeza^13,14^, Anupriya Tripathi^15,16,17^, Liisa M. Valsta^2^, Michael Inouye^8,18^, Pekka Jousilahti^2^, Veikko Salomaa^2^, Mohit Jain^12,19^, Rob Knight^13,14,20,21^, Leo Lahti^22^, Teemu J. Niiranen^1,2,23^

^1^Department of Internal Medicine, University of Turku, Turku, Finland
^2^Department of Public Health Solutions, Finnish Institute for Health and Welfare, Helsinki, Finland
^3^Transplantation and Liver Surgery Clinic, Helsinki University Hospital, University of Helsinki, Helsinki, Finland
^4^The Transplant Institute, Sahlgrenska University Hospital, Gothenburg, Sweden
^5^Department of Medicine, Kuopio University Hospital, University of Eastern Finland, Kuopio, Finland
^6^Department of Experimental Vascular Medicine, Amsterdam UMC, University of Amsterdam, Amsterdam, The Netherlands
^7^Institute for Molecular Medicine Finland, FIMM - HiLIFE, Helsinki, Finland.
^8^Cambridge Baker Systems Genomics Initiative, Baker Heart and Diabetes Institute, Melbourne, Victoria, Australia

^9^Department of Infectious Diseases, Central Clinical School, Monash University, Melbourne, Victoria, Australia

^10^Department of Clinical Pathology, The University of Melbourne, Melbourne, Victoria, Australia

^11^Department of Medicine, NAFLD Research Center, La Jolla, CA, USA

^12^Department of Medicine, University of California, San Diego, La Jolla, CA, USA

^13^Jacobs School of Engineering, University of California, San Diego, La Jolla, CA, USA

^14^Center for Microbiome Innovation, University of California San Diego, La Jolla, California, USA

^15^Collaborative Mass Spectrometry Innovation Center, Skaggs School of Pharmacy and Pharmaceutical Sciences, University of California, San Diego, La Jolla, California, USA

^16^Skaggs School of Pharmacy and Pharmaceutical Sciences, University of California, San Diego, La Jolla, California, USA

^17^Division of Biological Sciences, University of California, San Diego, La Jolla, California, USA
^18^Department of Public Health and Primary Care, Cambridge University, Cambridge, United Kingdom
^19^Department of Pharmacology, University of California San Diego, La Jolla, California, USA
^20^Department of Pediatrics, School of Medicine, University of California San Diego, La Jolla, California, USA
^21^Department of Computer Science & Engineering, University of California San Diego, La Jolla, California, USA
^22^Department of Future Technologies, University of Turku, Turku, Finland
^23^Division of Medicine, Turku University Hospital, Turku, Finland

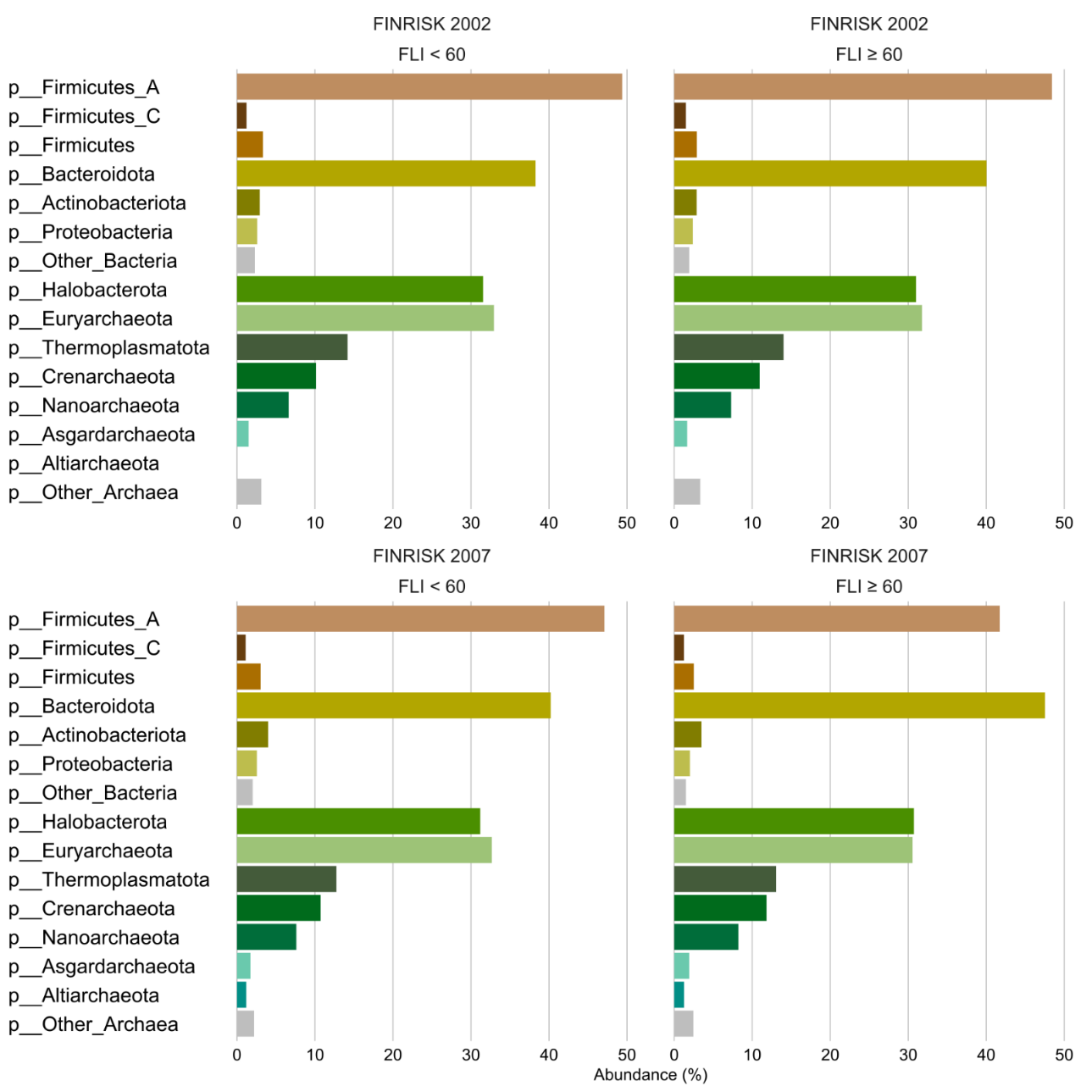

Figure S1. Baseline taxonomic composition of the FLI groups at phylum level in the main (FINRISK 2002) and validation cohort (FINRISK 2007). In each subplot, the 7 topmost phyla are bacterial, and the 8 bottom phyla are archaeal. Both bacterial and archaeal bars separately sum up to 100% total abundance (all assigned reads). Classification of the taxa follows the GTDB taxonomy.

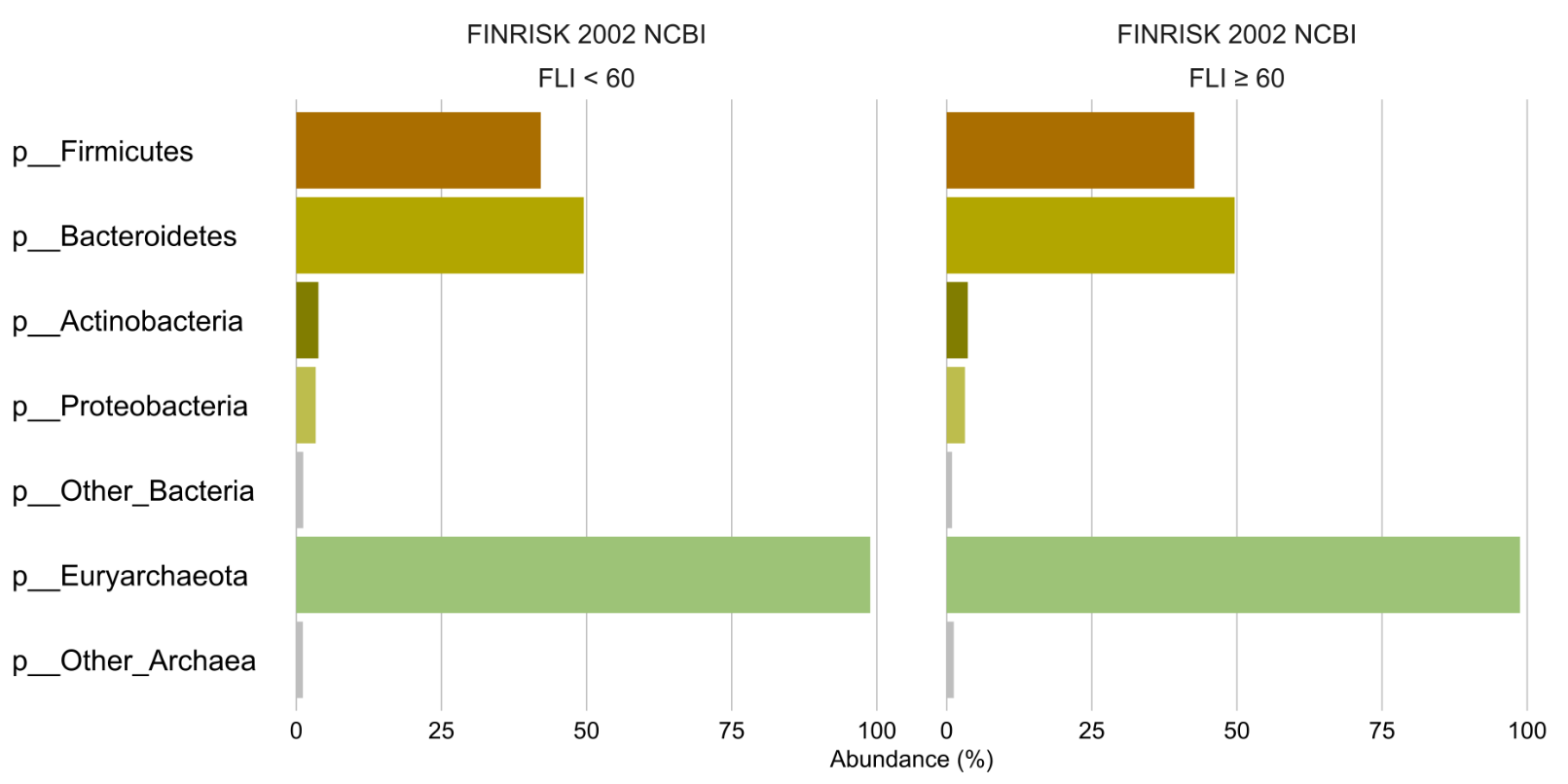

Figure S2. Baseline taxonomic composition of the FLI groups at phylum level in the main cohort (FINRISK 2002) when using SHOGUN taxonomic assignment against the NCBI RefSeq database. In the two subplots, the 5 topmost phyla are bacterial, and the 2 bottom phyla are archaeal. Both bacterial and archaeal bars separately sum up to 100% total abundance (all assigned reads). Classification of the taxa follows the NCBI taxonomy.

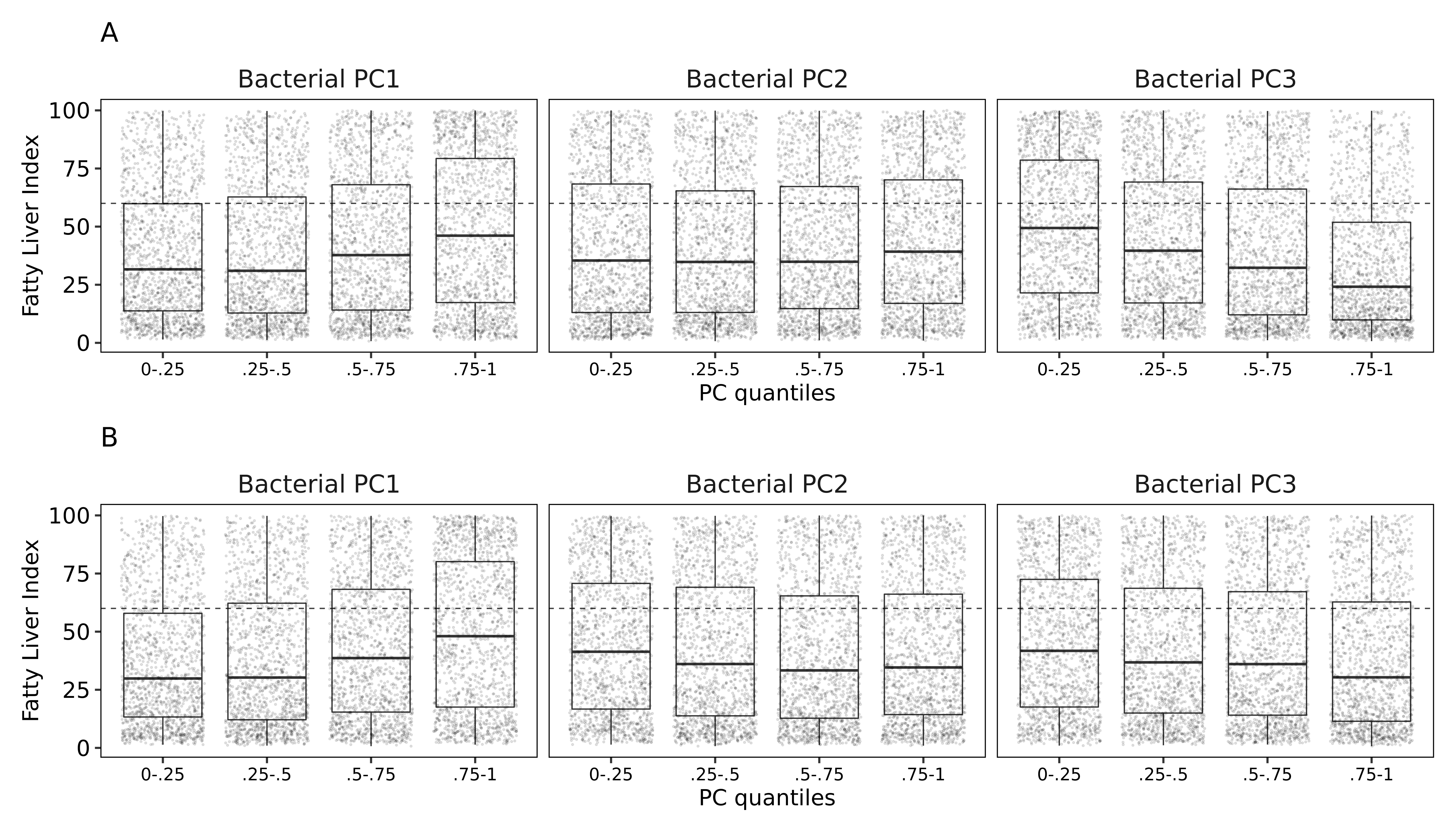

Figure S3. Comparison of the bacterial beta-diversity in FINRISK 2002 between (A) Centrifuge/GTDB annotated data with 3 first PC axes based on PhILR balances and (B) SHOGUN/NCBI annotated data with 3 first PC axes based on CLR-transformed taxa counts. When accounting for age, sex, and self-reported alcohol consumption, associations with continuous log10(FLI) are significant (uncorrected p < 0.001) in both data sets for PC1 and PC3, and in the Centrifuge/GTDB data for PC2.

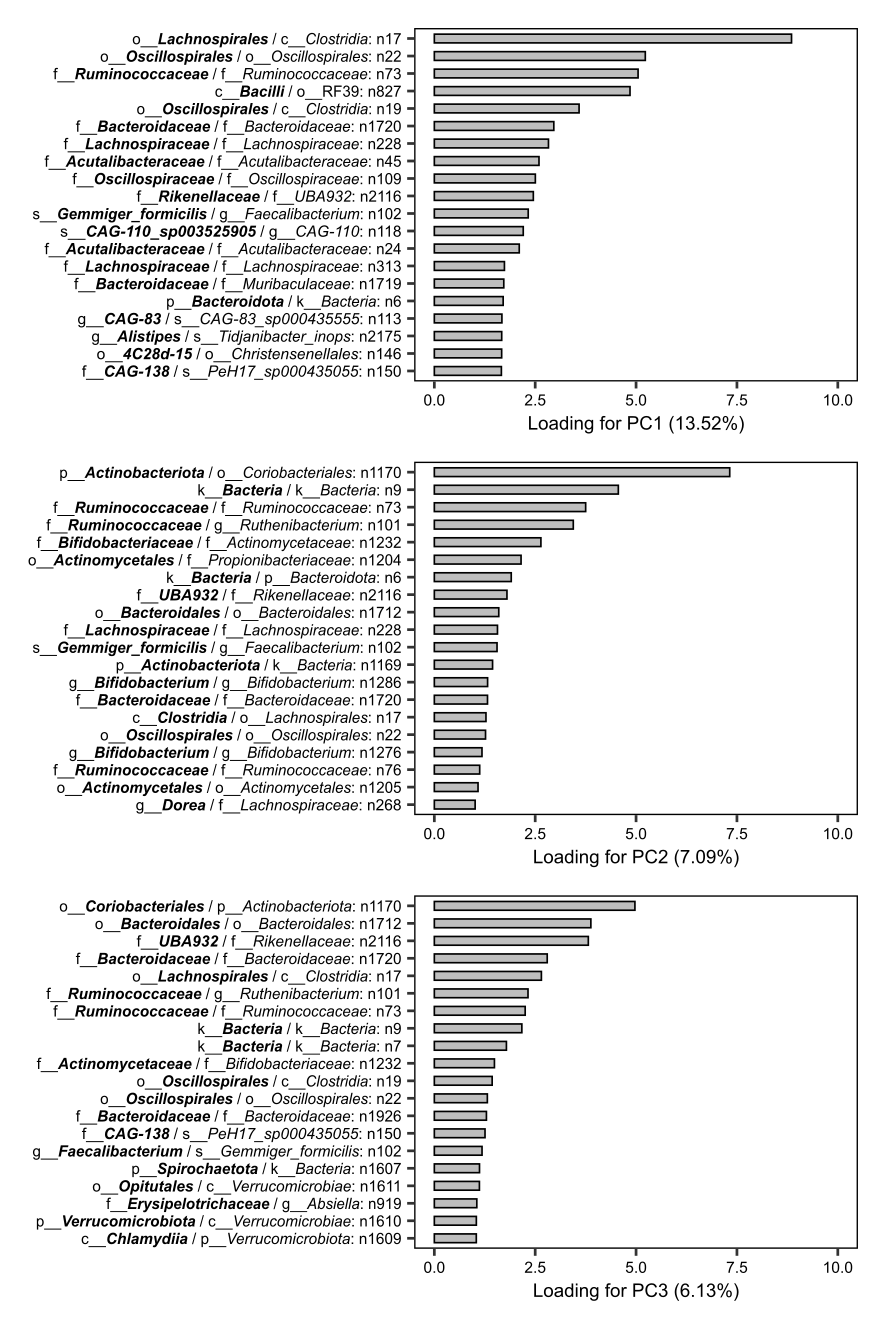
Figure S4. Loadings of the top 20 balances in the bacterial phylogenetic tree for the first principal components in FINRISK 2002 using the Centrifuge/GTDB annotated data. Taxonomic information is based on the GTDB standardized taxonomy (Parks et al., 2018) and the taxa are named based on a simple voting scheme of the two descendent clades of a given balance separately (in PhILR; Silverman et al., 2017). Balances have been rotated and the signs of their loadings on the PC axes have been switched accordingly, so that the sides of the balances indicated with bold font have a positive association with FLI. Due to the calculation of the balances, the opposite side of the balance thus has a negative association with FLI.

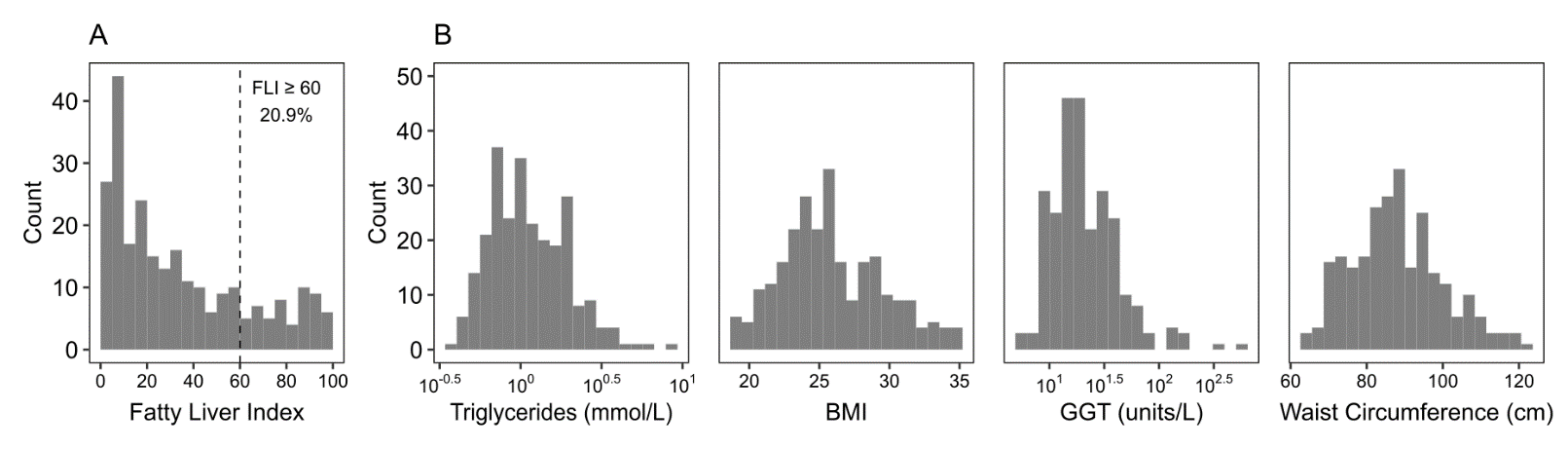
Figure S5. Distribution of FLI (A) and its components (B) in the validation cohort (FINRISK 2007). The cutoff at FLI = 60 used to divide the participants is indicated with a dashed line in panel A.

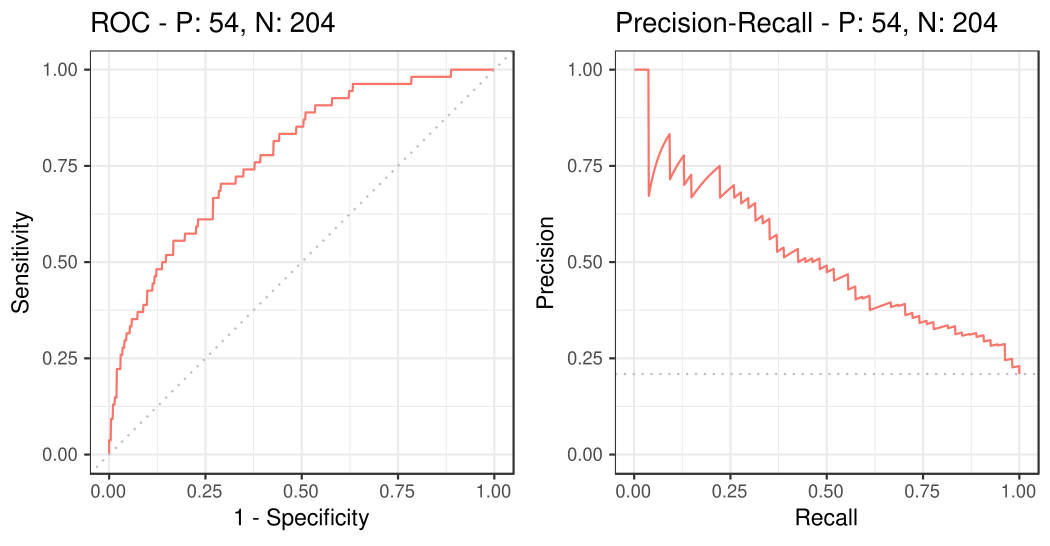
Figure S6. Receiver operating characteristic (ROC) and precision-recall curves of the 6 full LOGOCV models constructed on FINRISK 2002 data and tested on the validation cohort, *i.e*., FINRISK 2007 data. Baselines for a random classifier are shown with dashed lines. True number of participants in the high FLI group (positives; P) and in the low FLI group (negatives; N) in the validation data are indicated. For this figure, the predictions of the 6 models were averaged to be able to calculate single metrics for the combined models. With these averaged predictions, AUC = 0.78 and AUPRC = 0.51 (baseline at 0.21).

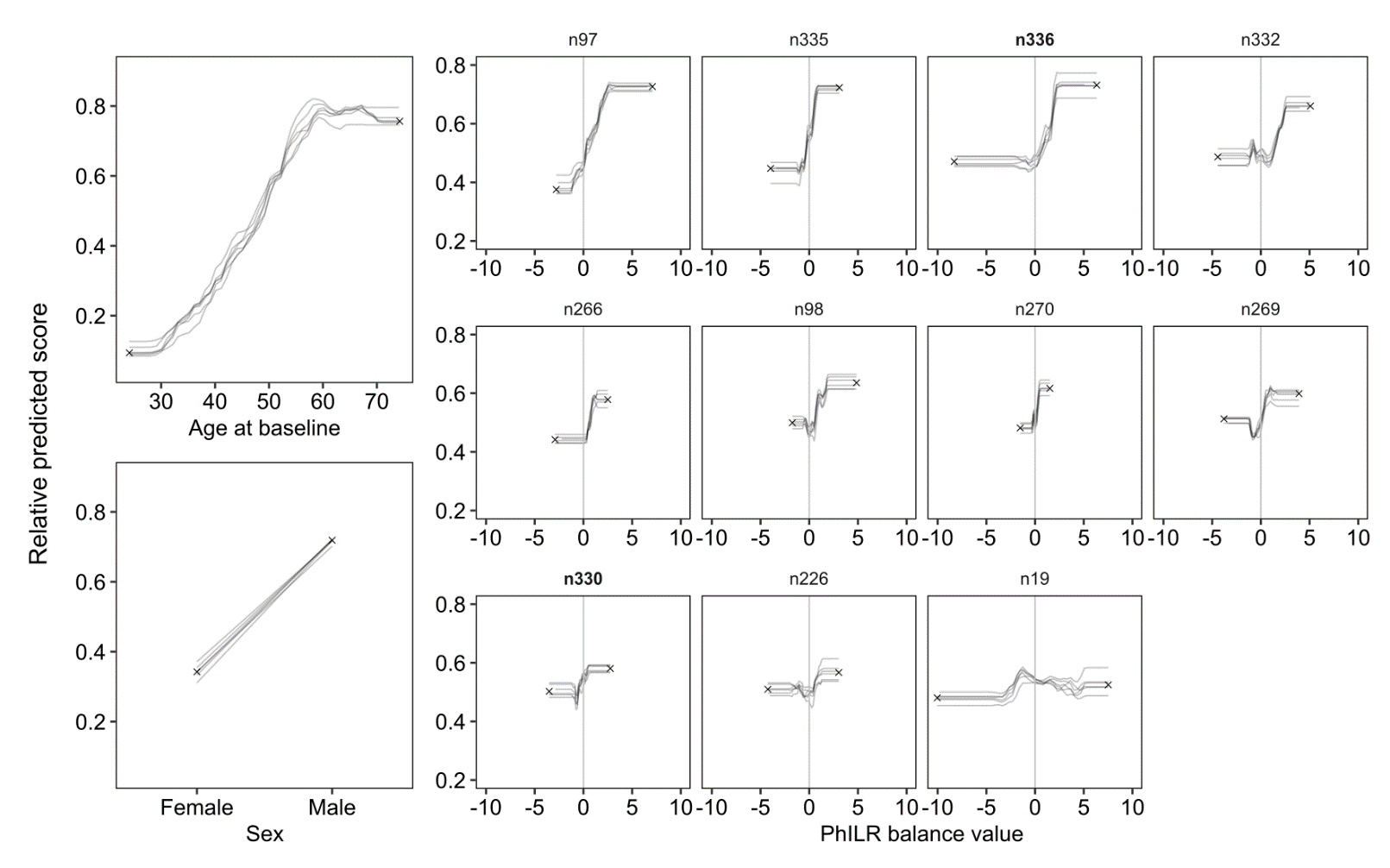
Figure S7. Partial dependence of features on the prediction outcome in the classification models discerning between the FLI < 60 and FLI ≥ 60 groups. Relative predicted score indicates the probability of classification in the FLI ≥ 60 group calculated independently of other features. Individual lines for the features show the variability of the partial dependence in the 6 LOGOCV models. The medians of the relative predicted scores at the minimum and maximum values of each feature (used in Figure 2) are shown as black crosses. The liver-specific balances are indicated with bold font. Because the signs of the PhILR balance values are dependent on the orientation of the branches at each balance (and these can be freely rotated in a phylogenetic tree), all partial dependences of the balances are shown here in the positive direction. The changes in relative predicted scores for each feature and the specific clades under each balance in this figure are also summarized in Figure 2.

**<Because of its large size, Figure S8 is supplied as an external file to retain legibility: Figure_S8.eps>**

Figure S8. Complete phylogenetic tree of taxa covered by the 11 predictive balances of the FLI classification model. Relative predicted scores in the models, number of pathways for relevant processes and presence of specific acetate and ethanol production pathways in the representative genomes are included. The maximum number of pathways is different per class of pathway. Branch distance in this cladogram is not proportional to evolutionary distance. The figure is based on the GTDB standardized bacterial taxonomy (Parks et al., 2018), and includes species and other taxa not found in the NCBI taxonomy. The liver-specific balances are indicated with bold font.

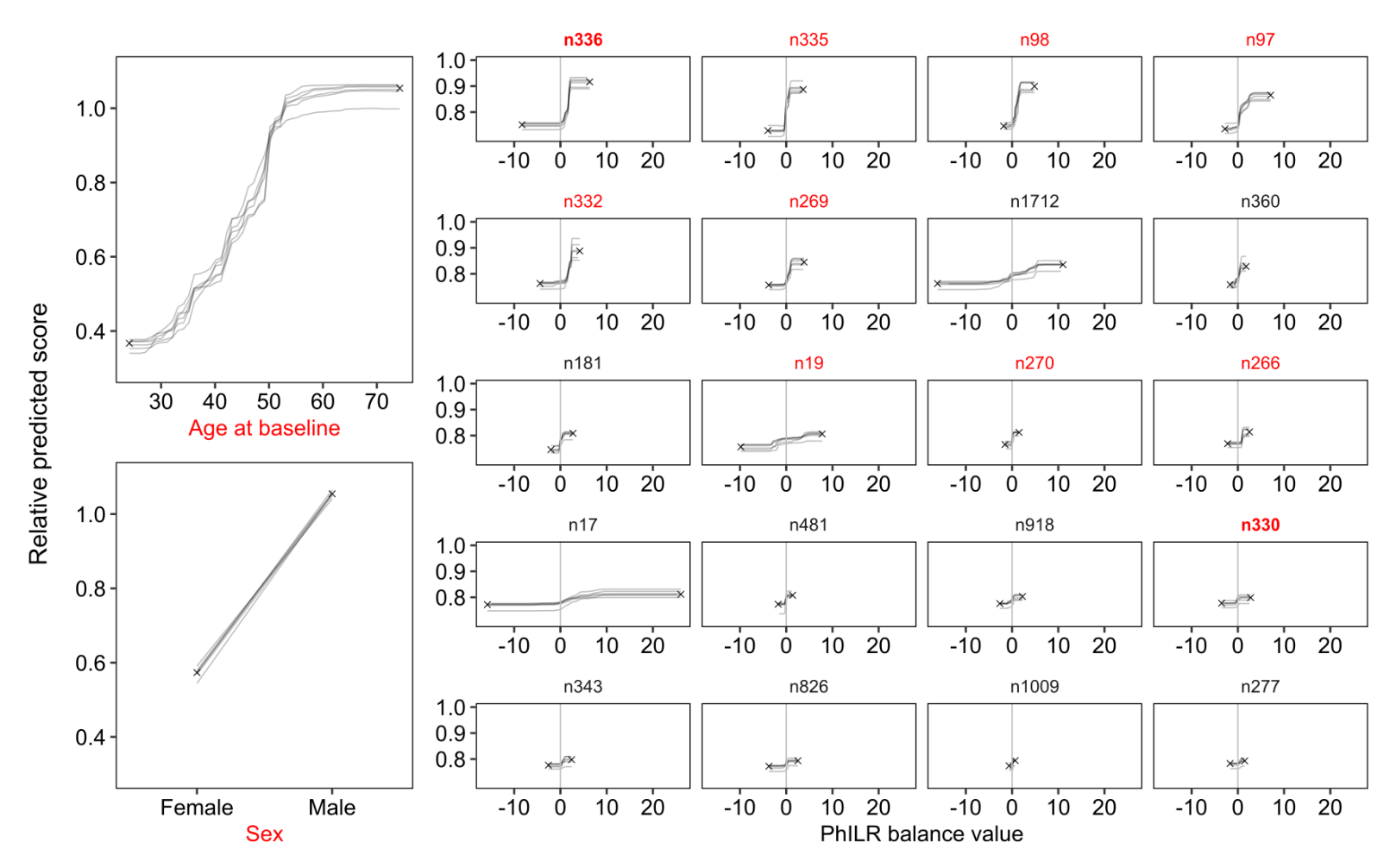
Figure S9. Partial dependence of features on the prediction outcome in the classification models discerning between FLI < 30 and FLI ≥ 60 groups. In these models, participants with FLI ≥ 30 but < 60 have been excluded. Relative predicted score indicates the probability of classification in the FLI ≥ 60 group calculated independently of other features. Individual lines for the features show the variability of the partial dependence in the 6 LOGOCV models. The medians of the relative predicted scores at the minimum and maximum values of each feature are shown as black crosses. Features in these models common with the main model are indicated with red font (only balance n226 was not included in the selected features). The liver-specific balances are indicated with bold font. Because the signs of the PhILR balance values are dependent on the orientation of the branches at each balance (and these can be freely rotated in a phylogenetic tree), all partial dependences of the balances are shown here in the positive direction.

**Table S1**. Features of the main (FINRISK 2002) and validation (FINRISK 2007) cohorts at the time of sampling. For categorical features, the numbers denote counts with percentage of total participants of each column in the brackets. For continuous features, the numbers are group means with standard deviation in the brackets.

|  | | FINRISK 2002 | | | FINRISK 2007 | | |
| --- | --- | --- | --- | --- | --- | --- | --- |
| Category | Variable | All | FLI < 60 | FLI ≥ 60 | All | FLI < 60 | FLI ≥ 60 |
|  | Participants, after exclusions | 6269 | 4359 (69.5) | 1910 (30.5) | 258 | 204 (79.1) | 54 (20.9) |
| Biometric | Age, years | 49.6 (12.9) | 47.9 (13.1) | 53.7 (11.3) | 36.1 (5.7) | 35.8 (5.8) | 36.9 (5.2) |
|  | Age, range | 24 - 74 | 24 - 74 | 24 - 74 | 24 - 44 | 24 - 44 | 24 - 44 |
|  | Sex, females | 3345 (53.4) | 2683 (61.6) | 662 (34.7) | 143 (55.4) | 131 (64.2) | 12 (22.2) |
|  | Weight, kg | 76.5 (15.2) | 70.0 (10.9) | 91.4 (13.1) | 76.1 (14.8) | 71.2 (10.6) | 94.8 (13.6) |
|  | Waist circumference, cm | 89.6 (13.4) | 83.1 (9.0) | 104.3 (9.6) | 88.1 (12.1) | 83.7 (8.9) | 104.6 (7.7) |
|  | BMI, kg/m^2^ | 27.0 (4.7) | 24.9 (3.1) | 31.7 (4.2) | 25.1 (3.6) | 24.8 (2.9) | 30.2 (2.8) |
|  | Systolic blood pressure, mmHg | 136.2 (20.3) | 133.0 (19.9) | 143.4 (19.5) | 126.4 (15.6) | 124.7 (15.4) | 132.9 (14.7) |
| Blood work | Gamma Glutamyl Transferase, units/L | 33.7 (37.1) | 24.3 (19.9) | 55.1 (54.3) | 29.3 (43.6) | 20.5 (24.2) | 62.1 (74.6) |
|  | Triglycerides, mmol/L | 1.4 (1.0) | 1.1 (0.5) | 2.1 (1.4) | 1.3 (0.9) | 1.1 (0.5) | 2.4 (1.3) |
| Lifestyle | Current smokers | 1466 (23.4) | 1025 (23.5) | 441 (23.1) | 51 (19.8) | 37 (18.1) | 14 (25.9) |
|  | High alcohol consumption^1^ | 549 (8.8) | 351 (8.1) | 198 (10.4) | 15 (5.8) | 9 (4.4) | 6 (11.1) |
|  | Diabetes | 340 (5.4) | 132 (3.0) | 208 (10.9) | 3 (1.2) | 1 (2.5) | 2 (3.7) |
|  | Cardiovascular disease | 240 (3.8) | 121 (2.8) | 119 (6.2) | 2 (0.4) | 1 (0.5) | 1 (1.9) |
| Geography | North Karelia | 1265 (20.2) | 850 (19.5) | 415 (21.7) | 147 (57.0) | 115 (56.4) | 32 (59.3) |
|  | North Savonia | 962 (15.3) | 666 (15.3) | 296 (15.5) | 0 | 0 | 0 |
|  | Turku/Loimaa | 820 (13.1) | 582 (13.4) | 238 (12.5) | 0 | 0 | 0 |
|  | Helsinki/Vantaa | 1090 (17.4) | 771 (17.7) | 319 (16.7) | 111 (43.0) | 89 (43.6) | 22 (40.7) |
|  | Oulu province | 833 (13.3) | 588 (13.5) | 245 (12.8) | 0 | 0 | 0 |
|  | Lapland | 1299 (20.7) | 902 (20.7) | 397 (20.8) | 0 | 0 | 0 |

^1^Defined as self-reported alcohol use of over > 20 g pure ethanol per day for women, or > 40 g for men.

**Table S2.** Classification model validation results with AUC. AUC are shown for all classification models and summarized per region omitted in training of the model but used for its validation. The number of participants in these validation sets are indicated in bold, and their units differ from other columns.

|  | Main model (FLI < 60 / FLI ≥ 60) | | | | Alternative model (FLI < 30 / FLI ≥ 60) | | | |
| --- | --- | --- | --- | --- | --- | --- | --- | --- |
| CV region | Full model | Covariates only | Balances  only | *N*  participants | Full model | Covariates only | Balances  only | *N*  participants |
| North Karelia | 0.761 | 0.712 | 0.671 | **379** | 0.801 | 0.762 | 0.719 | **282** |
| Lapland | 0.734 | 0.664 | 0.676 | **389** | 0.772 | 0.722 | 0.715 | **288** |
| North Savonia | 0.788 | 0.740 | 0.681 | **288** | 0.800 | 0.750 | 0.673 | **213** |
| Oulu province | 0.730 | 0.661 | 0.669 | **249** | 0.773 | 0.721 | 0.720 | **190** |
| Turku/Loimaa | 0.755 | 0.760 | 0.598 | **245** | 0.784 | 0.781 | 0.630 | **186** |
| Helsinki/Vantaa | 0.743 | 0.731 | 0.647 | **326** | 0.891 | 0.839 | 0.753 | **242** |
| Mean | 0.752 | 0.711 | 0.657 | **312.7** | 0.804 | 0.763 | 0.702 | **233.5** |

**Table S3.** Classification model validation results with AUPRC. AUPRC are shown for all classification models and summarized per region omitted in training of the model but used for its validation. The number of participants in the validation sets are indicated in bold, and the proportion of them in the FLI ≥ 60 group are indicated in italic.

|  | Main model (FLI < 60 / FLI ≥ 60) | | | | | Alternative model (FLI < 30 / FLI ≥ 60) | | | | |
| --- | --- | --- | --- | --- | --- | --- | --- | --- | --- | --- |
| CV region | Full model | Covariates only | Balances  only | *N*  participants | Proportion of  FLI ≥ 60 | Full model | Covariates only | Balances  only | *N*  participants | Proportion of  FLI ≥ 60 |
| North Karelia | 0.602 | 0.494 | 0.496 | **379** | *0.311* | 0.765 | 0.717 | 0.686 | **282** | *0.440* |
| Lapland | 0.576 | 0.456 | 0.470 | **389** | *0.314* | 0.737 | 0.643 | 0.642 | **288** | *0.424* |
| North Savonia | 0.597 | 0.469 | 0.500 | **288** | *0.285* | 0.770 | 0.680 | 0.646 | **213** | *0.446* |
| Oulu province | 0.524 | 0.385 | 0.534 | **249** | *0.273* | 0.695 | 0.570 | 0.651 | **190** | *0.384* |
| Turku/Loimaa | 0.533 | 0.522 | 0.400 | **245** | *0.298* | 0.729 | 0.739 | 0.524 | **186** | *0.366* |
| Helsinki/Vantaa | 0.553 | 0.498 | 0.430 | **326** | *0.304* | 0.813 | 0.751 | 0.646 | **242** | *0.376* |
| Mean | 0.564 | 0.471 | 0.472 | **312.7** | *0.297* | 0.752 | 0.683 | 0.633 | **233.5** | *0.406* |

**Table S4.** Performance of classification models discerning between FLI ≥ 60 and FLI < 60 groups, with data from the validation region excluded from feature selection and hyperparameter optimization. The number of participants in the validation sets are indicated in bold, and the proportion of them in the FLI ≥ 60 group are indicated in italic.

|  | AUC | | | | AUPRC | | |  | |
| --- | --- | --- | --- | --- | --- | --- | --- | --- | --- |
| CV region | Full model | Covariates only | Balances  only | Full model | | Covariates only | Balances  only | *N*  participants | Proportion of  FLI ≥ 60 |
| North Karelia | 0.755 | 0.712 | 0.670 | 0.602 | | 0.493 | 0.485 | **379** | *0.311* |
| Lapland | 0.742 | 0.670 | 0.700 | 0.585 | | 0.495 | 0.504 | **389** | *0.314* |
| North Savonia | 0.791 | 0.729 | 0.686 | 0.611 | | 0.458 | 0.494 | **288** | *0.285* |
| Oulu province | 0.737 | 0.656 | 0.676 | 0.543 | | 0.375 | 0.536 | **249** | *0.273* |
| Turku/Loimaa | 0.737 | 0.755 | 0.590 | 0.531 | | 0.495 | 0.407 | **245** | *0.298* |
| Helsinki/Vantaa | 0.749 | 0.729 | 0.668 | 0.571 | | 0.488 | 0.448 | **326** | *0.304* |
| Mean | 0.752 | 0.709 | 0.665 | 0.574 | | 0.467 | 0.479 | **312.7** | *0.297* |

**Table S5.** Performance of classification models discerning between FLI ≥ 60 and FLI < 60 groups in the external validation cohort (FINRISK 2007). The number of participants in the validation cohort are indicated in bold, and the proportion of them in the FLI ≥ 60 group are indicated in italic.

|  | AUC | | | | AUPRC | | |  | |
| --- | --- | --- | --- | --- | --- | --- | --- | --- | --- |
| CV region | Full model | Covariates only | Balances  only | Full model | | Covariates only | Balances  only | *N*  participants | Proportion of  FLI ≥ 60 |
| North Karelia | 0.773 | 0.725 | 0.688 | 0.514 | | 0.417 | 0.445 | **258** | *0.209* |
| Lapland | 0.776 | 0.728 | 0.690 | 0.522 | | 0.393 | 0.426 | **258** | *0.209* |
| North Savonia | 0.771 | 0.724 | 0.686 | 0.486 | | 0.394 | 0.423 | **258** | *0.209* |
| Oulu province | 0.780 | 0.722 | 0.684 | 0.505 | | 0.418 | 0.437 | **258** | *0.209* |
| Turku/Loimaa | 0.779 | 0.724 | 0.684 | 0.511 | | 0.403 | 0.436 | **258** | *0.209* |
| Helsinki/Vantaa | 0.767 | 0.717 | 0.684 | 0.504 | | 0.400 | 0.446 | **258** | *0.209* |
| Mean | 0.774 | 0.723 | 0.686 | 0.507 | | 0.404 | 0.436 | **258** | *0.209* |

**Table S6**. MetaCyc pathways analyzed in the representative genomes of GTDB taxa for production of ethanol and short chain fatty acids (acetate, butyrate, propionate), bile acid metabolism, and choline degradation to trimethylamine. One pathway might be included under multiple processes, based on its classification in MetaCyc.

| Process classification | MetaCyC pathway | Main substrate(s) | Product(s) | In *N* genomes |
| --- | --- | --- | --- | --- |
| Fermentation to Acetate | P41-PWY | pyruvate | **acetate**, (S)-lactate | **-** |
|  | PWY-5096 | pyruvate | **acetate**, L-alanine | **-** |
|  | PWY-5100 | pyruvate | **acetate**, (S)-lactate, CO_2_ | **-** |
|  | P142-PWY | pyruvate | **acetate**, CO_2_ | **-** |
|  | PWY-5482 | pyruvate | **acetate** | 7 |
|  | PWY-5483 | pyruvate | **acetate** | - |
|  | PWY-5485 | pyruvate | **acetate**, formate | - |
|  | PWY-5537 | pyruvate | **acetate**, succinate | - |
|  | PWY-5538 | pyruvate | **acetate**, succinate, CO_2_ | - |
|  | PWY-5600 | pyruvate | **acetate**, CO_2_ | - |
|  | PWY-5768 | pyruvate | **acetate** | - |
|  | PWY0-1312 | acetyl-CoA | **acetate** | 204 |
|  | PWY-5535 | acetyl-CoA | **acetate** | - |
|  | PWY-5536 | acetyl-CoA | **acetate** | - |
|  | P161-PWY | acetylene | **acetate**, ethanol | 10* |
|  | P124-PWY | D-glucopyranose | **acetate**, (S)-lactate | - |
|  | P461-PWY | hexitols | **acetate**, ethanol, (S)-lactate | - |
|  | PROPFERM-PWY | L-alanine | **acetate**, propanoate, ammonium, CO_2_ | - |
|  | P162-PWY | L-glutamate | **acetate**, butanoate | 42* |
|  | P163-PWY | L-lysine | **acetate**, butanoate | 12* |
| Fermentation to Butanoate | CENTFERM-PWY | pyruvate | **butanoate** | 34 |
|  | PWY-5677 | succinate | **butanoate** | 3 |
|  | PWY-5022 | 4-aminobutanoate | **butanoate**, acetate, ammonium | 8 |
|  | PWY-5676 | acetyl-CoA | **butanoate** | - |
|  | P162-PWY | L-glutamate | **butanoate**, acetate | 42* |
|  | GLUDEG-II-PWY | L-glutamate | **butanoate**, H_2_ | - |
|  | P163-PWY | L-lysine | **butanoate**, acetate | 12* |
| Fermentation to Propanoate | P108-PWY | pyruvate | **propanoate** | 7 |
|  | PWY-5494 | (R)-lactate | **propanoate** | - |
|  | PWY-8086 | (S)-lactate | **propanoate**, acetate, H_2_ | 1 |
|  | PWY-7013 | propylene glycol | **propanoate**, propan-1-ol | 6 |
|  | PROPFERM-PWY | L-alanine | **propanoate**, acetate, ammonium, CO_2_ | - |
|  | PWY-5088 | L-glutamate | **propanoate** | - |
| Fermentation to Ethanol | PWY-5480 | pyruvate | **ethanol**, formate | 68 |
|  | PWY-5486 | pyruvate | **ethanol** | - |
|  | PWY-6587 | pyruvate | **ethanol** | 25 |
|  | FERMENTATION-PWY | phosphoenolpyruvate | **ethanol**, acetate, succinate, 2-oxoglutarate, (R)-lactate, CO_2_, H_2_ | 6 |
|  | P161-PWY | acetylene | **ethanol**, acetate | 10* |
|  | P122-PWY | β-D-fructofuranose, D-glucopyranose | **ethanol**, (S/R)-lactate, CO_2_ | 34 |
|  | P461-PWY | hexitols | **ethanol**, acetate, (S)-lactate | - |
| Bile Acid Degradation | PWY-7754 | cholate / chenodeoxycholate | **deoxycholate /  lithocholate** | 98 |
|  | PWY-6518 | cholate | **7-epicholate**, **12-epicholate, isocholate** | 8 |
| Choline Degradation to TMA | PWY-7167 | choline | **trimethylamine** | 24 |
|  | PWY-3641 | L-carnitine | **trimethylamine**, pyruvate | - |

*Count duplicated because pathway is included under more than one process classification.

**<Because of its large size, Table S7 is supplied as an external file to retain legibility: Table_S7.xlsx>**

**Table S7.** Individual taxa in FINRISK 2002 positively and negatively associated with the high FLI (≥ 60) group in the ALDEx2 analysis. Results are sorted by effect size and divided to positive and negative associations separately in the Centrifuge/GTDB annotated data (our primary data set) and SHOGUN/NCBI annotated data. All taxa with a significant association (*Q* value < 0.001) with the high FLI group are included in this table.
